# Feature Extraction Tool Using Temporal Landmarks in Arterial Blood Pressure and Photoplethysmography Waveforms

**DOI:** 10.1101/2025.03.20.25324325

**Authors:** Ravi Pal, Akos Rudas, Tiffany Williams, Jeffrey N. Chiang, Anna Barney, Maxime Cannesson

## Abstract

Arterial blood pressure (ABP) and photoplethysmography (PPG) waveforms both contain vital physiological information for the prevention and treatment of cardiovascular diseases. Extracted features from these waveforms have diverse clinical applications, including predicting hyper- and hypo-tension, estimating cardiac output from ABP, and monitoring blood pressure and nociception from PPG. However, the lack of standardized tools for feature extraction limits their exploration and clinical utilization. In this study, we propose an automatic feature extraction tool that first detects temporal location of landmarks within each cardiac cycle of ABP and PPG waveforms, including the systolic phase onset, systolic phase peak, dicrotic notch, and diastolic phase peak using the iterative envelope mean method. Then, based on these landmarks, extracts 852 features per cardiac cycle, encompassing time-, statistical-, and frequency-domains. The tool’s ability to detect landmarks was evaluated using ABP and PPG waveforms from a large perioperative dataset (MLORD dataset) comprising 17,327 patients. We analyzed 34,267 cardiac cycles of ABP waveforms and 33,792 cardiac cycles of PPG waveforms. Additionally, to assess the tool’s real-time landmark detection capability, we retrospectively analyzed 3,000 cardiac cycles of both ABP and PPG waveforms, collected from a Philips IntelliVue MX800 patient monitor. The tool’s detection performance was assessed against markings by an experienced researcher, achieving average F1-scores and error rates for ABP and PPG as follows: (1) On MLORD dataset: systolic phase onset (99.77 %, 0.35 % and 99.52 %, 0.75 %), systolic phase peak (99.80 %, 0.30 % and 99.56 %, 0.70 %), dicrotic notch (98.24 %, 2.63 % and 98.72 %, 1.96 %), and diastolic phase peak (98.59 %, 2.11 % and 98.88 %, 1.73 %); (2) On real time data: systolic phase onset (98.18 %, 3.03 % and 97.94 %, 3.43 %), systolic phase peak (98.22 %, 2.97 % and 97.74 %, 3.77 %), dicrotic notch (97.72 %, 3.80 % and 98.16 %, 3.07 %), and diastolic phase peak (98.04 %, 3.27 % and 98.08 %, 3.20 %). This tool has significant potential for supporting clinical utilization of ABP and PPG waveform features and for facilitating feature-based machine learning models for various clinical applications where features derived from these waveforms play a critical role.

## Introduction

In this paper, we present a novel feature extraction tool for analyzing Arterial Blood Pressure (ABP) and Photoplethysmography (PPG) waveforms, capable of working with both recorded and real-time data. Cardiovascular diseases (CVDs) are recognized as the leading global cause of mortality, according to the World Health Organization^1,2,3^, responsible for 17.9 million deaths annually and constituting 31% of all deaths^2^. These diseases significantly alter the ABP waveform^6,7^, which contains valuable pathophysiological information essential for diagnosis and prevention^6^. Continuous monitoring of the ABP is the most commonly used measure for assessing hemodynamic stability^4^ and is crucial for critically ill patients with CVDs^5^. PPG, commonly referred to as the pulse oximetric wave, is a non-invasive technique used to evaluate blood oxygen levels (SpO2)^8^. Its waveform provides further insights into cardiovascular health, reflecting changes in the cardiovascular system^9^. Although, primarily used in anesthetic monitoring, over the past decade there has been a growing utilization of PPG to assess cardiovascular status^3,10^.

In critical care settings such as Intensive Care Units and operating rooms, invasive blood pressure monitoring plays a crucial role serving as the gold standard for continuous blood pressure measurement^11,12^. It helps to quickly diagnose cardiovascular issues and indicates how patients are responding to medications aimed at preventing complications that can disrupt blood flow to tissues^13^ such as hypo- or hyper-tension^11^. ABP can be measured at various anatomical sites, such as the radial, brachial, or femoral arteries, by inserting a catheter^4,14^. The radial artery is the most commonly used location for continuous ABP measurement in clinical practice due to technical simplicity and lower risk of major complications^14^. The characteristics of the ABP waveform change with physiology, measurement site and age^4^, and the peaks and valleys in the ABP waveform reflect different functions of the left side of the heart^11^. This has led to increased interest in ABP waveform analysis, which enables the evaluation of various critical measures such as vascular resistance, left ventricular stroke volume (SV), SV variation, and pulse pressure fluctuation during positive pressure respiration^11^.

The PPG waveform closely mirrors the ABP waveform but represents changes in microvascular tissue bed volume due to circulating blood pressure, rather than reflecting pressure directly^9,10^. This waveform provides a wealth of information about heart health, blood vessel dynamics, breathing patterns, and the autonomic nervous system^15^. It can be measured at various peripheral areas of the body, such as the fingers, ears, wrists, and toes. In clinical settings, the signal is usually recorded using a pulse oximeter on the index finger^9,10^. The rising popularity of smart wearables, capable of monitoring the PPG, offers increasing opportunities to track health and fitness in everyday life as devices, ranging from fitness bands to smart rings and smart watches, become ever more common^16,17^. Several factors including gender, age, measurement site, and health conditions may change the PPG waveform’s characteristics^10,16,17^, thus analysis of a patient’s PPG waveform could facilitate the extraction of a range of physiological measures useful for precise, personalized health assessment.

A cardiac cycle of ABP/PPG waveform can be categorized into two main phases—systolic phase and diastolic phase—defined by five temporal landmarks: systolic phase onset, systolic phase peak, dicrotic notch, diastolic phase peak, and diastolic phase endpoint (see figure 1). The systolic phase begins with the onset of the heartbeat, marking both the start of the cycle and the endpoint of the previous cycle^10^. This phase, spanning from onset to the dicrotic notch, reflects the rapid increase in arterial pressure as blood is ejected from the left ventricle, causing the arterial walls to expand^10^. The systolic peak represents the highest point of the pressure waveform^18^. Following the systolic peak, there is a noticeable drop in pressure with the end point of this phase marked by a local minimum known as the dicrotic notch, which signifies the moment when the aortic valves close^4,10,18^. The dicrotic notch indicates the shift from the cardiac systolic (contraction) phase to the diastolic (relaxation) phase^19^. During the diastolic phase, arterial pressure gradually decreases until it reaches its lowest point: the end of the diastolic phase, which is marked by the next systolic onset. These events within a cardiac cycle generate critical landmarks for extracting valuable physiological features from the waveforms.

**Figure 1.**
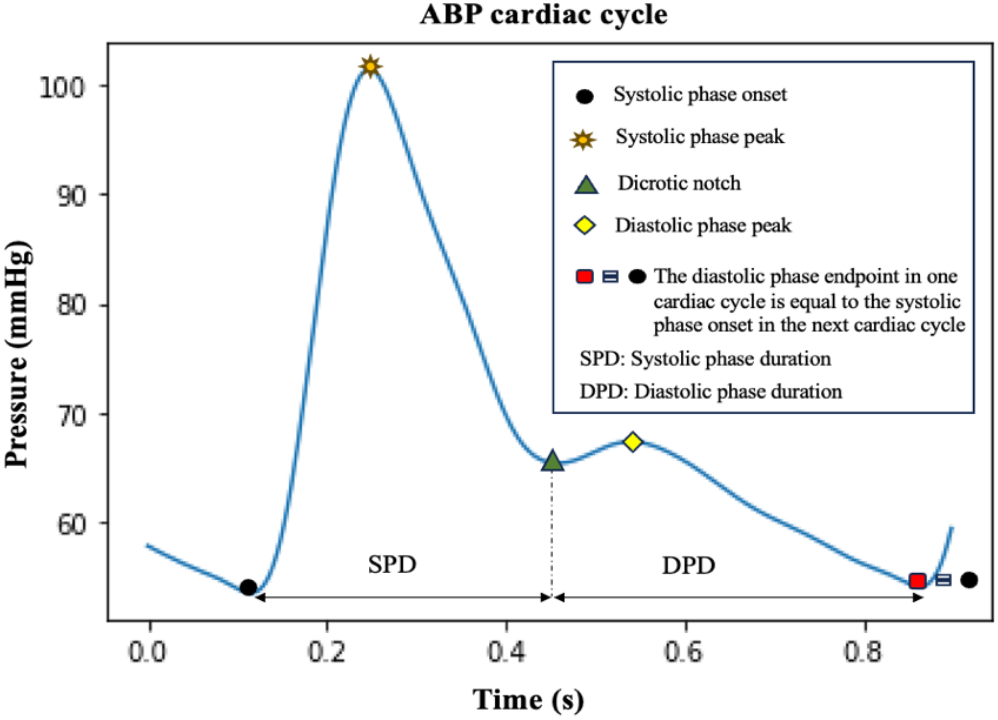
Illustration of the arterial blood pressure cardiac cycle, highlighting landmarks: systolic phase onset, systolic phase peak, dicrotic notch, diastolic phase peak, and diastolic phase endpoint, which corresponds to the onset of the systolic phase in the next cardiac cycle.

Detailed analysis of ABP waveforms provides deeper insight into overall hemodynamic status^20^, allowing prediction of episodes of hypo- or hyper-tension^21^ and estimation of cardiac output^22^. Unlike the invasive ABP measurement, PPG technology offers a non-invasive approach widely adopted in consumer wearables and medical devices^23^. In smartwatches, it is primarily utilized for monitoring heart rate, while in pulse oximeters, it measures blood oxygen saturation levels^15^. In more sophisticated wearables, its uses also include noninvasive blood pressure estimation^24,25^, atrial fibrillation detection^18,26^, sleep stage monitoring ^23,27^, mental stress detection^28,29^, and identification of sleep apnea^30,31^. In clinical use, PPG-based pulse oximetry is a standard method found in healthcare environments from intensive care units to home monitoring systems. In each instance, PPG signal analysis plays a crucial role in deriving meaningful physiological data. Several tools have been proposed for the analysis of ABP/PPG waveforms^15, 32-45^. However, these tools are often developed for the analysis of either PPG or ABP waveforms alone, and some cannot detect all landmarks within a cardiac cycle. Furthermore, some tools are limited to landmarks detection and cannot extract features from these waveforms (see Table 1).

**Table 1.** Different published algorithms/toolboxes for analyzing PPG/ABP waveforms.

| Tools | SPO<sub>D</sub> | SPP<sub>D</sub> | DN<sub>D</sub> | DPP<sub>D</sub> | F<sub>E</sub> | D<sub>PPG</sub> | D<sub>ABP</sub> | Q<sub>V</sub> |
| --- | --- | --- | --- | --- | --- | --- | --- | --- |
| ABP waveform delineator [32] | ✓ | ✓ | ✓ | X | X | X | ✓ | ✓ |
| Hilbert transform based approach [33] | ✓ | ✓ | X | X | X | ✓ | X | ✓ |
| Systolic peak detector [34] | X | ✓ | X | X | X | ✓ | X | ✓ |
| IEM-based dicrotic notch detection algorithm [35] | X | X | ✓ | X | X | ✓ | ✓ | ✓ |
| Physics-aware approach [36] | X | X | ✓ | X | X | X | ✓ | ✓ |
| pyPPG [15] | ✓ | ✓ | ✓ | ✓ | ✓ | ✓ | X | ✓ |
| PulseAnalyse [37] | ✓ | ✓ | ✓ | ✓ | X | ✓ | ✓ | X |
| PPGFeat [38] | ✓ | ✓ | ✓ | ✓ | X | ✓ | X | ✓ |
| NeuroKit2 [39] | ✓ | ✓ | X | X | X | ✓ | X | X |
| RRest [40] | ✓ | ✓ | X | X | X | ✓ | X | X |
| PPGSynth [41] | ✓ | X | X | X | X | ✓ | X | X |
| PhysioNet [42] | ✓ | ✓ | X | X | X | ✓ | ✓ | ✓ |
| HeartPy [43,44] | ✓ | X | X | X | X | ✓ | X | ✓ |
| BioSPPy [45] | ✓ | ✓ | X | X | X | ✓ | ✓ | X |

SPO<sub>D</sub>: systolic phase onset detection; SPP<sub>D</sub>: systolic phase peak detection; DN<sub>D</sub>: Dicrotic notch detection; DPP<sub>D</sub>: Diastolic phase peak detection; KD<sub>WFSD</sub>: Key points detections without first and second derivatives; F<sub>E</sub>: Feature extraction; D<sub>PPG</sub>: Developed for PPG waveform analysis; D<sub>ABP</sub>: Developed for arterial blood pressure waveform analysis; Q<sub>V</sub>: Quantitative validation; - = attribute not reported; X = No; ✓ = Yes.

In this paper we introduce a novel feature extraction tool, developed in the Python programming language. The tool works through two main stages: first, it detects the landmarks within a cardiac cycle of an ABP or PPG waveform using the iterative envelope mean (IEM) method^35,46^; second, it extracts 852 features per cardiac cycle using the detected landmarks. The IEM method decomposes a signal into two components: stationary and non-stationary ^46^. The tool uses the non-stationary component of the IEM method for its analysis. Note that recently, our group proposed an IEM-based algorithm to detect the dicrotic notch within a cardiac cycle, both when it is clearly visible and when it cannot be visually detected (DN-less cardiac cycles)^35^. In our current study, the proposed tool not only identifies the dicrotic notch but aims to detect the temporal location of all landmarks within a cardiac cycle (see figure 1). These landmarks are then used to extract salient features from each ABP or PPG cycle. The main contributions of this study are as follows:

- The tool can detect the temporal location of landmarks within a cardiac cycle in both the ABP and PPG waveforms.
- Using the detected landmarks, the tool can extract 852 features per cardiac cycle from the waveform, including time-, statistical-, and frequency-domain features.
- The tool has been evaluated using the large perioperative MLORD dataset^47^, which comprises data from 17,327 patients, as well as real-time data from the Philips IntelliVue MX800 bedside patient monitor in demonstration mode.
- To make it easier for researchers and clinicians to use our feature extraction tool for analyzing ABP and PPG waveforms, we designed a graphical user interface (GUI) that enables users to extract features, visualize detected landmarks within the cardiac cycles of ABP or PPG waveforms, and interact with its various functions such as pausing, playing, navigating to the previous window, or advancing to the next.

## Methods

### Description of the feature extraction tool

The steps taken by the feature extraction tool are (see figure 2(i)): (1) pre-processing the ABP/PPG signals, (2) detecting the temporal location of all landmarks using the non-stationary component of the IEM method, and (3) extracting 852 features per cardiac cycle based on the identified landmarks. Note, as shown in figure 1, the end of the diastolic phase marks the beginning of the systolic phase in the next cardiac cycle. Therefore, in this study, we have not separately evaluated the tool’s ability to detect the diastolic phase endpoint.

**Figure 2.**
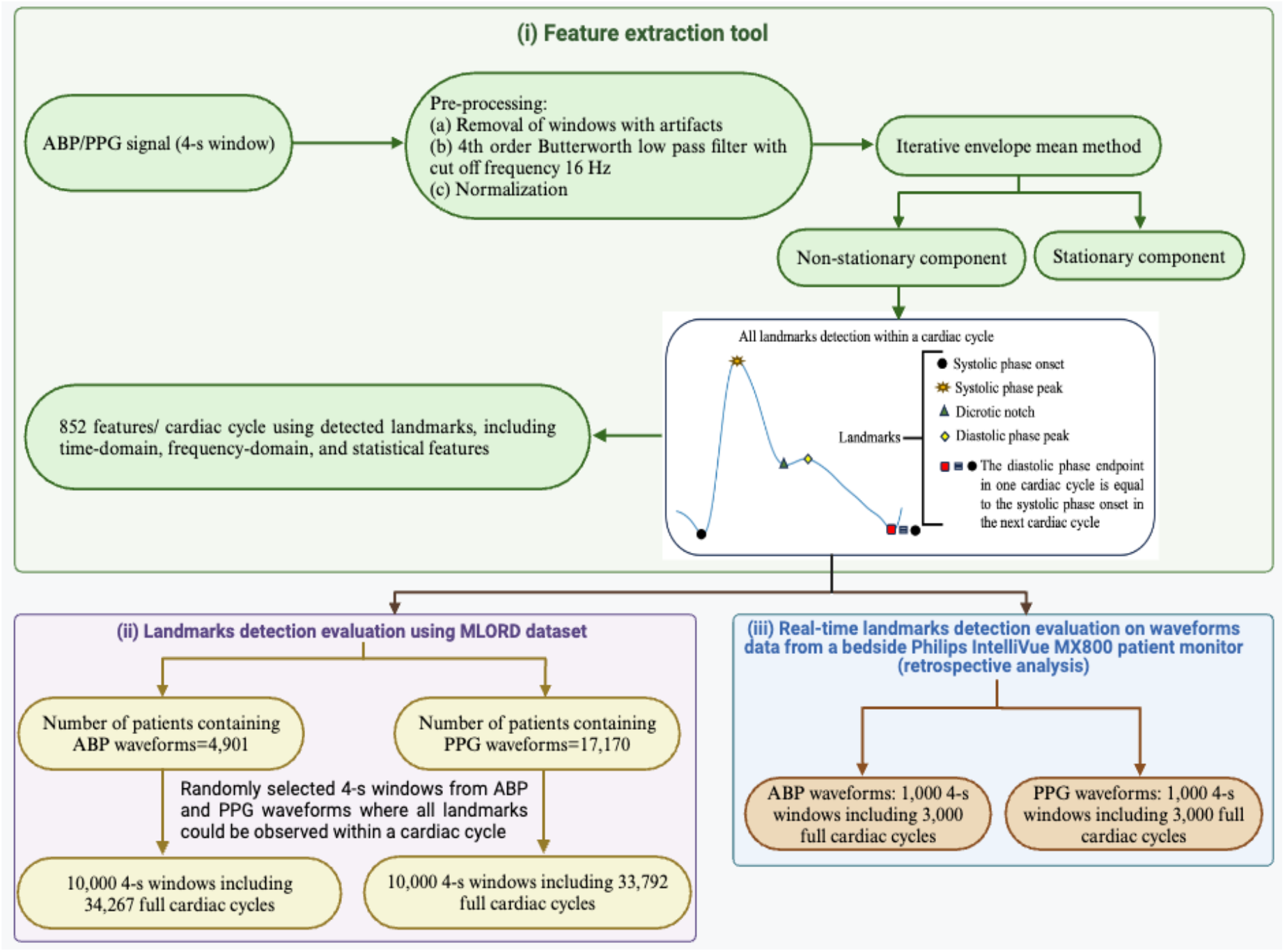
(i) Block diagram of the feature extraction tool; (ii) Landmark detection evaluation of the feature extraction tool using the MLORD dataset; (iii) Real-time landmark detection evaluation of the feature extraction tool using the waveform data from a bedside Philips IntelliVue MX800 patient monitor (retrospective analysis).

### Pre-processing

Firstly, our tool pre-processes non-overlapping 4-s windows of the ABP or PPG waveforms. As an example, figures 3 (i-a) and 3 (ii-a) illustrate a 4-s window of the ABP and PPG waveforms, respectively, each containing five complete cardiac cycles. In the pre-processing step, any window with artifacts was first removed from the dataset based on the following criteria: containing zero or negative values, or having fewer than three or more than ten peaks exceeding the 75th percentile of the window’s amplitude^35^. Once a window passed the artifact removal step, it was filtered using a 4th-order Butterworth low-pass filter with a cutoff frequency of 16 Hz to eliminate high-frequency noise^35^. Finally, the filtered signal was normalized using Eq.1.

**Figure 3.**
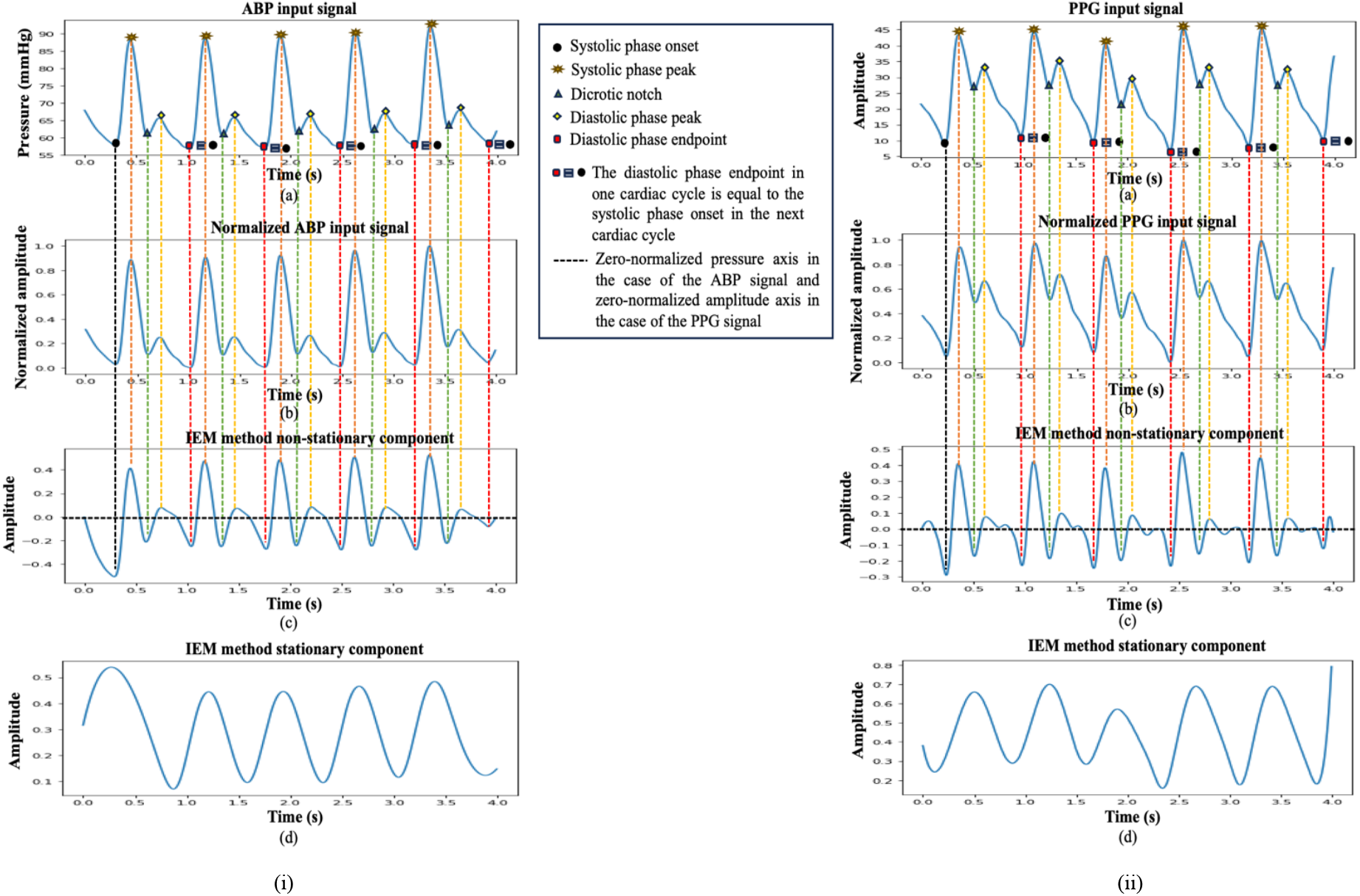
Illustrates the process of landmark detection within a cardiac cycle for (i) ABP and (ii) PPG signals using the feature extraction tool: Plots labeled (a) shows 4-s input signal windows with landmarks marked by an expert researcher, (b) displays normalized signals, (c) illustrates the non-stationary components derived using the IEM method, and (d) represents the stationary components obtained through the IEM method.

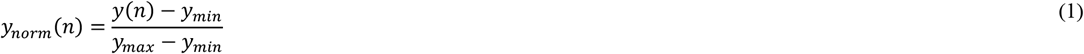

where *y*(*n*) is the input signal, *n* is the sample index in the input signal (*n* = 1,2, …. *N*), *y*_*min*_ and *y*_*max*_ represent the minimum and maximum values of the input signal, respectively, and *y*_*norm*_(*n*) denotes the normalized isignal. The normalized signals for the ABP and PPG waveforms are shown in Figures 3 (i-b) and 3 (ii-b), respectively.

### Automatic landmarks detection within a cardiac cycle

We used the IEM method to prepare the signal for detection of cardiac cycle landmarks^35,46^ because it can reveal the temporal location of landmarks, not only when they are visible in the input signal but also when they are less visible or cannot be directly interpreted from the input signal, especially the dicrotic notch^35^ and diastolic phase peak landmarks. The IEM method is a decomposition technique that separates an input signal into estimates of its non-stationary and stationary components. The process begins by smoothing the input signal and calculating the sample-by-sample mean of its upper and lower envelopes. This mean is then subtracted from the original signal, producing a new signal that serves as the input for the next iteration. By repeating this process for a specified number of iterations (*J*), the method estimates the non-stationary component (NSTS) of the original signal. Additionally, summing the envelope means from each iteration provides an estimate of the stationary component (STS) of the original signal. The IEM method has previously been used in conjunction with the fractal dimension method (IEM-FD filter) for analyzing lung sounds^46^ and for detecting the dicrotic notch in ABP and PPG waveforms, both when the notch is clearly visible and when it is not^35^. As a new application, this study utilizes the IEM method to detect the temporal location of all landmarks within the cardiac cycle of ABP and PPG waveforms. The IEM method has some data-driven parameters that need to be selected: i) the number of coefficients of the Savizky-Golay (SG) smoothing filter. Set to twice the duration of the shortest feature of interest in the signal, which for the inter-landmark duration is approximately 25 samples; and ii) the accuracy level for the stopping criteria, here set to 0.1. For a detailed explanation of the IEM method’s working process, including its mathematical formulation, and its evaluation we refer readers to our previous article^35,46^.

Figures 3(i-c) and 3(ii-c) display the nonstationary (NSTS) components found by the IEM method in the ABP and PPG waveforms, respectively, while figures 3(i-d) and 3(ii-d) demonstrate the stationary components of the IEM method in ABP and PPG waveforms, respectively. In figure 3, vertical dashed lines indicate landmark positions located within a cardiac cycle by visual inspection, while the black horizontal line in the NSTS component found by the IEM method represents the zero-normalized pressure axis.

The IEM method’s NSTS can reveal all the landmarks within a cardiac cycle; however, automatically locating their temporal positions involves several additional steps requiring precise identification of the relevant peaks and valleys. Challenges arise because pressure reflections within the arterial system can generate secondary waves, which frequently appear between the systolic peak and diastolic endpoint in the ABP waveform, creating additional peaks and valleys within each cardiac cycle^35^. Non-physiological oscillations in the PPG and ABP waveforms can introduce yet more peaks and valleys, further complicating the task of accurately determining landmarks. Therefore, while the IEM method is effective in revealing these landmarks, accurately distinguishing them amidst the multiple peaks, valleys, and oscillations requires careful analysis and additional processing steps. To address these issues, we introduced the following conditions: (1) In NSTS, the valley corresponding to the dicrotic notch and the peak corresponding to the diastolic phase peak must be at least 0.1s (25 samples) away from the peak and valley corresponding to the systolic phase peak and diastolic phase endpoint, respectively. (2) The y-axis values of the peaks corresponding to the systolic phase peak and diastolic phase peak must be greater than zero, while the y-axis values of the valleys corresponding to the systolic phase onset, and dicrotic notch, and diastolic phase endpoint must be less than zero.

### Feature extraction using the detected landmarks

The tool uses the temporal location of detected landmarks to extract features from the cardiac cycles of ABP or PPG waveforms, including time domain, frequency domain, and statistical features. These features include amplitude features (n_1_=30), amplitude ratio features (n_1_=210), duration features (n_1_=10), duration ratio features (n_1_=46), average features (n_1_=40), median features (n_1_=20), root mean square features (n_1_=20), skewness (n_1_=1; measures how asymmetrical a distribution is), kurtosis (n_1_=1; assesses the sharpness of the peak in the distribution curve), area features (n_1_=40), area ratio features (n_1_=180), as well as systolic rise phase width (SRPW) and overall decay phase width (ODPW) features (n_1_=36) measured at different percentages of a cardiac cycle in the ABP/PPG waveform. In addition, the tool extracts frequency-domain features (n_1_=12), first derivative features (n_1_=28), and second derivative features (n_1_=178) per cardiac cycle. We used code written in the Python programming language to extract each feature. Figure 4 illustrates 25-time domain amplitude and duration features. Their descriptions are provided in table 2. Note that the complete detailed description of all extracted features (n_1_=852) is provided in the supplementary document.

**Table 2.**
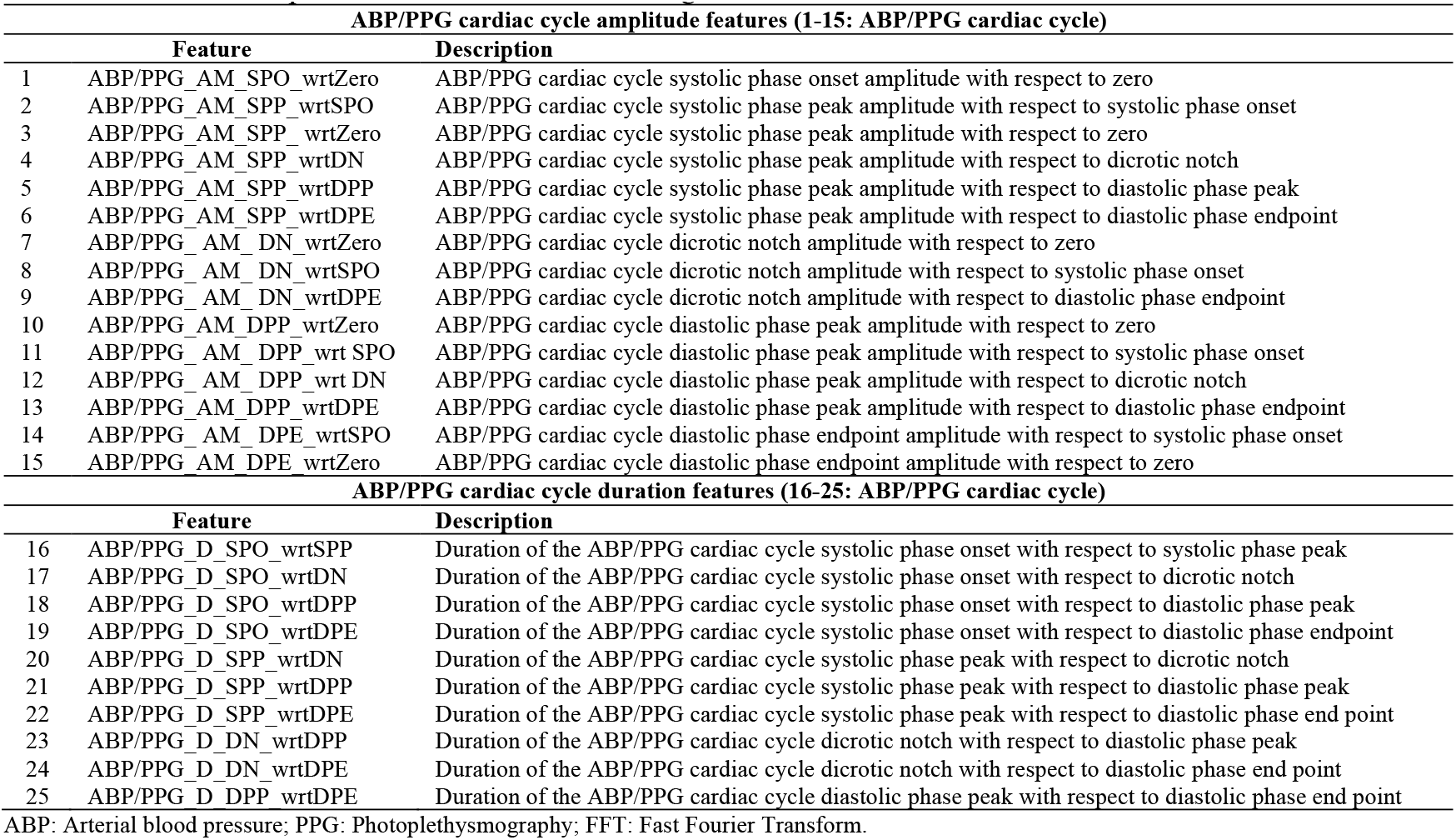
Detailed descriptions of the features shown in figure 4.

**Figure 4.**
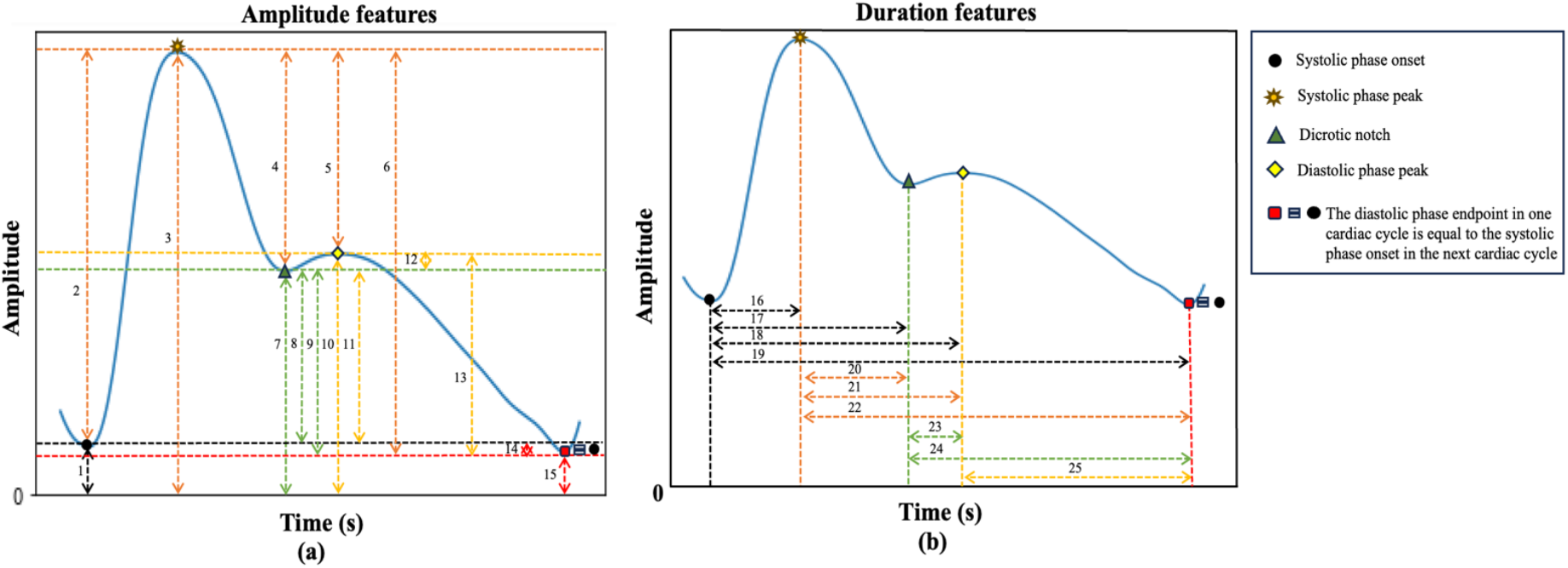
This figure illustrates a subset of time domain features extracted using detected landmarks from the PPG cardiac cycle by the tool. It highlights 25 features out of the total of 852. These features are categorized as follows: (a) Amplitude features (1-15), and (b) Duration features (16-25). Detailed descriptions of these features are provided in table 2.

### Graphical user interface

Additionally, we developed a graphical user interface (GUI) to facilitate interaction between researchers and our feature extraction tool. The GUI provides several key functions: mode selection (recorded data or real-time), waveform upload (ABP or PPG), and display of the waveform sampling frequency, along with an empirically set condition for the feature extraction tool. Users can view the detected landmarks within the cardiac cycle of an ABP/PPG waveform and generate a .csv file containing 852 features per cardiac cycle. The interface is designed for easy navigation, offering options to pause, play, view the previous window, advance to the next, or jump to a specific time point. For ABP waveforms, the GUI also displays detailed information, including systolic blood pressure, diastolic blood pressure, and mean arterial pressure for each complete cardiac cycle. Figures 5(a) and 5(b) show the GUI wiht examples of PPG and ABP waveforms, respectively.

**Figure 5.**
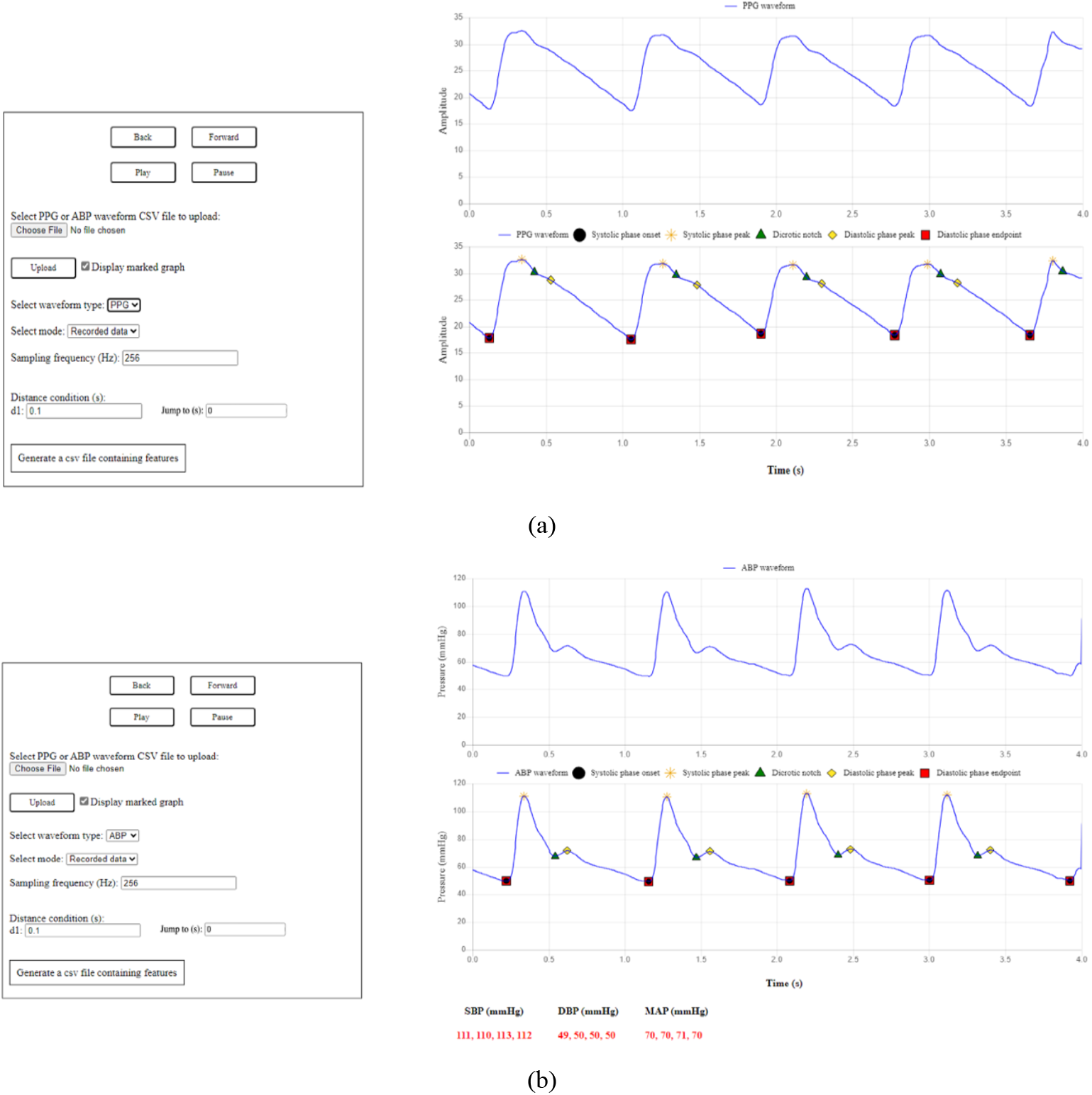
Displays the graphical user interface (GUI) of the feature extraction tool: (a) PPG waveform and (b) ABP waveform.

### Description of evaluation datasets

The MLORD dataset^47^, which includes data from 17,327 patients who underwent surgeries between 2019 and 2022 at the David Geffen School of Medicine at the University of California Los Angeles (UCLA), was used to evaluate the performance of the feature extraction tool. The dataset collection protocol for MLORD was approved by the UCLA Institutional Review Board (IRB# 19–000354). This large perioperative dataset contains both clinical and waveform data. Clinical data were obtained from electronic health record (EHR) systems, specifically Epic (Verona, WI, USA) and Surgical Information Systems (Alpharetta, GA, USA), while waveform data were recorded directly in the operating room using the Bernoulli data collection system (Cardiopulmonary, New Haven, CT, USA). The waveform data comprise over 72,264 hours of recordings, totaling 7.6 TB in size, and include digital physiological waveforms such as electrocardiograms (ECG), ABP, and PPG. For our study, we focused on ABP and PPG waveforms, both sampled at 256 Hz. Out of the 17,327 patients in the MLORD dataset, 4,901 have ABP waveforms, 17,170 have PPG waveforms, and 4,893 patients have both ABP and PPG waveforms. A more detailed description of the MLORD dataset can be found in^47^.

For the real-time evaluation, we ran our feature extraction tool on ABP and PPG waveforms extracted from a bedside Philips IntelliVue MX800 patient monitor in demonstration mode. The pipeline for real-time feature extraction consists of three main steps: (1) Transmitting PPG and/or ABP waveform data from the patient monitor to a standalone device (laptop) in real-time using open-access C# code (available at https://github.com/xeonfusion/VSCaptureMP), (2) Loading the live PPG and/or ABP waveforms into the feature extraction tool, and (3) Identifying landmarks within each cardiac cycle and extracting 852 features per cycle based on these landmarks. The real-time PPG/ABP waveform feature extraction pipeline is illustrated in Figure 6.

**Figure 6.**
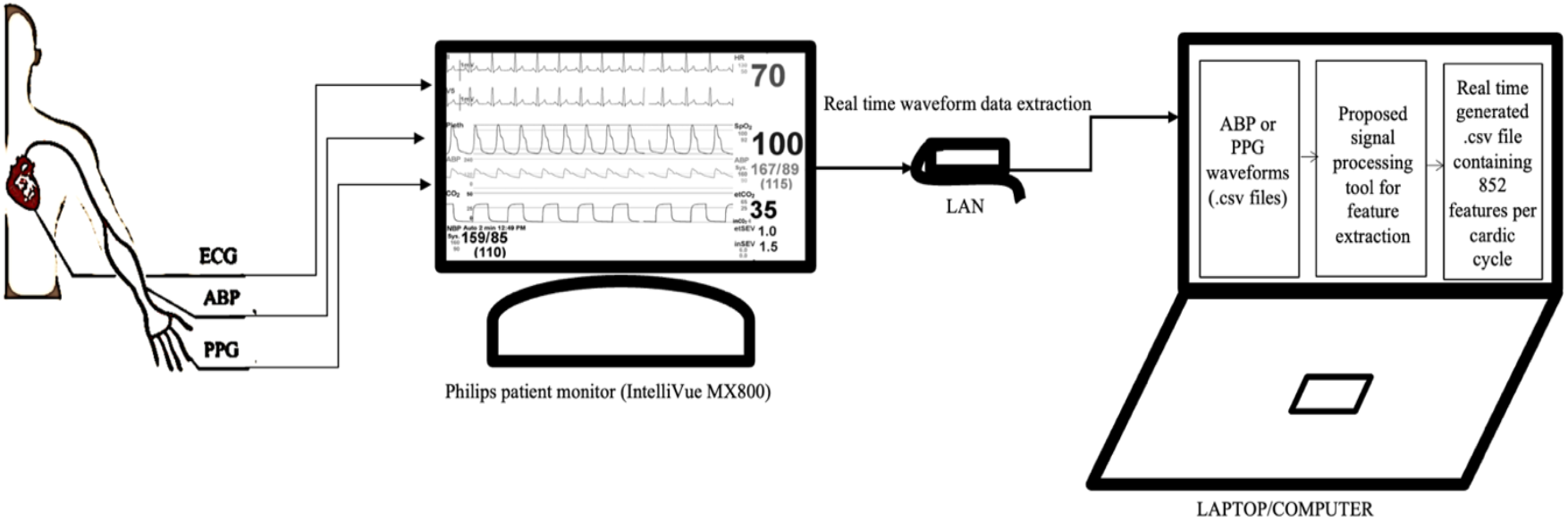
Real-time ABP/PPG waveform featurization pipeline using the feature extraction tool.

### Dataset markup for reference purposes

Due to the size of the MLORD dataset, manually marking landmarks within every cardiac cycle as a reference for evaluating the landmark detection performance of the tool was impractical. Additionally, in some cases—particularly with PPG waveforms— the dicrotic notch or diastolic phase peak is less distinct, making it challenging to identify their temporal location by inspection. Therefore, in our analysis, we randomly selected 10,000 4-s windows from ABP waveforms and the same number from PPG waveforms where all landmarks could be observed within a cardiac cycle. The choice of a 4-s window was pragmatic, based on ensuring good resolution of the waveform in the plots on the GUI (see figure 5) combined with ensuring a sufficient number of cardiac cycles per window (typically 4/5) for analysis. To maintain diversity and representativeness from the MLORD dataset, no more than 10 windows were selected per patient. The selected ABP windows include 34,267 cardiac cycles, and the PPG windows encompass 33,792 cardiac cycles. For the real-time evaluation, we retrospectively analyzed 3,000 cardiac cycles of ABP waveforms and 3,000 cardiac cycles of PPG waveforms collected from a bedside monitor (Philips IntelliVue MX800 patient monitor in demonstration mode). The protocol for evaluation is illustrated in figures 2(ii) and 2(iii). An experienced researcher visually marked the temporal location of all landmarks in these cardiac cycles, aided by the ‘find_peaks’ function from the SciPy Python library. These markings were then reviewed for accuracy by an engineer and an anesthesiologist, who conducted the review together. If they agreed that any 4-s window required adjustment, the researcher updated the markings accordingly. These visually marked windows were used as the reference for evaluating the performance of the feature extraction tool in detecting the temporal location of landmarks within the cardiac cycle. An example of an ABP signal and a PPG signal with visual mark-up is shown in Figure 3(i-a) and Figure 3(ii-a), respectively.

### Description of performance metrics

The landmark detection performance of the tool is assessed using four evaluative measures: sensitivity (SE), positive predictive value (PPV), F1-score, and error rate^32^. SE, defined as the probability of detecting true landmarks (as visually marked), is given by Eq. (2). The PPV, represented by Eq. (3), measures the proportion of detected landmarks that are true positives. The F1-score, calculated using Eq. (4), combines sensitivity and PPV to provide a comprehensive evaluation of the tool’s performance. Additionally, the tool’s performance is measured by the error rate, as specified in Eq. (5). Here, TP denotes true positives, FN stands for false negatives, and FP represents false positives. Additionally, Bland-Altman plots were utilized to assess the level of agreement between the landmarks marked by the researcher (the reference) and those detected by the tool.

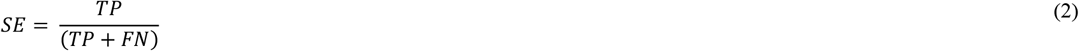

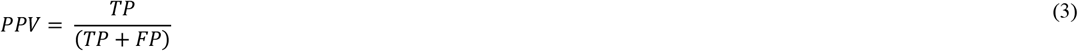

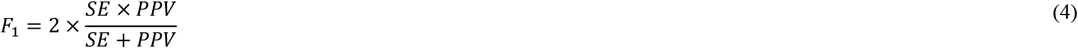

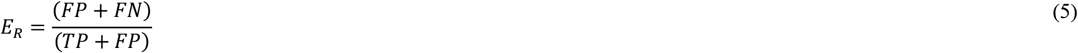

Note that a landmark is classified as a TP if it falls within ±8 ms (equivalent to 2 samples) of the reference mark; otherwise, it is considered a FP^32^. A FN is recorded when the tool fails to detect a landmark that should have been identified.

## Results

The performance of the feature extraction tool for detecting the temporal location of landmarks within a cardiac cycle in ABP and PPG waveforms is summarized in Table 3. This table reports the average SE, PPV, F1-score, and error rate for each landmark, with results presented as mean and standard deviation expressed as a percentage.

**Table 3.** Average evaluation measures of the performance of the feature extraction tool for detection of the temporal location of landmarks in the cardiac cycle of ABP or PPG waveforms on MLORD dataset and Real-time data (retrospective analysis). Results are reported as mean (standard deviation) % for Sensitivity (SE), Positive Predictive Value (PPV), F1-score and Error Rate (ER).

| Dataset | Waveform | Number of cardiac cycles | Landmark | SE (%) | PPV (%) | F<sub>1</sub> (%) | ER (%) |
| --- | --- | --- | --- | --- | --- | --- | --- |
| MLORD dataset | ABP | n=34,267 | SPO | 100 (0) | 99.65 (3.96) | 99.77 (2.73) | 0.35 (3.96) |
|  |  |  | SPP | 100 (0) | 99.70 (3.77) | 99.80 (2.63) | 0.30 (3.77) |
|  |  |  | DN | 100 (0) | 97.37 (11.01) | 98.24 (7.80) | 2.63 (11.01) |
|  |  |  | DPP | 100 (0) | 97.89 (9.87) | 98.59 (6.97) | 2.11 (9.87) |
|  | PPG | n=33,792 | SPO | 100 (0) | 99.25 (5.50) | 99.52 (3.64) | 0.75 (5.50) |
|  |  |  | SPP | 100 (0) | 99.30 (5.32) | 99.56 (3.54) | 0.70 (5.32) |
|  |  |  | DN | 100 (0) | 98.04 (9.12) | 98.72 (6.24) | 1.96 (9.12) |
|  |  |  | DPP | 100 (0) | 98.27 (8.56) | 98.88 (5.85) | 1.73 (8.56) |
| Real-time data | ABP | n=3,000 | SPO | 100 (0) | 96.97 (9.59) | 98.18 (5.75) | 3.03 (9.59) |
|  |  |  | SPP | 100 (0) | 97.03 (9.50) | 98.22 (5.70) | 2.97 (9.50) |
|  |  |  | DN | 100 (0) | 96.20 (10.60) | 97.72 (6.36) | 3.80 (10.60) |
|  |  |  | DPP | 100 (0) | 96.73 (9.91) | 98.04 (5.95) | 3.27 (9.91) |
|  | PPG | n=3,000 | SPO | 100 (0) | 96.57 (10.14) | 97.94 (6.08) | 3.43 (10.14) |
|  |  |  | SPP | 100 (0) | 96.23 (10.56) | 97.74 (6.34) | 3.77 (10.56) |
|  |  |  | DN | 100 (0) | 96.93 (9.64) | 98.16 (5.78) | 3.07 (9.64) |
|  |  |  | DPP | 100 (0) | 96.80 (9.82) | 98.08 (5.89) | 3.20 (9.82) |

ABP: Arterial blood pressure; PPG: Photoplethysmography; SPO: Systolic phase onset; SPP: Systolic phase peak; DN: Dicrotic notch; DPP: Diastolic phase peak; DPE: Diastolic phase endpoint;

### Landmarks detection in arterial blood pressure and photoplethysmography waveforms on the MLORD dataset

On average, the feature extraction tool achieved a SE of 100 %, a PPV exceeding 97%, and an F1-score above 98 % for detecting the temporal location of landmarks in both ABP and PPG waveforms in the MLORD dataset. The average error rate for detecting the temporal location of landmarks in both waveform types was consistently below 3 %. Additionally, examples in Figure 5(a) for PPG and Figure 5(b) for ABP waveforms demonstrate the tool’s ability to accurately identify the temporal location of landmarks within the cardiac cycle, despite morphological variations.

Bland-Altman plots comparing the agreement between the locations of landmarks found by the feature extraction tool and the reference locations (figure 7) show a mean bias close to zero with narrow limits of agreement (±0.01) for all landmarks in both ABP (Figure 7(i)) and PPG (Figure 7(ii)) waveforms. The plots, labeled (a) through (d), compare the following landmarks: (a) systolic phase onset, (b) systolic phase peak, (c) dicrotic notch, and (d) diastolic phase peak. A high level of agreement between the reference landmarks, as marked visually, and those detected by the feature extraction tool across both ABP and PPG waveforms is indicated.

**Figure 7.**
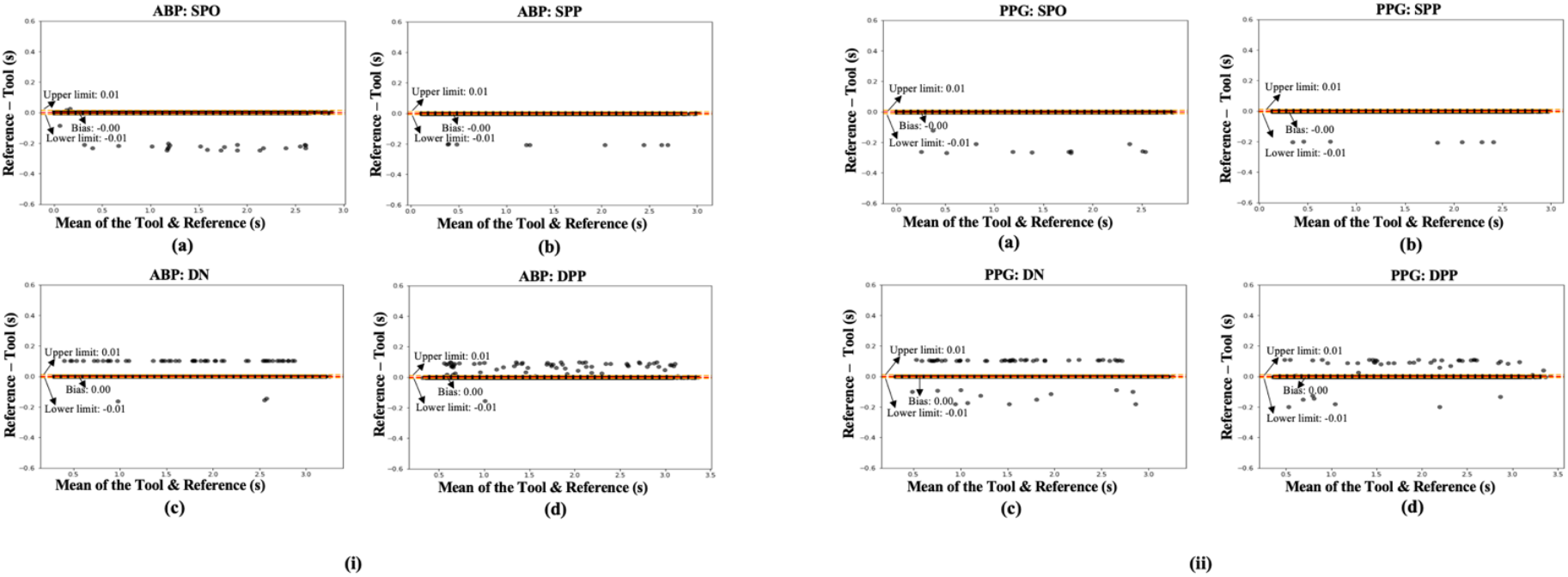
Bland Altman plots showing the agreement between the reference (visually marked) location and the location of landmarks identified by the feature extraction tool in both (i) arterial blood pressure and (ii) photoplethysmography waveforms on the MLORD dataset. The plots labeled (a) through (e) show comparisons for: (a) systolic phase onset, (b) systolic phase peak, (c) dicrotic notch, and (d) diastolic phase peak. For all plots, including (i) (a-e) and (ii) (a-e), the number of cardiac cycles are n = 33,000.

### Real time landmarks detection in arterial blood pressure and photoplethysmography waveforms

One of the key motivations for this study was to develop a feature extraction tool that not only works with recorded datasets but can also be applied to real-time data for analyzing ABP and PPG waveforms. When detecting landmark’s in real-time in both ABP and PPG waveforms, the tool achieved, on average, a sensitivity (SE) of 100 %, a positive predictive value (PPV) greater than 96 %, an F1-score above 97 %, and an error rate below 4 % (See Table 3).

Moreover, the Bland-Altman plots comparing the reference landmarks to those detected by the feature extraction tool in real time (figure 8), show a mean bias close to zero, with narrow limits of agreement ((±0.02) for all landmarks in both ABP and PPG waveforms. The plots, labeled from (a) to (d), compare the following landmarks : (a) systolic phase onset, (b) systolic phase peak, (c)dicrotic notch, and (d) diastolic phase peak. These results show strong agreement between the reference landmarks, visually marked by the researcher, and the landmarks detected in real-time by the feature extraction tool in both ABP and PPG waveforms.

**Figure 8.**
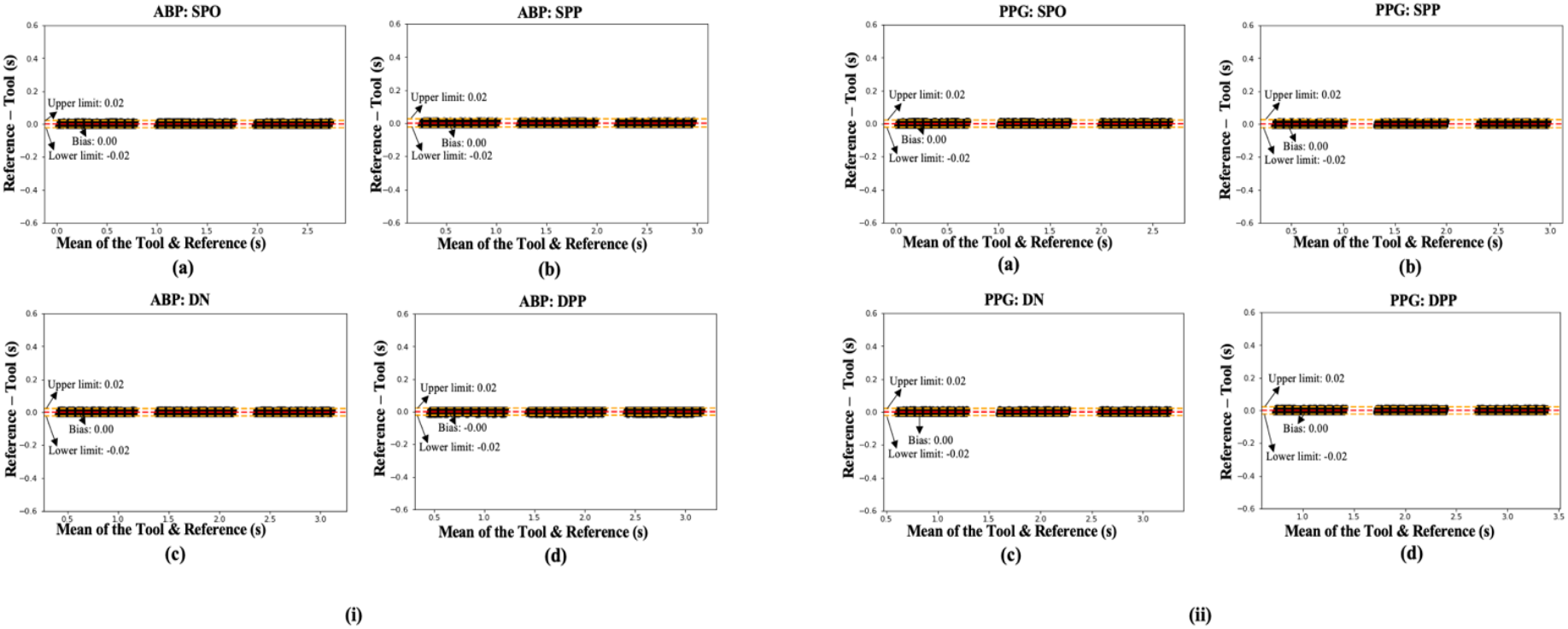
Bland Altman plots showing the agreement between the reference (visually marked) location and the location of landmarks identified by the feature extraction tool in real-time in both (i) arterial blood pressure and (ii) photoplethysmography waveforms on the data from the Philips IntelliVue MX800 bedside patient monitor (retrospective analysis). The plots labeled (a) through (e) show comparisons for: (a) systolic phase onset, (b) systolic phase peak, (c) dicrotic notch, and (d) diastolic phase peak. For all plots, including (i) (a-e) and (ii) (a-e), the number of cardiac cycles are n = 3000.

## Discussion

In this paper, we introduce a novel feature extraction tool designed to extract features from both ABP and PPG waveforms based on the detected landmarks within a cardiac cycle. Our tool is capable of deriving a wide range of features per cardiac cycle, including time-domain, statistical, and frequency-domain features (see supplementary material for the detailed description of the features). Most of the features extracted by the tool, whether related to ABP or PPG waveform, are either already in clinical use ^9,21,24,50,51,52^ or hold potential for clinical application. The feature extraction tool offers a comprehensive set of features that can be valuable for training machine learning models in specific diagnosis and risk-prediction tasks. Moreover, it can play a crucial role in mobile health or wearable devices, especially those based on the non-invasive PPG waveform, where the extracted features can be essential for tracking health and fitness in everyday life ^53,54^.

A key aspect of effective feature extraction is the precise identification of temporal locations of landmarks within each cardiac cycle, as these points form the foundation for extracting clinically useful features from ABP and PPG waveforms. To address this, our feature extraction tool operates in two primary stages: first, it locates the temporal location of landmarks within the ABP/PPG waveforms; second, it extracts relevant features based on these detected landmarks. The primary advantage of our tool is its ability to analyze both ABP and PPG waveforms in recorded and real-time modes, whereas most existing methods in the literature are developed exclusively for either ABP or PPG waveform analysis (as shown in Table 1), or have not been tested in real-time. Additionally, the IEM method used in our tool to prepare the signal for detecting landmarks within a cardiac cycle is computationally efficient, with a complexity of *O(JN)* as described in Big-O notation^55^, where *J* represent the number of iterations and *N* denotes the signal length^35^.

We have developed a GUI for the feature extraction tool that supports both functionalities: recorded datasets and real-time waveform analysis. The GUI of our tool significantly enhances both usability and accessibility. Designed to be intuitive and user-friendly, it facilitates seamless interaction with the tool. The GUI offers visualization of the waveforms and detected landmarks, aiding in the precise verification of results. This user-centric design not only enhances the tool’s effectiveness but also simplifies its potential integration into clinical and research settings. The improved interface ensures that users can easily interact with the tool, analyze waveforms in detail, and extract their morphological features, ultimately supporting better analysis of waveforms for clinical applications or research.

Our study has several limitations: first, while the feature extraction tool performs well on a large dataset (MLORD^47^) and shows resilience to noise under typical recording conditions, it has not been specifically tested for high-frequency artifacts/noise. Incorporating PPG or ABP denoising techniques^56,57^ as an additional pre-processing step could enhance its performance in such scenarios. Second, during the pre-processing step, we used fixed criteria to identify and remove artifact-affected windows. This approach may unintentionally exclude valid windows that do not meet these predetermined standards. To address this, future studies could benefit from an adaptive approach that refines artifact detection criteria. Additionally, incorporating automated classification methods, as explored in^58,59^, could improve the differentiation between clean and artifact-affected windows, ultimately making the feature extraction tool more robust and effective in waveform analysis. Third, the fixed SG filter parameter (i.e., the number of coefficients) used in the IEM method can pose challenges, particularly in the presence of high-frequency background noise. This can lead to inaccurate envelope mean estimations and hinder the detection of landmarks in non-stationary outputs. Additionally, the IEM method’s reliance on a fixed accuracy level (*β*) for its stopping criteria may further affect landmark identification. Future studies could explore automated approaches for selecting the parameters used in the IEM method based on the characteristics of the data, which could help address these issues. Fourth, we used simulated waveforms from the demonstration mode of the Philips IntelliVue MX800 bedside patient monitor to test the tool’s ability to detect landmarks within a cardiac cycle of ABP and PPG waveforms and to featurize in real-time. Future studies should focus on evaluating the tool’s performance in real-time with real patient data (operational mode of the patient monitor) to assess its practical utility in clinical settings. Fifth, in this study, we evaluated the feature extraction tool’s ability to detect landmarks within a cardiac cycle of ABP and PPG waveforms. However, the clinical usability of the features the tool can extract has not been explicitly explored here.

## Conclusions

This study presents an automated feature extraction tool which significantly advances the analysis of the physiological ABP and PPG waveforms. Using the IEM method, the tool first detects the temporal location of landmarks within each cardiac cycle with minimal computational cost. It then extracts 852 features per cardiac cycle from the ABP or PPG waveforms based on these detected landmarks and stores them in a .csv file. These extracted features can be integrated into machine learning models for healthcare applications, paving the way for innovative advancements in patient monitoring and diagnosis. The feature extraction tool has demonstrated high performance for detecting the temporal location of landmarks in both ABP and PPG waveforms when evaluated using the large perioperative MLORD dataset and real-time data, achieving an average F1-score exceeding 97 % and an average error rate of less than 4 %. Additionally, the user-friendly GUI of our tool provides researchers and healthcare professionals with a valuable resource for in-depth analysis of ABP or PPG waveforms. Furthermore, this combination of high performance, usability, and integration potential makes our tool an attractive and powerful asset for advancing healthcare applications. Future research will focus on evaluating the performance of our tool for detecting landmarks within a cardiac cycle of ABP and PPG waveforms across diverse datasets from various clinical settings, as well as conducting real-time assessments with patients to evaluate its generalizability and real-world applicability. In parallel, we will also investigate the clinical relevance and usefulness of the features extracted by our tool.

## Data Availability

The interested parties may reach out to the first author at or the corresponding author at to request access to the MLORD dataset and the proposed ABP/PPG waveform featurization tool.

## Acknowledgements

This work was supported by the National Institutes of Health (NIH): R01EB029751 and R01HL144692.

## Author contributions statement

R.P. contributed to study design, data analysis, conceptualization, and manuscript preparation. A.R., T.W., and J.N.C. contributed to study design and review & editing of the manuscript. A.B. contributed to study design, supervision of the study, review & editing of the manuscript, and conceptualization. M.C. contributed to study design, supervision of the study, review & editing of the manuscript, funding acquisition, and conceptualization. All authors have read and approved the final version of the manuscript.

## Declaration of competing interest

Dr. Cannesson is a consultant for Edwards Lifesciences and Masimo Corp, and has funded research from Edwards Lifesciences and Masimo Corp. He is also the founder of Sironis and Perceptive Medical and he owns patents and receives royalties for closed loop hemodynamic management technologies that have been licensed to Edwards Lifesciences.

## Notes

### Author Declarations

Institutional Review Board of University of California, Los Angeles gave ethical approval for this work.

