## Supplementary material for "Feature Extraction Tool Using Temporal Landmarks in Arterial Blood Pressure and Photoplethysmography Waveforms": Supplementary_material.docx

**Detailed descriptions of the features (n1=852) that the feature extraction tool can extract from arterial blood pressure (ABP) and photoplethysmography (PPG) waveforms per cardiac cycle**

1. **Original and normalized amplitude features (1-30)**

| 1 | ABP/PPG_AM_SPO_wrtZero | ABP/PPG cardiac cycle systolic phase onset amplitude with respect to zero |
| --- | --- | --- |
| 2 | ABP/PPG_AM_SPP_wrtZero | ABP/PPG cardiac cycle systolic phase peak amplitude with respect to zero |
| 3 | ABP/PPG_AM_DN_wrtZero | ABP/PPG cardiac cycle dicrotic notch amplitude with respect to zero |
| 4 | ABP/PPG_AM_DPP_wrtZero | ABP/PPG cardiac cycle of diastolic phase peak amplitude with respect to zero |
| 5 | ABP/PPG_AM_DPE_wrtZero | ABP/PPG cardiac cycle diastolic phase end point amplitude with respect to zero |
| 6 | ABP/PPG_NAM_SPO_wrtZero | Normalized ABP/PPG cardiac cycle systolic phase onset amplitude with respect to zero |
| 7 | ABP/PPG_NAM_SPP_wrtZero | Normalized ABP/PPG cardiac cycle systolic phase peak amplitude with respect to zero |
| 8 | ABP/PPG_NAM_DN_wrtZero | Normalized ABP/PPG cardiac cycle dicrotic notch amplitude with respect to zero |
| 9 | ABP/PPG_NAM_DPP_wrtZero | Normalized ABP/PPG cardiac cycle diastolic phase peak amplitude with respect to zero |
| 10 | ABP/PPG_NAM_DPE_wrtZero | Normalized ABP/PPG cardiac cycle diastolic phase end point amplitude with respect to zero |
| 11 | ABP/PPG_AM_SPP_wrtSPO | ABP/PPG cardiac cycle systolic phase peak amplitude with respect to systolic phase onset |
| 12 | ABP/PPG_ AM_ DN_wrtSPO | ABP/PPG cardiac cycle dicrotic notch amplitude with respect to systolic phase onset |
| 13 | ABP/PPG_ AM_ DPP_wrtSPO | ABP/PPG cardiac cycle diastolic phase peak amplitude with respect to systolic phase onset |
| 14 | ABP/PPG_ AM_ DPE_wrtSPO | ABP/PPG cardiac cycle diastolic phase end point amplitude with respect to systolic phase onset |
| 15 | ABP/PPG_ AM_ SPP_wrtDN | ABP/PPG cardiac cycle systolic phase peak amplitude with respect to dicrotic notch |
| 16 | ABP/PPG_ AM_ SPP_wrtDPP | ABP/PPG cardiac cycle systolic phase peak amplitude with respect to diastolic phase peak |
| 17 | ABP/PPG_ AM_ SPP_wrtDPE | ABP/PPG cardiac cycle systolic phase peak amplitude with respect to diastolic phase end point |
| 18 | ABP/PPG_ AM_ DN_wrt DPE | ABP/PPG cardiac cycle dicrotic notch amplitude with respect to diastolic phase end point |
| 19 | ABP/PPG_ AM_ DPP_wrtDN | ABP/PPG cardiac cycle diastolic phase peak amplitude with respect to dicrotic notch |
| 20 | ABP/PPG_ AM_ DPP_wrtDPE | ABP/PPG cardiac cycle diastolic phase peak amplitude with respect to diastolic phase end point |
| 21 | ABP/PPG_NAM_SPP_wrtSPO | Normalized ABP/PPG cardiac cycle systolic phase peak amplitude with respect to systolic phase onset |
| 22 | ABP/PPG_NAM_DN_wrtSPO | Normalized ABP/PPG cardiac cycle dicrotic notch amplitude with respect to systolic phase onset |
| 23 | ABP/PPG_NAM_DPP_wrtSPO | Normalized ABP/PPG cardiac cycle diastolic phase peak amplitude with respect to systolic phase onset |
| 24 | ABP/PPG_NAM_DPE_wrtSPO | Normalized ABP/PPG cardiac cycle diastolic phase end point amplitude with respect to systolic phase onset |
| 25 | ABP/PPG_NAM_SPP_wrtDN | Normalized ABP/PPG cardiac cycle systolic phase peak amplitude with respect to dicrotic notch |
| 26 | ABP/PPG_NAM_SPP_wrtDPP | Normalized ABP/PPG cardiac cycle systolic phase peak amplitude with respect to diastolic phase peak |
| 27 | ABP/PPG_NAM_SPP_wrtDPE | Normalized ABP/PPG cardiac cycle systolic phase peak amplitude with respect to diastolic phase end point |
| 28 | ABP/PPG_NAM_DN_wrtDPE | Normalized ABP/PPG cardiac cycle dicrotic notch amplitude with respect to diastolic phase end point |
| 29 | ABP/PPG_NAM_DPP_wrtDN | Normalized ABP/PPG cardiac cycle diastolic phase peak amplitude with respect to dicrotic notch |
| 30 | ABP/PPG_NAM_DPP_wrtDPE | Normalized ABP/PPG cardiac cycle diastolic phase peak amplitude with respect to diastolic phase end point |

2. **Amplitude ratio features (31-135)**

| 31 | ABP/PPG_AM_RT_SPP_wrtZero_ABP/PPG_SPO_wrtZero | The ratio of the ABP/PPG cardiac cycle systolic phase peak amplitude with respect to zero to the ABP/PPG cardiac cycle systolic phase onset amplitude with respect to zero |
| --- | --- | --- |
| 32 | ABP/PPG_AM_RT_SPP_wrtSPO_ABP/PPG_SPO_wrtZero | The ratio of the ABP/PPG cardiac cycle systolic phase peak amplitude with respect to systolic phase onset to the ABP/PPG cardiac cycle systolic phase onset amplitude with respect to zero |
| 33 | ABP/PPG_AM_RT_SPP_wrtDN_ABP/PPG_SPO_wrtZero | The ratio of the ABP/PPG cardiac cycle systolic phase peak amplitude with respect to dicrotic notch to the ABP/PPG cardiac cycle systolic phase onset amplitude with respect to zero |
| 34 | ABP/PPG_AM_RT_SPP_wrtDPP_ABP/PPG_SPO_wrtZero | The ratio of the ABP/PPG cardiac cycle systolic phase peak amplitude with respect to diastolic phase peak to the ABP/PPG cardiac cycle systolic phase onset amplitude with respect to zero |
| 35 | ABP/PPG_AM_RT_SPP_wrtDPE_ABP/PPG_SPO_wrtZero | The ratio of the ABP/PPG cardiac cycle systolic phase peak amplitude with respect to diastolic phase end point to the ABP/PPG cardiac cycle systolic phase onset amplitude with respect to zero |
| 36 | ABP/PPG_AM_RT_DN_wrtZero_ABP/PPG_SPO_wrtZero | The ratio of the ABP/PPG cardiac cycle dicrotic notch amplitude with respect to zero to the ABP/PPG cardiac cycle systolic phase onset amplitude with respect to zero |
| 37 | ABP/PPG_AM_RT_DN_wrtSPO_ABP/PPG_SPO_wrtZero | The ratio of the ABP/PPG cardiac cycle dicrotic notch amplitude with respect to systolic phase onset to the ABP/PPG cardiac cycle systolic phase onset amplitude with respect to zero |
| 38 | ABP/PPG_AM_RT_DN_wrtDPE_ABP/PPG_SPO_wrtZero | The ratio of the ABP/PPG cardiac cycle dicrotic notch amplitude with respect to diastolic phase end point to the ABP/PPG cardiac cycle systolic phase onset amplitude with respect to zero |
| 39 | ABP/PPG_AM_RT_DPP_wrtZero_ABP/PPG_SPO_wrtZero | The ratio of the ABP/PPG cardiac cycle diastolic phase peak amplitude with respect to zero to the ABP/PPG cardiac cycle systolic phase onset amplitude with respect to zero |
| 40 | ABP/PPG_AM_RT_DPP_wrtSPO_ABP/PPG_SPO_wrtZero | The ratio of the ABP/PPG cardiac cycle diastolic phase peak amplitude with respect to systolic phase onset to the ABP/PPG cardiac cycle systolic phase onset amplitude with respect to zero |
| 41 | ABP/PPG_AM_RT_DPP_wrtDN_ABP/PPG_SPO_wrtZero | The ratio of the ABP/PPG cardiac cycle diastolic phase peak amplitude with respect to dicrotic notch to the ABP/PPG cardiac cycle systolic phase onset amplitude with respect to zero |
| 42 | ABP/PPG_AM_RT_DPP_wrtDPE_ABP/PPG_SPO_wrtZero | The ratio of the ABP/PPG cardiac cycle diastolic phase peak amplitude with respect to diastolic phase end point to the ABP/PPG cardiac cycle systolic phase onset amplitude with respect to zero |
| 43 | ABP/PPG_AM_RT_DPE_wrtZero_ABP/PPG_SPO_wrtZero | The ratio of the ABP/PPG cardiac cycle diastolic phase end point amplitude with respect to zero to the ABP/PPG cardiac cycle systolic phase onset amplitude with respect to zero |
| 44 | ABP/PPG_AM_RT_DPE_wrtSPO_ABP/PPG_SPO_wrtZero | The ratio of the ABP/PPG cardiac cycle diastolic phase end point amplitude with respect to systolic phase onset to the ABP/PPG cardiac cycle systolic phase onset amplitude with respect to zero |
| 45 | ABP/PPG_AM_RT_DN_wrtZero_ABP/PPG_SPP_wrtZero | The ratio of the ABP/PPG cardiac cycle dicrotic notch amplitude with respect to zero to the ABP/PPG cardiac cycle systolic phase peak amplitude with respect to zero |
| 46 | ABP/PPG_AM_RT_DN_wrtSPO_ABP/PPG_SPP_wrtZero | The ratio of the ABP/PPG cardiac cycle dicrotic notch amplitude with respect to systolic phase onset to the ABP/PPG cardiac cycle systolic phase peak amplitude with respect to zero |
| 47 | ABP/PPG_AM_RT_DN_wrtDPE_ABP/PPG_SPP_wrtZero | The ratio of the ABP/PPG cardiac cycle dicrotic notch amplitude with respect to diastolic phase end point to the ABP/PPG cardiac cycle systolic phase peak amplitude with respect to zero |
| 48 | ABP/PPG_AM_RT_DPP_wrtZero_ABP/PPG_SPP_wrtZero | The ratio of the ABP/PPG cardiac cycle diastolic phase peak amplitude with respect to zero to the ABP/PPG cardiac cycle systolic phase peak amplitude with respect to zero |
| 49 | ABP/PPG_AM_RT_DPP_wrtSPO_ABP/PPG_SPP_wrtZero | The ratio of the ABP/PPG cardiac cycle diastolic phase peak amplitude with respect to systolic phase onset to the ABP/PPG cardiac cycle systolic phase peak amplitude with respect to zero |
| 50 | ABP/PPG_AM_RT_DPP_wrtDN_ABP/PPG_SPP_wrtZero | The ratio of the ABP/PPG cardiac cycle diastolic phase peak amplitude with respect to dicrotic notch to the ABP/PPG cardiac cycle systolic phase peak amplitude with respect to zero |
| 51 | ABP/PPG_AM_RT_DPP_wrtDPE_ABP/PPG_SPP_wrtZero | The ratio of the ABP/PPG cardiac cycle diastolic phase peak amplitude with respect to diastolic phase end point to the ABP/PPG cardiac cycle systolic phase peak amplitude with respect to zero |
| 52 | ABP/PPG_AM_RT_DPE_wrtZero_ABP/PPG_SPP_wrtZero | The ratio of the ABP/PPG cardiac cycle diastolic phase end point amplitude with respect to zero to the ABP/PPG cardiac cycle systolic phase peak amplitude with respect to zero |
| 53 | ABP/PPG_AM_RT_DPE_wrtSPO_ABP/PPG_SPP_wrtZero | The ratio of the ABP/PPG cardiac cycle diastolic phase end point amplitude with respect to systolic phase onset to the ABP/PPG cardiac cycle systolic phase peak amplitude with respect to zero |
| 54 | ABP/PPG_AM_RT_DN_wrtZero_ABP/PPG_SPP_wrtSPO | The ratio of the ABP/PPG cardiac cycle dicrotic notch amplitude with respect to zero to the ABP/PPG cardiac cycle systolic phase peak amplitude with respect to systolic phase onset |
| 55 | ABP/PPG_AM_RT_DN_wrtSPO_ABP/PPG_SPP_wrtSPO | The ratio of the ABP/PPG cardiac cycle dicrotic notch amplitude with respect to systolic phase onset to the ABP/PPG cardiac cycle systolic phase peak amplitude with respect to systolic phase onset |
| 56 | ABP/PPG_AM_RT_DN_wrtDPE_ABP/PPG_SPP_wrtSPO | The ratio of the ABP/PPG cardiac cycle dicrotic notch amplitude with respect to diastolic phase end point to the ABP/PPG cardiac cycle systolic phase peak amplitude with respect to systolic phase onset |
| 57 | ABP/PPG_AM_RT_DPP_wrtZero_ABP/PPG_SPP_wrtSPO | The ratio of the ABP/PPG cardiac cycle diastolic phase peak amplitude with respect to zero to the ABP/PPG cardiac cycle systolic phase peak amplitude with respect to systolic phase onset |
| 58 | ABP/PPG_AM_RT_DPP_wrtSPO_ABP/PPG_SPP_wrtSPO | The ratio of the ABP/PPG cardiac cycle diastolic phase peak amplitude with respect to systolic phase onset to the ABP/PPG cardiac cycle systolic phase peak amplitude with respect to systolic phase onset |
| 59 | ABP/PPG_AM_RT_DPP_wrtDN_ABP/PPG_SPP_wrtSPO | The ratio of the ABP/PPG cardiac cycle diastolic phase peak amplitude with respect to dicrotic notch to the ABP/PPG cardiac cycle systolic phase peak amplitude with respect to systolic phase onset |
| 60 | ABP/PPG_AM_RT_DPP_wrtDPE_ABP/PPG_SPP_wrtSPO | The ratio of the ABP/PPG cardiac cycle diastolic phase peak amplitude with respect to diastolic phase end point to the ABP/PPG cardiac cycle systolic phase peak amplitude with respect to systolic phase onset |
| 61 | ABP/PPG_AM_RT_DPE_wrtZero_ABP/PPG_SPP_wrtSPO | The ratio of the ABP/PPG cardiac cycle diastolic phase end point amplitude with respect to zero to the ABP/PPG cardiac cycle systolic phase peak amplitude with respect to systolic phase onset |
| 62 | ABP/PPG_AM_RT_DPE_wrtSPO_ABP/PPG_SPP_wrtSPO | The ratio of the ABP/PPG cardiac cycle diastolic phase end point amplitude with respect to systolic phase onset to the ABP/PPG cardiac cycle systolic phase peak amplitude with respect to systolic phase onset |
| 63 | ABP/PPG_AM_RT_DN_wrtZero_ABP/PPG_SPP_wrtDN | The ratio of the ABP/PPG cardiac cycle dicrotic notch amplitude with respect to zero to the ABP/PPG cardiac cycle systolic phase peak amplitude with respect to dicrotic notch |
| 64 | ABP/PPG_AM_RT_DN_wrtSPO_ABP/PPG_SPP_wrtDN | The ratio of the ABP/PPG cardiac cycle dicrotic notch amplitude with respect to systolic phase onset to the ABP/PPG cardiac cycle systolic phase peak amplitude with respect to dicrotic notch |
| 65 | ABP/PPG_AM_RT_DN_wrtDPE_ABP/PPG_SPP_wrtDN | The ratio of the ABP/PPG cardiac cycle dicrotic notch amplitude with respect to diastolic phase end point to the ABP/PPG cardiac cycle systolic phase peak amplitude with respect to dicrotic notch |
| 66 | ABP/PPG_AM_RT_DPP_wrtZero_ABP/PPG_SPP_wrtDN | The ratio of the ABP/PPG cardiac cycle diastolic phase peak amplitude with respect to zero to the ABP/PPG cardiac cycle systolic phase peak amplitude with respect to dicrotic notch |
| 67 | ABP/PPG_AM_RT_DPP_wrtSPO_ABP/PPG_SPP_wrtDN | The ratio of the ABP/PPG cardiac cycle diastolic phase peak amplitude with respect to systolic phase onset to the ABP/PPG cardiac cycle systolic phase peak amplitude with respect to dicrotic notch |
| 68 | ABP/PPG_AM_RT_DPP_wrtDN_ABP/PPG_SPP_wrtDN | The ratio of the ABP/PPG cardiac cycle diastolic phase peak amplitude with respect to dicrotic notch to the ABP/PPG cardiac cycle systolic phase peak amplitude with respect to dicrotic notch |
| 69 | ABP/PPG_AM_RT_DPP_wrtDPE_ABP/PPG_SPP_wrtDN | The ratio of the ABP/PPG cardiac cycle diastolic phase peak amplitude with respect to diastolic phase end point to the ABP/PPG cardiac cycle systolic phase peak amplitude with respect to dicrotic notch |
| 70 | ABP/PPG_AM_RT_DPE_wrtZero_ABP/PPG_SPP_wrtDN | The ratio of the ABP/PPG cardiac cycle diastolic phase end point amplitude with respect to zero to the ABP/PPG cardiac cycle systolic phase peak amplitude with respect to dicrotic notch |
| 71 | ABP/PPG_AM_RT_DPE_wrtSPO_ABP/PPG_SPP_wrtDN | The ratio of the ABP/PPG cardiac cycle diastolic phase end point amplitude with respect to systolic phase onset to the ABP/PPG cardiac cycle systolic phase peak amplitude with respect to dicrotic notch |
| 72 | ABP/PPG_AM_RT_DN_wrtZero_ABP/PPG_SPP_wrtDPP | The ratio of the ABP/PPG cardiac cycle dicrotic notch amplitude with respect to zero to the ABP/PPG cardiac cycle systolic phase peak amplitude with respect to diastolic phase peak |
| 73 | ABP/PPG_AM_RT_DN_wrtSPO_ABP/PPG_SPP_wrtDPP | The ratio of the ABP/PPG cardiac cycle dicrotic notch amplitude with respect to systolic phase onset to the ABP/PPG cardiac cycle systolic phase peak amplitude with respect to diastolic phase peak |
| 74 | ABP/PPG_AM_RT_DN_wrtDPE_ABP/PPG_SPP_wrtDPP | The ratio of the ABP/PPG cardiac cycle dicrotic notch amplitude with respect to diastolic phase end point to the ABP/PPG cardiac cycle systolic phase peak amplitude with respect to diastolic phase peak |
| 75 | ABP/PPG_AM_RT_DPP_wrtZero_ABP/PPG_SPP_wrtDPP | The ratio of the ABP/PPG cardiac cycle diastolic phase peak amplitude with respect to zero to the ABP/PPG cardiac cycle systolic phase peak amplitude with respect to diastolic phase peak |
| 76 | ABP/PPG_AM_RT_DPP_wrtSPO_ABP/PPG_SPP_wrtDPP | The ratio of the ABP/PPG cardiac cycle diastolic phase peak amplitude with respect to systolic phase onset to the ABP/PPG cardiac cycle systolic phase peak amplitude with respect to diastolic phase peak |
| 77 | ABP/PPG_AM_RT_DPP_wrtDN_ABP/PPG_SPP_wrtDPP | The ratio of the ABP/PPG cardiac cycle diastolic phase peak amplitude with respect to dicrotic notch to the ABP/PPG cardiac cycle systolic phase peak amplitude with respect to diastolic phase peak |
| 78 | ABP/PPG_AM_RT_DPP_wrtDPE_ABP/PPG_SPP_wrtDPP | The ratio of the ABP/PPG cardiac cycle diastolic phase peak amplitude with respect to diastolic phase end point to the ABP/PPG cardiac cycle systolic phase peak amplitude with respect to diastolic phase peak |
| 79 | ABP/PPG_AM_RT_DPE_wrtZero_ABP/PPG_SPP_wrtDPP | The ratio of the ABP/PPG cardiac cycle diastolic phase end point amplitude with respect to zero to the ABP/PPG cardiac cycle systolic phase peak amplitude with respect to diastolic phase peak |
| 80 | ABP/PPG_AM_RT_DPE_wrtSPO_ABP/PPG_SPP_wrtDPP | The ratio of the ABP/PPG cardiac cycle diastolic phase end point amplitude with respect to systolic phase onset to the ABP/PPG cardiac cycle systolic phase peak amplitude with respect to diastolic phase peak |
| 81 | ABP/PPG_AM_RT_DN_wrtZero_ABP/PPG_SPP_wrtDPE | The ratio of the ABP/PPG cardiac cycle dicrotic notch amplitude with respect to zero to the ABP/PPG cardiac cycle systolic phase peak amplitude with respect to diastolic phase end point |
| 82 | ABP/PPG_AM_RT_DN_wrtSPO_ABP/PPG_SPP_wrtDPE | The ratio of the ABP/PPG cardiac cycle dicrotic notch amplitude with respect to systolic phase onset to the ABP/PPG cardiac cycle systolic phase peak amplitude with respect to diastolic phase end point |
| 83 | ABP/PPG_AM_RT_DN_wrtDPE_ABP/PPG_SPP_wrtDPE | The ratio of the ABP/PPG cardiac cycle dicrotic notch amplitude with respect to diastolic phase end point to the ABP/PPG cardiac cycle systolic phase peak amplitude with respect to diastolic phase end point |
| 84 | ABP/PPG_AM_RT_DPP_wrtZero_ABP/PPG_SPP_wrtDPE | The ratio of the ABP/PPG cardiac cycle diastolic phase peak amplitude with respect to zero to the ABP/PPG cardiac cycle systolic phase peak amplitude with respect to diastolic phase end point |
| 85 | ABP/PPG_AM_RT_DPP_wrtSPO_ABP/PPG_SPP_wrtDPE | The ratio of the ABP/PPG cardiac cycle diastolic phase peak amplitude with respect to systolic phase onset to the ABP/PPG cardiac cycle systolic phase peak amplitude with respect to diastolic phase end point |
| 86 | ABP/PPG_AM_RT_DPP_wrtDN_ABP/PPG_SPP_wrtDPE | The ratio of the ABP/PPG cardiac cycle diastolic phase peak amplitude with respect to dicrotic notch to the ABP/PPG cardiac cycle systolic phase peak amplitude with respect to diastolic phase end point |
| 87 | ABP/PPG_AM_RT_DPP_wrtDPE_ABP/PPG_SPP_wrtDPE | The ratio of the ABP/PPG cardiac cycle diastolic phase peak amplitude with respect to diastolic phase end point to the ABP/PPG cardiac cycle systolic phase peak amplitude with respect to diastolic phase end point |
| 88 | ABP/PPG_AM_RT_DPE_wrtZero_ABP/PPG_SPP_wrtDPE | The ratio of the ABP/PPG cardiac cycle diastolic phase end point amplitude with respect to zero to the ABP/PPG cardiac cycle systolic phase peak amplitude with respect to diastolic phase end point |
| 89 | ABP/PPG_AM_RT_DPE_wrtSPO_ABP/PPG_SPP_wrtDPE | The ratio of the ABP/PPG cardiac cycle diastolic phase end point amplitude with respect to systolic phase peak to the ABP/PPG cardiac cycle systolic phase peak amplitude with respect to diastolic phase end point |
| 90 | ABP/PPG_AM_RT_DPP_wrtZero_ABP/PPG_DN_wrtZero | The ratio of the ABP/PPG cardiac cycle diastolic phase peak amplitude with respect to zero to the ABP/PPG cardiac cycle dicrotic notch amplitude with respect to zero |
| 91 | ABP/PPG_AM_RT_DPP_wrtSPO_ABP/PPG_DN_wrtZero | The ratio of the ABP/PPG cardiac cycle diastolic phase peak amplitude with respect to systolic phase onset to the ABP/PPG cardiac cycle dicrotic notch amplitude with respect to zero |
| 92 | ABP/PPG_AM_RT_DPP_wrtDN_ABP/PPG_DN_wrtZero | The ratio of the ABP/PPG cardiac cycle diastolic phase peak amplitude with respect to dicrotic notch to the ABP/PPG cardiac cycle dicrotic notch amplitude with respect to zero |
| 93 | ABP/PPG_AM_RT_DPP_wrtDPE_ABP/PPG_DN_wrtZero | The ratio of the ABP/PPG cardiac cycle diastolic phase peak amplitude with respect to diastolic phase end point to the ABP/PPG cardiac cycle dicrotic notch amplitude with respect to zero |
| 94 | ABP/PPG_AM_RT_DPE_wrtZero_ABP/PPG_DN_wrtZero | The ratio of the ABP/PPG cardiac cycle diastolic phase end point amplitude with respect to zero to the ABP/PPG cardiac cycle dicrotic notch amplitude with respect to zero |
| 95 | ABP/PPG_AM_RT_DPE_wrtSPO_ABP/PPG_DN_wrtZero | The ratio of the ABP/PPG cardiac cycle diastolic phase end point amplitude with respect to systolic phase onset to the ABP/PPG cardiac cycle dicrotic notch amplitude with respect to zero |
| 96 | ABP/PPG_AM_RT_DPP_wrtZero_ABP/PPG_DN_wrtSPO | The ratio of the ABP/PPG cardiac cycle diastolic phase peak amplitude with respect to zero to the ABP/PPG cardiac cycle dicrotic notch amplitude with respect to systolic phase onset |
| 97 | ABP/PPG_AM_RT_DPP_wrtSPO_ABP/PPG_DN_wrtSPO | The ratio of the ABP/PPG cardiac cycle diastolic phase peak amplitude with respect to systolic phase onset to the ABP/PPG cardiac cycle dicrotic notch amplitude with respect to systolic phase onset |
| 98 | ABP/PPG_AM_RT_DPP_wrtDN_ABP/PPG_DN_wrtSPO | The ratio of the ABP/PPG cardiac cycle diastolic phase peak amplitude with respect to dicrotic notch to the ABP/PPG cardiac cycle dicrotic notch amplitude with respect to systolic phase onset |
| 99 | ABP/PPG_AM_RT_DPP_wrtDPE_ABP/PPG_DN_wrtSPO | The ratio of the ABP/PPG cardiac cycle diastolic phase peak amplitude with respect to diastolic phase end point to the ABP/PPG cardiac cycle dicrotic notch amplitude with respect to systolic phase onset |
| 100 | ABP/PPG_AM_RT_DPE_wrtZero_ABP/PPG_DN_wrtSPO | The ratio of the ABP/PPG cardiac cycle diastolic phase end point amplitude with respect to zero to the ABP/PPG cardiac cycle dicrotic notch amplitude with respect to systolic phase onset |
| 101 | ABP/PPG_AM_RT_DPE_wrtSPO_ABP/PPG_DN_wrtSPO | The ratio of the ABP/PPG cardiac cycle diastolic phase end point amplitude with respect to systolic phase onset to the ABP/PPG cardiac cycle dicrotic notch amplitude with respect to systolic phase onset |
| 102 | ABP/PPG_AM_RT_DPP_wrtZero_ABP/PPG_DN_wrtDPE | The ratio of the ABP/PPG cardiac cycle diastolic phase peak amplitude with respect to zero to the ABP/PPG cardiac cycle dicrotic notch amplitude with respect to diastolic phase end point |
| 103 | ABP/PPG_AM_RT_DPP_wrtSPO_ABP/PPG_DN_wrtDPE | The ratio of the ABP/PPG cardiac cycle diastolic phase peak amplitude with respect to systolic phase onset to the ABP/PPG cardiac cycle dicrotic notch amplitude with respect to diastolic phase end point |
| 104 | ABP/PPG_AM_RT_DPP_wrtDN_ABP/PPG_DN_wrtDPE | The ratio of the ABP/PPG cardiac cycle diastolic phase peak amplitude with respect to dicrotic notch to the ABP/PPG cardiac cycle dicrotic notch amplitude with respect to diastolic phase end point |
| 105 | ABP/PPG_AM_RT_DPP_wrtDPE_ABP/PPG_DN_wrtDPE | The ratio of the ABP/PPG cardiac cycle diastolic phase peak amplitude with respect to diastolic phase end point to the ABP/PPG cardiac cycle dicrotic notch amplitude with respect to diastolic phase end point |
| 106 | ABP/PPG_AM_RT_DPE_wrtZero_ABP/PPG_DN_wrtDPE | The ratio of the ABP/PPG cardiac cycle diastolic phase end point amplitude with respect to zero to the ABP/PPG cardiac cycle dicrotic notch amplitude with respect to diastolic phase end point |
| 107 | ABP/PPG_AM_RT_DPE_wrtSPO_ABP/PPG_DN_wrtDPE | The ratio of the ABP/PPG cardiac cycle diastolic phase end point amplitude with respect to systolic phase onset to the ABP/PPG cardiac cycle dicrotic notch amplitude with respect to diastolic phase end point |
| 108 | ABP/PPG_AM_RT_DPE_wrtZero_ABP/PPG_DPP_wrtZero | The ratio of the ABP/PPG cardiac cycle diastolic phase end point amplitude with respect to zero to the ABP/PPG cardiac cycle diastolic phase peak amplitude with respect to zero |
| 109 | ABP/PPG_AM_RT_DPE_wrtSPO_ABP/PPG_DPP_wrtZero | The ratio of the ABP/PPG cardiac cycle diastolic phase end point amplitude with respect to systolic phase onset to the ABP/PPG cardiac cycle diastolic phase peak amplitude with respect to zero |
| 110 | ABP/PPG_AM_RT_DPE_wrtZero_ABP/PPG_DPP_wrtSPO | The ratio of the ABP/PPG cardiac cycle diastolic phase end point amplitude with respect to zero to the ABP/PPG cardiac cycle diastolic phase peak amplitude with respect to systolic phase onset |
| 111 | ABP/PPG_AM_RT_DPE_wrtSPO_ABP/PPG_DPP_wrtSPO | The ratio of the ABP/PPG cardiac cycle diastolic phase end point amplitude with respect to systolic phase onset to the ABP/PPG cardiac cycle diastolic phase peak amplitude with respect to systolic phase onset |
| 112 | ABP/PPG_AM_RT_DPE_wrtZero_ABP/PPG_DPP_wrtDN | The ratio of the ABP/PPG cardiac cycle diastolic phase end point amplitude with respect to zero to the ABP/PPG cardiac cycle diastolic phase peak amplitude with respect to dicrotic notch |
| 113 | ABP/PPG_AM_RT_DPE_wrtSPO_ABP/PPG_DPP_wrtDN | The ratio of the ABP/PPG cardiac cycle diastolic phase end point amplitude with respect to systolic phase onset to the ABP/PPG cardiac cycle diastolic phase peak amplitude with respect to dicrotic notch |
| 114 | ABP/PPG_AM_RT_DPE_wrtZero_ABP/PPG_DPP_wrtDPE | The ratio of the ABP/PPG cardiac cycle diastolic phase end point amplitude with respect to zero to the ABP/PPG cardiac cycle diastolic phase peak amplitude with respect to diastolic phase end point |
| 115 | ABP/PPG_AM_RT_DPE_wrtSPO_ABP/PPG_DPP_wrtDPE | The ratio of the ABP/PPG cardiac cycle diastolic phase end point amplitude with respect to systolic phase onset to the ABP/PPG cardiac cycle diastolic phase peak amplitude with respect to diastolic phase end point |
| 116 | ABP/PPG_AM_RT_SPP_wrtSPO_ABP/PPG_SPP_wrtZero | The ratio of the ABP/PPG cardiac cycle systolic phase peak amplitude with respect to systolic phase onset to the ABP/PPG cardiac cycle systolic phase peak amplitude with respect to zero |
| 117 | ABP/PPG_AM_RT_SPP_wrtDN_ABP/PPG_SPP_wrtZero | The ratio of the ABP/PPG cardiac cycle systolic phase peak amplitude with respect to dicrotic notch to the ABP/PPG cardiac cycle systolic phase peak amplitude with respect to zero |
| 118 | ABP/PPG_AM_RT_SPP_wrtDPP_ABP/PPG_SPP_wrtZero | The ratio of the ABP/PPG cardiac cycle systolic phase peak amplitude with respect to diastolic phase peak to the ABP/PPG cardiac cycle systolic phase peak amplitude with respect to zero |
| 119 | ABP/PPG_AM_RT_SPP_wrtDPE_ABP/PPG_SPP_wrtZero | The ratio of the ABP/PPG cardiac cycle systolic phase peak amplitude with respect to diastolic phase end point to the ABP/PPG cardiac cycle systolic phase peak amplitude with respect to zero |
| 120 | ABP/PPG_AM_RT_SPP_wrtDN_ABP/PPG_SPP_wrtSPO | The ratio of the ABP/PPG cardiac cycle systolic phase peak amplitude with respect to dicrotic notch to the ABP/PPG cardiac cycle systolic phase peak amplitude with respect to systolic phase onset |
| 121 | ABP/PPG_AM_RT_SPP_wrtDPP_ABP/PPG_SPP_wrtSPO | The ratio of the ABP/PPG cardiac cycle systolic phase peak amplitude with respect to diastolic phase peak to the ABP/PPG cardiac cycle systolic phase peak amplitude with respect to systolic phase onset |
| 122 | ABP/PPG_AM_RT_SPP_wrtDPE_ABP/PPG_SPP_wrtSPO | The ratio of the ABP/PPG cardiac cycle systolic phase peak amplitude with respect to diastolic phase end point to the ABP/PPG cardiac cycle systolic phase peak amplitude with respect to systolic phase onset |
| 123 | ABP/PPG_AM_RT_SPP_wrtDPP_ABP/PPG_SPP_wrtDN | The ratio of the ABP/PPG cardiac cycle systolic phase peak amplitude with respect to diastolic phase peak to the ABP/PPG cardiac cycle systolic phase peak amplitude with respect to dicrotic notch |
| 124 | ABP/PPG_AM_RT_SPP_wrtDPE_ABP/PPG_SPP_wrtDN | The ratio of the ABP/PPG cardiac cycle systolic phase peak amplitude with respect to diastolic phase end point to the ABP/PPG cardiac cycle systolic phase peak amplitude with respect to dicrotic notch |
| 125 | ABP/PPG_AM_RT_SPP_wrtDPE_ABP/PPG_SPP_wrtDPP | The ratio of the ABP/PPG cardiac cycle systolic phase peak amplitude with respect to diastolic phase end point to the ABP/PPG cardiac cycle systolic phase peak amplitude with respect to diastolic phase peak |
| 126 | ABP/PPG_AM_RT_DN_wrtSPO_ABP/PPG_DN_wrtZero | The ratio of the ABP/PPG cardiac cycle dicrotic notch amplitude with respect to systolic phase onset to the ABP/PPG dicrotic notch amplitude with respect to zero |
| 127 | ABP/PPG_AM_RT_DN_wrtDPE_ABP/PPG_DN_wrtZero | The ratio of the ABP/PPG cardiac cycle dicrotic notch amplitude with respect to diastolic phase end point to the ABP/PPG dicrotic notch amplitude with respect to zero |
| 128 | ABP/PPG_AM_RT_DN_wrtDPE_ABP/PPG_DN_wrtSPO | The ratio of the ABP/PPG cardiac cycle dicrotic notch amplitude with respect to diastolic phase end point to the ABP/PPG dicrotic notch amplitude with respect to systolic phase onset |
| 129 | ABP/PPG_AM_RT_DPP_wrtSPO_ABP/PPG_DPP_wrtZero | The ratio of the ABP/PPG cardiac cycle diastolic phase peak amplitude with respect to systolic phase onset to the ABP/PPG diastolic phase peak with respect to zero |
| 130 | ABP/PPG_AM_RT_DPP_wrtDN_ABP/PPG_DPP_wrtZero | The ratio of the ABP/PPG cardiac cycle diastolic phase peak amplitude with respect to dicrotic notch to the ABP/PPG diastolic phase peak with respect to zero |
| 131 | ABP/PPG_AM_RT_DPP_wrtDPE_ABP/PPG_DPP_wrtZero | The ratio of the ABP/PPG cardiac cycle diastolic phase peak amplitude with respect to diastolic phase end point to the ABP/PPG diastolic phase peak with respect to zero |
| 132 | ABP/PPG_AM_RT_DPP_wrtDN_ABP/PPG_DPP_wrtSPO | The ratio of the ABP/PPG cardiac cycle diastolic phase peak amplitude with respect to dicrotic notch to the ABP/PPG diastolic phase peak with respect to systolic phase onset |
| 133 | ABP/PPG_AM_RT_DPP_wrtDPE_ABP/PPG_DPP_wrtSPO | The ratio of the ABP/PPG cardiac cycle diastolic phase peak amplitude with respect to diastolic phase end point to the ABP/PPG diastolic phase peak with respect to systolic phase onset |
| 134 | ABP/PPG_AM_RT_DPP_wrtDN_ABP/PPG_DPP_wrtDPE | The ratio of the ABP/PPG cardiac cycle diastolic phase peak amplitude with respect to dicrotic notch to the ABP/PPG diastolic phase peak with respect to diastolic phase end point |
| 135 | ABP/PPG_AM_RT_DPE_wrtSPO_ABP/PPG_DPE_wrtZero | The ratio of the ABP/PPG cardiac cycle diastolic phase end point amplitude with respect to systolic phase onset to the ABP/PPG diastolic phase end point with respect to zero |

3. **Normalized amplitude ratio features (136-240)**

| 136 | ABP/PPG_NAM_RT_SPP_wrtZero_ABP/PPG_SPO_wrtZero | The ratio of the normalized ABP/PPG cardiac cycle systolic phase peak amplitude with respect to zero to the normalized ABP/PPG cardiac cycle systolic phase onset amplitude with respect to zero |
| --- | --- | --- |
| 137 | ABP/PPG_NAM_RT_SPP_wrtSPO_ABP/PPG_SPO_wrtZero | The ratio of the normalized ABP/PPG cardiac cycle systolic phase peak amplitude with respect to systolic phase onset to the normalized ABP/PPG cardiac cycle systolic phase onset amplitude with respect to zero |
| 138 | ABP/PPG_NAM_RT_SPP_wrtDN_ABP/PPG_SPO_wrtZero | The ratio of the normalized ABP/PPG cardiac cycle systolic phase peak amplitude with respect to dicrotic notch to the normalized ABP/PPG cardiac cycle systolic phase onset amplitude with respect to zero |
| 139 | ABP/PPG_NAM_RT_SPP_wrtDPP_ABP/PPG_SPO_wrtZero | The ratio of the normalized ABP/PPG cardiac cycle systolic phase peak amplitude with respect to diastolic phase peak to the normalized ABP/PPG cardiac cycle systolic phase onset amplitude with respect to zero |
| 140 | ABP/PPG_NAM_RT_SPP_wrtDPE_ABP/PPG_SPO_wrtZero | The ratio of the normalized ABP/PPG cardiac cycle systolic phase peak amplitude with respect to diastolic phase end point to the normalized ABP/PPG cardiac cycle systolic phase onset amplitude with respect to zero |
| 141 | ABP/PPG_NAM_RT_DN_wrtZero_ABP/PPG_SPO_wrtZero | The ratio of the normalized ABP/PPG cardiac cycle dicrotic notch with respect to zero to the normalized ABP/PPG cardiac cycle systolic phase onset amplitude with respect to zero |
| 142 | ABP/PPG_NAM_RT_DN_wrtSPO_ABP/PPG_SPO_wrtZero | The ratio of the normalized ABP/PPG cardiac cycle dicrotic notch with respect to systolic phase onset to the normalized ABP/PPG cardiac cycle systolic phase onset amplitude with respect to zero |
| 143 | ABP/PPG_NAM_RT_DN_wrtDPE_ABP/PPG_SPO_wrtZero | The ratio of the normalized ABP/PPG cardiac cycle dicrotic notch with respect to diastolic phase end point to the normalized ABP/PPG cardiac cycle systolic phase onset amplitude with respect to zero |
| 144 | ABP/PPG_NAM_RT_DPP_wrtZero_ABP/PPG_SPO_wrtZero | The ratio of the normalized ABP/PPG cardiac cycle diastolic phase peak with respect to zero to the normalized ABP/PPG cardiac cycle systolic phase onset amplitude with respect to zero |
| 145 | ABP/PPG_NAM_RT_DPP_wrtSPO_ABP/PPG_SPOwrtZero | The ratio of the normalized ABP/PPG cardiac cycle diastolic phase peak amplitude with respect to systolic phase onset to the normalized ABP/PPG cardiac cycle systolic phase onset amplitude with respect to zero |
| 146 | ABP/PPG_NAM_RT_DPP_wrtDN_ABP/PPG_SPO_wrtZero | The ratio of the normalized ABP/PPG cardiac cycle diastolic phase peak amplitude with respect to dicrotic notch to the normalized ABP/PPG cardiac cycle systolic phase onset amplitude with respect to zero |
| 147 | ABP/PPG_NAM_RT_DPP_wrtDPE_ABP/PPG_SPOwrtZero | The ratio of the normalized ABP/PPG cardiac cycle diastolic phase peak with respect to diastolic phase end point to the normalized ABP/PPG cardiac cycle systolic phase onset amplitude with respect to zero |
| 148 | ABP/PPG_NAM_RT_DPE_wrtZero_ABP/PPG_SPO_wrtZero | The ratio of the normalized ABP/PPG cardiac cycle diastolic phase end point with respect to zero to the normalized ABP/PPG cardiac cycle systolic phase onset amplitude with respect to zero |
| 149 | ABP/PPG_NAM_RT_DPE_wrtSPO_ABP/PPG_SPO_wrtZero | The ratio of the normalized ABP/PPG cardiac cycle diastolic phase end point amplitude with respect to systolic phase onset to the normalized ABP/PPG cardiac cycle systolic phase onset amplitude with respect to zero |
| 150 | ABP/PPG_NAM_RT_DN_wrtZero_ABP/PPG_SPP_wrtZero | The ratio of the normalized ABP/PPG cardiac cycle dicrotic notch with respect to zero to the normalized ABP/PPG cardiac cycle systolic phase peak amplitude with respect to zero |
| 151 | ABP/PPG_NAM_RT_DN_wrtSPO_ABP/PPG_SPP_wrtZero | The ratio of the normalized ABP/PPG cardiac cycle dicrotic notch with respect to systolic phase onset to the normalized ABP/PPG cardiac cycle systolic phase peak amplitude with respect to zero |
| 152 | ABP/PPG_NAM_RT_DN_wrtDPE_ABP/PPG_SPP_wrtZero | The ratio of the normalized ABP/PPG cardiac cycle dicrotic notch with respect to diastolic phase end point to the normalized ABP/PPG cardiac cycle systolic phase peak amplitude with respect to zero |
| 153 | ABP/PPG_NAM_RT_DPP_wrtZero_ABP/PPG_SPP_wrtZero | The ratio of the normalized ABP/PPG cardiac cycle diastolic phase peak with respect to zero to the normalized ABP/PPG cardiac cycle systolic phase peak amplitude with respect to zero |
| 154 | ABP/PPG_NAM_RT_DPP_wrtSPO_ABP/PPG_SPP_wrtZero | The ratio of the normalized ABP/PPG cardiac cycle diastolic phase peak amplitude with respect to systolic phase onset to the normalized ABP/PPG cardiac cycle systolic phase peak amplitude with respect to zero |
| 155 | ABP/PPG_NAM_RT_DPP_wrtDN_ABP/PPG_SPP_wrtZero | The ratio of the normalized ABP/PPG cardiac cycle diastolic phase peak amplitude with respect to dicrotic notch to the normalized ABP/PPG cardiac cycle systolic phase peak amplitude with respect to zero |
| 156 | ABP/PPG_NAM_RT_DPP_wrtDPE_ABP/PPG_SPP_wrtZero | The ratio of the normalized ABP/PPG cardiac cycle diastolic phase peak with respect to diastolic phase end point to the normalized ABP/PPG cardiac cycle systolic phase peak amplitude with respect to zero |
| 157 | ABP/PPG_NAM_RT_DPE_wrtZero_ABP/PPG_SPP_wrtZero | The ratio of the normalized ABP/PPG cardiac cycle diastolic phase end point with respect to zero to the normalized ABP/PPG cardiac cycle systolic phase peak amplitude with respect to zero |
| 158 | ABP/PPG_NAM_RT_DPE_wrtSPO_ABP/PPG_SPP_wrtZero | The ratio of the normalized ABP/PPG cardiac cycle diastolic phase end point amplitude with respect to systolic phase onset to the normalized ABP/PPG cardiac cycle systolic phase peak amplitude with respect to zero |
| 159 | ABP/PPG_NAM_RT_DN_wrtZero_ABP/PPG_SPP_wrtSPO | The ratio of the normalized ABP/PPG cardiac cycle dicrotic notch with respect to zero to the normalized ABP/PPG cardiac cycle systolic phase peak amplitude with respect to systolic phase onset |
| 160 | ABP/PPG_NAM_RT_DN_wrtSPO_ABP/PPG_SPP_wrtSPO | The ratio of the normalized ABP/PPG cardiac cycle dicrotic notch with respect to systolic phase onset to the normalized ABP/PPG cardiac cycle systolic phase peak amplitude with respect to systolic phase onset |
| 161 | ABP/PPG_NAM_RT_DN_wrtDPE_ABP/PPG_SPP_wrtSPO | The ratio of the normalized ABP/PPG cardiac cycle dicrotic notch with respect to diastolic phase end point to the normalized ABP/PPG cardiac cycle systolic phase peak amplitude with respect to systolic phase onset |
| 162 | ABP/PPG_NAM_RT_DPP_wrtZero_ABP/PPG_SPP_wrtSPO | The ratio of the normalized ABP/PPG cardiac cycle diastolic phase peak with respect to zero to the normalized ABP/PPG cardiac cycle systolic phase peak amplitude with respect to systolic phase onset |
| 163 | ABP/PPG_NAM_RT_DPP_wrtSPO_ABP/PPG_SPP_wrtSPO | The ratio of the normalized ABP/PPG cardiac cycle diastolic phase peak amplitude with respect to systolic phase onset to the normalized ABP/PPG cardiac cycle systolic phase peak amplitude with respect to systolic phase onset |
| 164 | ABP/PPG_NAM_RT_DPP_wrtDN_ABP/PPG_SPP_wrtSPO | The ratio of the normalized ABP/PPG cardiac cycle diastolic phase peak amplitude with respect to dicrotic notch to the normalized ABP/PPG cardiac cycle systolic phase peak amplitude with respect to systolic phase onset |
| 165 | ABP/PPG_NAM_RT_DPP_wrtDPE_ABP/PPG_SPP_wrtSPO | The ratio of the normalized ABP/PPG cardiac cycle diastolic phase peak with respect to diastolic phase end point to the normalized ABP/PPG cardiac cycle systolic phase peak amplitude with respect to systolic phase onset |
| 166 | ABP/PPG_NAM_RT_DPE_wrtZero_ABP/PPG_SPP_wrtSPO | The ratio of the normalized ABP/PPG cardiac cycle diastolic phase end point with respect to zero to the normalized ABP/PPG cardiac cycle systolic phase peak amplitude with respect to systolic phase onset |
| 167 | ABP/PPG_NAM_RT_DPE_wrtSPO_ABP/PPG_SPP_wrtSPO | The ratio of the normalized ABP/PPG cardiac cycle diastolic phase end point amplitude with respect to systolic phase onset to the normalized ABP/PPG cardiac cycle systolic phase peak amplitude with respect to systolic phase onset |
| 168 | ABP/PPG_NAM_RT_DN_wrtZero_ABP/PPG_SPP_wrtDN | The ratio of the normalized ABP/PPG cardiac cycle dicrotic notch with respect to zero to the normalized ABP/PPG cardiac cycle systolic phase peak amplitude with respect to dicrotic notch |
| 169 | ABP/PPG_NAM_RT_DN_wrtSPO_ABP/PPG_SPP_wrtDN | The ratio of the normalized ABP/PPG cardiac cycle dicrotic notch with respect to systolic phase onset to the normalized ABP/PPG cardiac cycle systolic phase peak amplitude with respect to dicrotic notch |
| 170 | ABP/PPG_NAM_RT_DN_wrtDPE_ABP/PPG_SPP_wrtDN | The ratio of the normalized ABP/PPG cardiac cycle dicrotic notch with respect to diastolic phase end point to the normalized ABP/PPG cardiac cycle systolic phase peak amplitude with respect to dicrotic notch |
| 171 | ABP/PPG_NAM_RT_DPP_wrtZero_ABP/PPG_SPP_wrtDN | The ratio of the normalized ABP/PPG cardiac cycle diastolic phase peak with respect to zero to the normalized ABP/PPG cardiac cycle systolic phase peak amplitude with respect to dicrotic notch |
| 172 | ABP/PPG_NAM_RT_DPP_wrtSPO_ABP/PPG_SPP_wrtDN | The ratio of the normalized ABP/PPG cardiac cycle diastolic phase peak amplitude with respect to systolic phase onset to the normalized ABP/PPG cardiac cycle systolic phase peak amplitude with respect to dicrotic notch |
| 173 | ABP/PPG_NAM_RT_DPP_wrtDN_ABP/PPG_SPP_wrtDN | The ratio of the normalized ABP/PPG cardiac cycle diastolic phase peak amplitude with respect to dicrotic notch to the normalized ABP/PPG cardiac cycle systolic phase peak amplitude with respect to dicrotic notch |
| 174 | ABP/PPG_NAM_RT_DPP_wrtDPE_ABP/PPG_SPP_wrtDN | The ratio of the normalized ABP/PPG cardiac cycle diastolic phase peak with respect to diastolic phase end point to the normalized ABP/PPG cardiac cycle systolic phase peak amplitude with respect to dicrotic notch |
| 175 | ABP/PPG_NAM_RT_DPE_wrtZero_ABP/PPG_SPP_wrtDN | The ratio of the normalized ABP/PPG cardiac cycle diastolic phase end point with respect to zero to the normalized ABP/PPG cardiac cycle systolic phase peak amplitude with respect to dicrotic notch |
| 176 | ABP/PPG_NAM_RT_DPE_wrtSPO_ABP/PPG_SPP_wrtDN | The ratio of the normalized ABP/PPG cardiac cycle diastolic phase end point amplitude with respect to systolic phase onset to the normalized ABP/PPG cardiac cycle systolic phase peak amplitude with respect to dicrotic notch |
| 177 | ABP/PPG_NAM_RT_DN_wrtZero_ABP/PPG_SPP_wrtDPP | The ratio of the normalized ABP/PPG cardiac cycle dicrotic notch with respect to zero to the normalized ABP/PPG cardiac cycle systolic phase peak amplitude with respect to diastolic phase peak |
| 178 | ABP/PPG_NAM_RT_DN_wrtSPO_ABP/PPG_SPP_wrtDPP | The ratio of the normalized ABP/PPG cardiac cycle dicrotic notch with respect to systolic phase onset to the normalized ABP/PPG cardiac cycle systolic phase peak amplitude with respect to diastolic phase peak |
| 179 | ABP/PPG_NAM_RT_DN_wrtDPE_ABP/PPG_SPP_wrtDPP | The ratio of the normalized ABP/PPG cardiac cycle dicrotic notch with respect to diastolic phase end point to the normalized ABP/PPG cardiac cycle systolic phase peak amplitude with respect to diastolic phase peak |
| 180 | ABP/PPG_NAM_RT_DPP_wrtZero_ABP/PPG_SPP_wrtDPP | The ratio of the normalized ABP/PPG cardiac cycle diastolic phase peak with respect to zero to the normalized ABP/PPG cardiac cycle systolic phase peak amplitude with respect to diastolic phase peak |
| 181 | ABP/PPG_NAM_RT_DPP_wrtSPO_ABP/PPG_SPP_wrtDPP | The ratio of the normalized ABP/PPG cardiac cycle diastolic phase peak amplitude with respect to systolic phase onset to the normalized ABP/PPG cardiac cycle systolic phase peak amplitude with respect to diastolic phase peak |
| 182 | ABP/PPG_NAM_RT_DPP_wrtDN_ABP/PPG_SPP_wrtDPP | The ratio of the normalized ABP/PPG cardiac cycle diastolic phase peak amplitude with respect to dicrotic notch to the normalized ABP/PPG cardiac cycle systolic phase peak amplitude with respect to diastolic phase peak |
| 183 | ABP/PPG_NAM_RT_DPP_wrtDPE_ABP/PPG_SPP_wrtDPP | The ratio of the normalized ABP/PPG cardiac cycle diastolic phase peak with respect to diastolic phase end point to the normalized ABP/PPG cardiac cycle systolic phase peak amplitude with respect to diastolic phase peak |
| 184 | ABP/PPG_NAM_RT_DPE_wrtZero_ABP/PPG_SPP_wrtDPP | The ratio of the normalized ABP/PPG cardiac cycle diastolic phase end point with respect to zero to the normalized ABP/PPG cardiac cycle systolic phase peak amplitude with respect to diastolic phase peak |
| 185 | ABP/PPG_NAM_RT_DPE_wrtSPO_ABP/PPG_SPP_wrtDPP | The ratio of the normalized ABP/PPG cardiac cycle diastolic phase end point amplitude with respect to systolic phase onset to the normalized ABP/PPG cardiac cycle systolic phase peak amplitude with respect to diastolic phase peak |
| 186 | ABP/PPG_NAM_RT_DN_wrtZero_ABP/PPG_SPP_wrtDPE | The ratio of the normalized ABP/PPG cardiac cycle dicrotic notch with respect to zero to the normalized ABP/PPG cardiac cycle systolic phase peak amplitude with respect to diastolic phase end point |
| 187 | ABP/PPG_NAM_RT_DN_wrtSPO_ABP/PPG_SPP_wrtDPE | The ratio of the normalized ABP/PPG cardiac cycle dicrotic notch with respect to systolic phase onset to the normalized ABP/PPG cardiac cycle systolic phase peak amplitude with respect to diastolic phase end point |
| 188 | ABP/PPG_NAM_RT_DN_wrtDPE_ABP/PPG_SPP_wrtDPE | The ratio of the normalized ABP/PPG cardiac cycle dicrotic notch with respect to diastolic phase end point to the normalized ABP/PPG cardiac cycle systolic phase peak amplitude with respect to diastolic phase end point |
| 189 | ABP/PPG_NAM_RT_DPP_wrtZero_ABP/PPG_SPP_wrtDPE | The ratio of the normalized ABP/PPG cardiac cycle diastolic phase peak with respect to zero to the normalized ABP/PPG cardiac cycle systolic phase peak amplitude with respect to diastolic phase end point |
| 190 | ABP/PPG_NAM_RT_DPP_wrtSPO_ABP/PPG_SPP_wrtDPE | The ratio of the normalized ABP/PPG cardiac cycle diastolic phase peak amplitude with respect to systolic phase onset to the normalized ABP/PPG cardiac cycle systolic phase peak amplitude with respect to diastolic phase end point |
| 191 | ABP/PPG_NAM_RT_DPP_wrtDN_ABP/PPG_SPP_wrtDPE | The ratio of the normalized ABP/PPG cardiac cycle diastolic phase peak amplitude with respect to dicrotic notch to the normalized ABP/PPG cardiac cycle systolic phase peak amplitude with respect to diastolic phase end point |
| 192 | ABP/PPG_NAM_RT_DPP_wrtDPE_ABP/PPG_SPP_wrtDPE | The ratio of the normalized ABP/PPG cardiac cycle diastolic phase peak with respect to diastolic phase end point to the normalized ABP/PPG cardiac cycle systolic phase peak amplitude with respect to diastolic phase end point |
| 193 | ABP/PPG_NAM_RT_DPE_wrtZero_ABP/PPG_SPP_wrtDPE | The ratio of the normalized ABP/PPG cardiac cycle diastolic phase end point with respect to zero to the normalized ABP/PPG cardiac cycle systolic phase peak amplitude with respect to diastolic phase end point |
| 194 | ABP/PPG_NAM_RT_DPE_wrtSPO_ABP/PPG_SPP_wrtDPE | The ratio of the normalized ABP/PPG cardiac cycle diastolic phase end point amplitude with respect to systolic phase onset to the normalized ABP/PPG cardiac cycle systolic phase peak amplitude with respect to diastolic phase end point |
| 195 | ABP/PPG_NAM_RT_DPP_wrtZero_ABP/PPG_DN_wrtZero | The ratio of the normalized ABP/PPG cardiac cycle diastolic phase peak amplitude with respect to zero to the normalized ABP/PPG cardiac cycle dicrotic notch amplitude with respect to zero |
| 196 | ABP/PPG_NAM_RT_DPP_wrtSPO_ABP/PPG_DN_wrtZero | The ratio of the normalized ABP/PPG cardiac cycle diastolic phase peak amplitude with respect to systolic phase onset to the normalized ABP/PPG cardiac cycle dicrotic notch amplitude with respect to zero |
| 197 | ABP/PPG_NAM_RT_DPP_wrtDN_ABP/PPG_DN_wrtZero | The ratio of the normalized ABP/PPG cardiac cycle diastolic phase peak amplitude with respect to dicrotic notch to the normalized ABP/PPG cardiac cycle dicrotic notch amplitude with respect to zero |
| 198 | ABP/PPG_NAM_RT_DPP_wrtDPE_ABP/PPG_DN_wrtZero | The ratio of the normalized ABP/PPG cardiac cycle diastolic phase peak amplitude with respect to diastolic phase end point to the normalized ABP/PPG cardiac cycle dicrotic notch amplitude with respect to zero |
| 199 | ABP/PPG_NAM_RT_DPE_wrtZero_ABP/PPG_DN_wrtZero | The ratio of the normalized ABP/PPG cardiac cycle diastolic phase end point amplitude with respect to zero to the normalized ABP/PPG cardiac cycle dicrotic notch amplitude with respect to zero |
| 200 | ABP/PPG_NAM_RT_DPE_wrtSPO_ABP/PPG_DN_wrtZero | The ratio of the normalized ABP/PPG cardiac cycle diastolic phase end point amplitude with respect to systolic phase onset to the normalized ABP/PPG cardiac cycle dicrotic notch amplitude with respect to zero |
| 201 | ABP/PPG_NAM_RT_DPP_wrtZero_ABP/PPG_DN_wrtSPO | The ratio of the normalized ABP/PPG cardiac cycle diastolic phase peak amplitude with respect to zero to the normalized ABP/PPG cardiac cycle dicrotic notch amplitude with respect to systolic phase onset |
| 202 | ABP/PPG_NAM_RT_DPP_wrtSPO_ABP/PPG_DN_wrtSPO | The ratio of the normalized ABP/PPG cardiac cycle diastolic phase peak amplitude with respect to systolic phase onset to the normalized ABP/PPG cardiac cycle dicrotic notch amplitude with respect to systolic phase onset |
| 203 | ABP/PPG_NAM_RT_DPP_wrtDN_ABP/PPG_DN_wrtSPO | The ratio of the normalized ABP/PPG cardiac cycle diastolic phase peak amplitude with respect to dicrotic notch to the normalized ABP/PPG cardiac cycle dicrotic notch amplitude with respect to systolic phase onset |
| 204 | ABP/PPG_NAM_RT_DPP_wrtDPE_ABP/PPG_DN_wrtSPO | The ratio of the normalized ABP/PPG cardiac cycle diastolic phase peak amplitude with respect to diastolic phase end point to the normalized ABP/PPG cardiac cycle dicrotic notch amplitude with respect to systolic phase onset |
| 205 | ABP/PPG_NAM_RT_DPE_wrtZero_ABP/PPG_DN_wrtSPO | The ratio of the normalized ABP/PPG cardiac cycle diastolic phase end point amplitude with respect to zero to the normalized ABP/PPG cardiac cycle dicrotic notch amplitude with respect to systolic phase onset |
| 206 | ABP/PPG_NAM_RT_DPE_wrtSPO_ABP/PPG_DN_wrtSPO | The ratio of the normalized ABP/PPG cardiac cycle diastolic phase end point amplitude with respect to systolic phase onset to the normalized ABP/PPG cardiac cycle dicrotic notch amplitude with respect to systolic phase onset |
| 207 | ABP/PPG_NAM_RT_DPP_wrtZero_ABP/PPG_DN_wrtDPE | The ratio of the normalized ABP/PPG cardiac cycle diastolic phase peak amplitude with respect to zero to the normalized ABP/PPG cardiac cycle dicrotic notch amplitude with respect to diastolic phase end point |
| 208 | ABP/PPG_NAM_RT_DPP_wrtSPO_ABP/PPG_DN_wrtDPE | The ratio of the normalized ABP/PPG cardiac cycle diastolic phase peak amplitude with respect to systolic phase onset to the normalized ABP/PPG cardiac cycle dicrotic notch amplitude with respect to diastolic phase end point |
| 209 | ABP/PPG_NAM_RT_DPP_wrtDN_ABP/PPG_DN_wrtDPE | The ratio of the normalized ABP/PPG cardiac cycle diastolic phase peak amplitude with respect to dicrotic notch to the normalized ABP/PPG cardiac cycle dicrotic notch amplitude with respect to diastolic phase end point |
| 210 | ABP/PPG_NAM_RT_DPP_wrtDPE_ABP/PPG_DN_wrtDPE | The ratio of the normalized ABP/PPG cardiac cycle diastolic phase peak amplitude with respect to diastolic phase end point to the normalized ABP/PPG cardiac cycle dicrotic notch amplitude with respect to diastolic phase end point |
| 211 | ABP/PPG_NAM_RT_DPE_wrtZero_ABP/PPG_DN_wrtDPE | The ratio of the normalized ABP/PPG cardiac cycle diastolic phase end point amplitude with respect to zero to the normalized ABP/PPG cardiac cycle dicrotic notch amplitude with respect to diastolic phase end point |
| 212 | ABP/PPG_NAM_RT_DPE_wrtSPO_ABP/PPG_DN_wrtDPE | The ratio of the normalized ABP/PPG cardiac cycle diastolic phase end point amplitude with respect to systolic phase onset to the normalized ABP/PPG cardiac cycle dicrotic notch amplitude with respect to diastolic phase end point |
| 213 | ABP/PPG_NAM_RT_DPE_wrtZero_ABP/PPG_DPP_wrtZero | The ratio of the normalized ABP/PPG cardiac cycle diastolic phase end point amplitude with respect to zero to the normalized ABP/PPG cardiac cycle diastolic phase peak amplitude with respect to zero |
| 214 | ABP/PPG_NAM_RT_DPE_wrtSPO_ABP/PPG_DPP_wrtZero | The ratio of the normalized ABP/PPG cardiac cycle diastolic phase end point amplitude with respect to systolic phase onset to the normalized ABP/PPG cardiac cycle diastolic phase peak amplitude with respect to zero |
| 215 | ABP/PPG_NAM_RT_DPE_wrtZero_ABP/PPG_DPP_wrtSPO | The ratio of the normalized ABP/PPG cardiac cycle diastolic phase end point amplitude with respect to zero to the normalized ABP/PPG cardiac cycle diastolic phase peak amplitude with respect to systolic phase onset |
| 216 | ABP/PPG_NAM_RT_DPE_wrtSPO_ABP/PPG_DPP_wrtSPO | The ratio of the normalized ABP/PPG cardiac cycle diastolic phase end point amplitude with respect to systolic phase onset to the normalized ABP/PPG cardiac cycle diastolic phase peak amplitude with respect to systolic phase onset |
| 217 | ABP/PPG_NAM_RT_DPE_wrtZero_ABP/PPG_DPP_wrtDN | The ratio of the normalized ABP/PPG cardiac cycle diastolic phase end point amplitude with respect to zero to the normalized ABP/PPG cardiac cycle diastolic phase peak amplitude with respect to dicrotic notch |
| 218 | ABP/PPG_NAM_RT_DPE_wrtSPO_ABP/PPG_DPP_wrtDN | The ratio of the normalized ABP/PPG cardiac cycle diastolic phase end point amplitude with respect to systolic phase onset to the normalized ABP/PPG cardiac cycle diastolic phase peak amplitude with respect to dicrotic notch |
| 219 | ABP/PPG_NAM_RT_DPE_wrtZero_ABP/PPG_DPP_wrtDPE | The ratio of the normalized ABP/PPG cardiac cycle diastolic phase end point amplitude with respect to zero to the normalized ABP/PPG cardiac cycle diastolic phase peak amplitude with respect to diastolic phase end point |
| 220 | ABP/PPG_NAM_RT_DPE_wrtSPO_ABP/PPG_DPP_wrtDPE | The ratio of the normalized ABP/PPG cardiac cycle diastolic phase end point amplitude with respect to systolic phase onset to the normalized ABP/PPG cardiac cycle diastolic phase peak amplitude with respect to diastolic phase end point |
| 221 | ABP/PPG_NAM_RT_SPP_wrtSPO_ABP/PPG_SPP_wrtZero | The ratio of the normalized ABP/PPG cardiac cycle systolic phase peak amplitude with respect to systolic phase onset to the normalized ABP/PPG cardiac cycle systolic phase peak amplitude with respect to zero |
| 222 | ABP/PPG_NAM_RT_SPP_wrtDN_ABP/PPG_SPP_wrtZero | The ratio of the normalized ABP/PPG cardiac cycle systolic phase peak amplitude with respect to dicrotic notch to the normalized ABP/PPG cardiac cycle systolic phase peak amplitude with respect to zero |
| 223 | ABP/PPG_NAM_RT_SPP_wrtDPP_ABP/PPG_SPP_wrtZero | The ratio of the normalized ABP/PPG cardiac cycle systolic phase peak amplitude with respect to diastolic phase peak to the normalized ABP/PPG cardiac cycle systolic phase peak amplitude with respect to zero |
| 224 | ABP/PPG_NAM_RT_SPP_wrtDPE_ABP/PPG_SPP_wrtZero | The ratio of the normalized ABP/PPG cardiac cycle systolic phase peak amplitude with respect to diastolic phase end point to the normalized ABP/PPG cardiac cycle systolic phase peak amplitude with respect to zero |
| 225 | ABP/PPG_NAM_RT_SPP_wrtDN_ABP/PPG_SPP_wrtSPO | The ratio of the normalized ABP/PPG cardiac cycle systolic phase peak amplitude with respect to dicrotic notch to the normalized ABP/PPG cardiac cycle systolic phase peak amplitude with respect to systolic phase onset |
| 226 | ABP/PPG_NAM_RT_SPP_wrtDPP_ABP/PPG_SPP_wrtSPO | The ratio of the normalized ABP/PPG cardiac cycle systolic phase peak amplitude with respect to diastolic phase peak to the normalized ABP/PPG cardiac cycle systolic phase peak amplitude with respect to systolic phase onset |
| 227 | ABP/PPG_NAM_RT_SPP_wrtDPE_ABP/PPG_SPP_wrtSPO | The ratio of the normalized ABP/PPG cardiac cycle systolic phase peak amplitude with respect to diastolic phase end point to the normalized ABP/PPG cardiac cycle systolic phase peak amplitude with respect to systolic phase onset |
| 228 | ABP/PPG_NAM_RT_SPP_wrtDPP_ABP/PPG_SPP_wrtDN | The ratio of the normalized ABP/PPG cardiac cycle systolic phase peak amplitude with respect to diastolic phase peak to the normalized ABP/PPG cardiac cycle systolic phase peak amplitude with respect to systolic dicrotic notch |
| 229 | ABP/PPG_NAM_RT_SPP_wrtDPE_ABP/PPG_SPP_wrtDN | The ratio of the normalized ABP/PPG cardiac cycle systolic phase peak amplitude with respect to diastolic phase end point to the normalized ABP/PPG cardiac cycle systolic phase peak amplitude with respect to dicrotic notch |
| 230 | ABP/PPG_NAM_RT_SPP_wrtDPE_ABP/PPG_SPP_wrtDPP | The ratio of the normalized ABP/PPG cardiac cycle systolic phase peak amplitude with respect to diastolic phase end point to the normalized ABP/PPG cardiac cycle systolic phase peak amplitude with respect to diastolic phase peak |
| 231 | ABP/PPG_NAM_RT_DN_wrtSPO_ABP/PPG_DN_wrtZero | The ratio of the normalized ABP/PPG cardiac cycle dicrotic notch amplitude with respect to systolic phase onset to the normalized ABP/PPG cardiac cycle dicrotic notch amplitude with respect to zero |
| 232 | ABP/PPG_NAM_RT_DN_wrtDPE_ABP/PPG_DN_wrtZero | The ratio of the normalized ABP/PPG cardiac cycle dicrotic notch amplitude with respect to diastolic phase end point to the normalized ABP/PPG cardiac cycle dicrotic notch amplitude with respect to zero |
| 233 | ABP/PPG_NAM_RT_DN_wrtDPE_ABP/PPG_DN_wrtSPO | The ratio of the normalized ABP/PPG cardiac cycle dicrotic notch amplitude with respect to diastolic phase end point to the normalized ABP/PPG cardiac cycle dicrotic notch amplitude with respect to systolic phase onset |
| 234 | ABP/PPG_NAM_RT_DPP_wrtSPO_ABP/PPG_DPP_wrtZero | The ratio of the normalized ABP/PPG cardiac cycle diastolic phase peak amplitude with respect to systolic phase onset to the normalized ABP/PPG cardiac cycle diastolic phase peak amplitude with respect to zero |
| 235 | ABP/PPG_NAM_RT_DPP_wrtDN_ABP/PPG_DPP_wrtZero | The ratio of the normalized ABP/PPG cardiac cycle diastolic phase peak amplitude with respect to dicrotic notch to the normalized ABP/PPG cardiac cycle diastolic phase peak amplitude with respect to zero |
| 236 | ABP/PPG_NAM_RT_DPP_wrtDPE_ABP/PPG_DPP_wrtZero | The ratio of the normalized ABP/PPG cardiac cycle diastolic phase peak amplitude with respect to diastolic phase end point to the normalized ABP/PPG cardiac cycle diastolic phase peak amplitude with respect to zero |
| 237 | ABP/PPG_NAM_RT_DPP_wrtDN_ABP/PPG_DPP_wrtSPO | The ratio of the normalized ABP/PPG cardiac cycle diastolic phase peak amplitude with respect to dicrotic notch to the normalized ABP/PPG cardiac cycle diastolic phase peak amplitude with respect to systolic phase onset |
| 238 | ABP/PPG_NAM_RT_DPP_wrtDPE_ABP/PPG_DPP_wrtSPO | The ratio of the normalized ABP/PPG cardiac cycle diastolic phase peak amplitude with respect to diastolic phase end point to the normalized ABP/PPG cardiac cycle diastolic phase peak amplitude with respect to systolic phase onset |
| 239 | ABP/PPG_NAM_RT_DPP_wrtDN_ABP/PPG_DPP_wrtDPE | The ratio of the normalized ABP/PPG cardiac cycle diastolic phase peak amplitude with respect to dicrotic notch to the normalized ABP/PPG cardiac cycle diastolic phase peak amplitude with respect to diastolic phase end point |
| 240 | ABP/PPG_NAM_RT_DPE_wrtSPO_ABP/PPG_DPE_wrtZero | The ratio of the normalized ABP/PPG cardiac cycle diastolic phase end point amplitude with respect to systolic phase onset to the normalized ABP/PPG cardiac cycle diastolic phase end point amplitude with respect to zero |

4. **Duration features (241-250)**

| 241 | ABP/PPG_D_SPO_wrtSPP | Duration of the ABP/PPG cardiac cycle systolic phase onset with respect to systolic phase peak |
| --- | --- | --- |
| 242 | ABP/PPG_D_SPO_wrtDN | Duration of the ABP/PPG cardiac cycle systolic phase onset with respect to dicrotic notch |
| 243 | ABP/PPG_D_SPO_wrtDPP | Duration of the ABP/PPG cardiac cycle systolic phase onset with respect to diastolic phase peak |
| 244 | ABP/PPG_D_SPO_wrtDPE | Duration of the ABP/PPG cardiac cycle systolic phase onset with respect to diastolic phase end point |
| 245 | ABP/PPG_D_SPP_wrtDN | Duration of the ABP/PPG cardiac cycle systolic phase peak with respect to dicrotic notch |
| 246 | ABP/PPG_D_SPP_wrtDPP | Duration of the ABP/PPG cardiac cycle systolic phase peak with respect to diastolic phase peak |
| 247 | ABP/PPG_D_SPP_wrtDPE | Duration of the ABP/PPG cardiac cycle systolic phase peak with respect to diastolic phase end point |
| 248 | ABP/PPG_D_DN_wrtDPP | Duration of the ABP/PPG cardiac cycle dicrotic notch with respect to diastolic phase peak |
| 249 | ABP/PPG_D_DN_wrtDPE | Duration of the ABP/PPG cardiac cycle dicrotic notch with respect to diastolic phase end point |
| 250 | ABP/PPG_D_DPP_wrtDPE | Duration of the ABP/PPG cardiac cycle diastolic phase peak with respect to diastolic phase end point |

5. **Duration ratio features (251-296)**

| 251 | ABP/PPG_D_RT_SPP_wrtDN_ABP/PPG_SPO_wrtSPP | The ratio of the ABP/PPG cardiac cycle systolic phase peak duration with respect to dicrotic notch to the ABP/PPG systolic phase onset duration with respect to systolic phase peak |
| --- | --- | --- |
| 252 | ABP/PPG_D_RT_SPP_wrtDPP_ABP/PPG_SPO_wrtSPP | The ratio of the ABP/PPG cardiac cycle systolic phase peak duration with respect to diastolic phase peak to the ABP/PPG cardiac cycle systolic phase onset duration with respect to systolic phase peak |
| 253 | ABP/PPG_D_RT_SPP_wrtDPE_ABP/PPG_SPO_wrtSPP | The ratio of the ABP/PPG cardiac cycle systolic phase peak duration with respect to diastolic phase end point to the ABP/PPG cardiac cycle systolic phase onset duration with respect to systolic phase peak |
| 254 | ABP/PPG_D_RT_DN_wrtDPP_ABP/PPG_SPO_wrtSPP | The ratio of the ABP/PPG cardiac cycle dicrotic notch duration with respect to diastolic phase peak to the ABP/PPG cardiac cycle systolic phase onset duration with respect to systolic phase peak |
| 255 | ABP/PPG_D_RT_DN_wrtDPE_ABP/PPG_SPO_wrtSPP | The ratio of the ABP/PPG cardiac cycle dicrotic notch duration with respect to diastolic phase end point to the ABP/PPG cardiac cycle systolic phase onset duration with respect to systolic phase peak |
| 256 | ABP/PPG_D_RT_DPP_wrtDPE_ABP/PPG_SPO_wrtSPP | The ratio of the ABP/PPG cardiac cycle diastolic phase peak duration with respect to diastolic phase end point to the ABP/PPG cardiac cycle systolic phase onset duration with respect to systolic phase peak |
| 257 | ABP/PPG_D_RT_SPP_wrtDN_ABP/PPG_SPO_wrtDN | The ratio of the ABP/PPG cardiac cycle systolic phase peak duration with respect to dicrotic notch to the ABP/PPG cardiac cycle systolic phase onset duration with respect to dicrotic notch |
| 258 | ABP/PPG_D_RT_SPP_wrtDPP_ABP/PPG_SPO_wrtDN | The ratio of the ABP/PPG cardiac cycle systolic phase peak duration with respect to diastolic phase peak to the ABP/PPG cardiac cycle systolic phase onset duration with respect to dicrotic notch |
| 259 | ABP/PPG_D_RT_SPP_wrtDPE_ABP/PPG_SPO_wrtDN | The ratio of the ABP/PPG cardiac cycle systolic phase peak duration with respect to diastolic phase end point to the ABP/PPG cardiac cycle systolic phase onset duration with respect to dicrotic notch |
| 260 | ABP/PPG_D_RT_DN_wrtDPP_ABP/PPG_SPO_wrtDN | The ratio of the ABP/PPG cardiac cycle dicrotic notch duration with respect to diastolic phase peak to the ABP/PPG cardiac cycle systolic phase onset duration with respect to dicrotic notch |
| 261 | ABP/PPG_D_RT_DN_wrtDPE_ABP/PPG_SPO_wrtDN | The ratio of the ABP/PPG cardiac cycle dicrotic notch duration with respect to diastolic phase end point to the ABP/PPG cardiac cycle systolic phase onset duration with respect to dicrotic notch |
| 262 | ABP/PPG_D_RT_DPP_wrtDPE_ABP/PPG_SPO_wrtDN | The ratio of the ABP/PPG cardiac cycle diastolic phase peak duration with respect to diastolic phase end point to the ABP/PPG cardiac cycle systolic phase onset duration with respect to dicrotic notch |
| 263 | ABP/PPG_D_RT_SPP_wrtDN_ABP/PPG_SPO_wrtDN | The ratio of the ABP/PPG cardiac cycle systolic phase peak duration with respect to dicrotic notch to the ABP/PPG cardiac cycle systolic phase onset duration with respect to dicrotic notch |
| 264 | ABP/PPG_D_RT_SPP_wrtDPP_ABP/PPG_SPO_wrtDN | The ratio of the ABP/PPG cardiac cycle systolic phase peak duration with respect to diastolic phase peak to the ABP/PPG cardiac cycle systolic phase onset duration with respect to dicrotic notch |
| 265 | ABP/PPG_D_RT_SPP_wrtDPE_ABP/PPG_SPO_wrtDN | The ratio of the ABP/PPG cardiac cycle systolic phase peak duration with respect to diastolic phase end point to the ABP/PPG cardiac cycle systolic phase onset duration with respect to dicrotic notch |
| 266 | ABP/PPG_D_RT_DN_wrtDPP_ABP/PPG_SPO_wrtDN | The ratio of the ABP/PPG cardiac cycle dicrotic notch duration with respect to diastolic phase peak to the ABP/PPG cardiac cycle systolic phase onset duration with respect to dicrotic notch |
| 267 | ABP/PPG_D_RT_DN_wrtDPE_ABP/PPG_SPO_wrtDN | The ratio of the ABP/PPG cardiac cycle dicrotic notch duration with respect to diastolic phase end point to the ABP/PPG cardiac cycle systolic phase onset duration with respect to dicrotic notch |
| 268 | ABP/PPG_D_RT_DPP_wrtDPE_ABP/PPG_SPO_wrtDPP | The ratio of the ABP/PPG cardiac cycle diastolic phase peak duration with respect to diastolic phase end point to the ABP/PPG cardiac cycle systolic phase onset duration with respect to diastolic phase peak |
| 269 | ABP/PPG_D_RT_SPP_wrtDN_ABP/PPG_SPO_wrtDPE | The ratio of the ABP/PPG cardiac cycle systolic phase peak duration with respect to dicrotic notch to the ABP/PPG cardiac cycle systolic phase onset duration with respect to diastolic phase end point |
| 270 | ABP/PPG_D_RT_SPP_wrtDPP_ABP/PPG_SPO_wrtDPE | The ratio of the ABP/PPG cardiac cycle systolic phase peak duration with respect to diastolic phase peak to the ABP/PPG cardiac cycle systolic phase onset duration with respect to diastolic phase end point |
| 271 | ABP/PPG_D_RT_SPP_wrtDPE_ABP/PPG_SPO_wrtDPE | The ratio of the ABP/PPG cardiac cycle systolic phase peak duration with respect to diastolic phase end point to the ABP/PPG cardiac cycle systolic phase onset duration with respect to diastolic phase end point |
| 272 | ABP/PPG_D_RT_DN_wrtDPP_ABP/PPG_SPO_wrtDPE | The ratio of the ABP/PPG cardiac cycle dicrotic notch duration with respect to diastolic phase peak to the ABP/PPG cardiac cycle systolic phase onset duration with respect to diastolic phase end point |
| 273 | ABP/PPG_D_RT_DN_wrtDPE_ABP/PPG_SPO_wrtDPE | The ratio of the ABP/PPG cardiac cycle dicrotic notch duration with respect to diastolic phase end point to the ABP/PPG cardiac cycle systolic phase onset duration with respect to diastolic phase end point |
| 274 | ABP/PPG_D_RT_DPP_wrtDPE_ABP/PPG_SPO_wrtDPE | The ratio of the ABP/PPG cardiac cycle diastolic phase peak duration with respect to diastolic phase end point to the ABP/PPG cardiac cycle systolic phase onset duration with respect to diastolic phase end point |
| 275 | ABP/PPG_D_RT_DN_wrtDPP_ABP/PPG_SPP_wrtDN | The ratio of the ABP/PPG cardiac cycle dicrotic notch duration with respect to diastolic phase peak to the ABP/PPG cardiac cycle systolic phase peak duration with respect to dicrotic notch |
| 276 | ABP/PPG_D_RT_DN_wrtDPE_ABP/PPG_SPP_wrtDN | The ratio of the ABP/PPG cardiac cycle dicrotic notch duration with respect to diastolic phase end point to the ABP/PPG cardiac cycle systolic phase peak duration with respect to dicrotic notch |
| 277 | ABP/PPG_D_RT_DPP_wrtDPE_ABP/PPG_SPP_wrtDN | The ratio of the ABP/PPG cardiac cycle diastolic phase peak duration with respect to diastolic phase end point to the ABP/PPG cardiac cycle systolic phase peak duration with respect to dicrotic notch |
| 278 | ABP/PPG_D_RT_DN_wrtDPP_ABP/PPG_SPP_wrtDPP | The ratio of the ABP/PPG cardiac cycle dicrotic notch duration with respect to diastolic phase peak to the ABP/PPG cardiac cycle systolic phase peak duration with respect to diastolic phase peak |
| 279 | ABP/PPG_D_RT_DN_wrtDPE_ABP/PPG_SPP_wrtDPP | The ratio of the ABP/PPG cardiac cycle dicrotic notch duration with respect to diastolic phase end point to the ABP/PPG cardiac cycle systolic phase peak duration with respect to diastolic phase peak |
| 280 | ABP/PPG_D_RT_DPP_wrtDPE_ABP/PPG_SPP_wrtDPP | The ratio of the ABP/PPG cardiac cycle diastolic phase peak duration with respect to diastolic phase end point to the ABP/PPG cardiac cycle systolic phase peak duration with respect to diastolic phase peak |
| 281 | ABP/PPG_D_RT_DN_wrtDPP_ABP/PPG_SPP_wrtDPE | The ratio of the ABP/PPG cardiac cycle dicrotic notch duration with respect to diastolic phase peak to the ABP/PPG cardiac cycle systolic phase peak duration with respect to diastolic phase end point |
| 282 | ABP/PPG_D_RT_DN_wrtDPE_ABP/PPG_SPP_wrtDPE | The ratio of the ABP/PPG cardiac cycle dicrotic notch duration with respect to diastolic phase end point to the ABP/PPG cardiac cycle systolic phase peak duration with respect to diastolic phase end point |
| 283 | ABP/PPG_D_RT_DPP_wrtDPE_ABP/PPG_SPP_wrtDPE | The ratio of the ABP/PPG cardiac cycle diastolic phase peak duration with respect to diastolic phase end point to the ABP/PPG cardiac cycle systolic phase peak duration with respect to diastolic phase end point |
| 284 | ABP/PPG_D_RT_DPP_wrtDPE_ABP/PPG_DN_wrtDPP | The ratio of the ABP/PPG cardiac cycle diastolic phase peak duration with respect to diastolic phase end point to the ABP/PPG cardiac cycle dicrotic notch duration with respect to diastolic phase peak |
| 285 | ABP/PPG_D_RT_DPP_wrtDPE_ABP/PPG_DN_wrtDPE | The ratio of the ABP/PPG cardiac cycle diastolic phase peak duration with respect to diastolic phase end point to the ABP/PPG cardiac cycle dicrotic notch duration with respect to diastolic phase end point |
| 286 | ABP/PPG_D_RT_SPO_wrtDN_ABP/PPG_SPO_wrtSPP | The ratio of the ABP/PPG cardiac cycle systolic phase onset duration with respect to dicrotic notch to the ABP/PPG cardiac cycle systolic phase onset duration with respect to systolic phase peak |
| 287 | ABP/PPG_D_RT_SPO_wrtDPP_ABP/PPG_SPO_wrtSPP | The ratio of the ABP/PPG cardiac cycle systolic phase onset duration with respect to diastolic phase peak to the ABP/PPG cardiac cycle systolic phase onset duration with respect to systolic phase peak |
| 288 | ABP/PPG_D_RT_SPO_wrtDPE_ABP/PPG_SPO_wrtSPP | The ratio of the ABP/PPG cardiac cycle systolic phase onset duration with respect to diastolic phase end point to the ABP/PPG cardiac cycle systolic phase onset duration with respect to systolic phase peak |
| 289 | ABP/PPG_D_RT_SPO_wrtDPP_ABP/PPG_SPO_wrtDN | The ratio of the ABP/PPG cardiac cycle systolic phase onset duration with respect to diastolic phase peak to the ABP/PPG cardiac cycle systolic phase onset duration with respect to dicrotic notch |
| 290 | ABP/PPG_D_RT_SPO_wrtDPE_ABP/PPG_SPO_wrtDN | The ratio of the ABP/PPG cardiac cycle systolic phase onset duration with respect to diastolic phase end point to the ABP/PPG cardiac cycle systolic phase onset duration with respect to dicrotic notch |
| 291 | ABP/PPG_D_RT_SPO_wrtDPE_ABP/PPG_SPO_wrtDPP | The ratio of the ABP/PPG cardiac cycle systolic phase onset duration with respect to diastolic phase end point to the ABP/PPG cardiac cycle systolic phase onset duration with respect to diastolic phase peak |
| 292 | ABP/PPG_D_RT_SPP_wrtDPP_ABP/PPG_SPP_wrtDN | The ratio of the ABP/PPG cardiac cycle systolic phase peak duration with respect to diastolic phase peak to the ABP/PPG cardiac cycle systolic phase peak duration with respect to dicrotic notch |
| 293 | ABP/PPG_D_RT_SPP_wrtDPE_ABP/PPG_SPP_wrtDN | The ratio of the ABP/PPG cardiac cycle systolic phase peak duration with respect to diastolic phase end point to the ABP/PPG cardiac cycle systolic phase peak duration with respect to dicrotic notch |
| 294 | ABP/PPG_D_RT_SPP_wrtDPE_ABP/PPG_SPP_wrtDPP | The ratio of the ABP/PPG cardiac cycle systolic phase peak duration with respect to diastolic phase end point to the ABP/PPG cardiac cycle systolic phase peak duration with respect to diastolic phase peak |
| 295 | ABP/PPG_D_RT_DN_wrtDPE_ABP/PPG_DN_wrtDPP | The ratio of the ABP/PPG cardiac cycle dicrotic notch duration with respect to diastolic phase end point to the ABP/PPG cardiac cycle dicrotic notch duration with respect to diastolic phase peak |
| 296 | ABP/PPG_HR | The ratio of the ABP/PPG cardiac cycle |

6. **Average features (297-306)**

| 297 | ABP/PPG_D_Avg_SPO_wrtSPP | Average duration of the ABP/PPG cardiac cycle systolic phase onset with respect to systolic phase peak |
| --- | --- | --- |
| 298 | ABP/PPG_D_Avg_SPO_wrtDN | Average duration of the ABP/PPG cardiac cycle systolic phase onset with respect to dicrotic notch |
| 299 | ABP/PPG_D_Avg_SPO_wrtDPP | Average duration of the ABP/PPG cardiac cycle systolic phase onset with respect to diastolic phase peak |
| 300 | ABP/PPG_D_Avg_SPO_wrtDPE | Average duration of the ABP/PPG cardiac cycle systolic phase onset with respect to diastolic phase end point |
| 301 | ABP/PPG_D_Avg_SPP_wrtDN | Average duration of the ABP/PPG cardiac cycle systolic phase peak with respect to dicrotic notch |
| 302 | ABP/PPG_D_Avg_SPP_wrtDPP | Average duration of the ABP/PPG cardiac cycle systolic phase peak with respect to diastolic phase peak |
| 303 | ABP/PPG_D_Avg_SPP_wrtDPE | Average duration of the ABP/PPG cardiac cycle systolic phase peak with respect to diastolic phase end point |
| 304 | ABP/PPG_D_Avg_DN_wrtDPP | Average duration of the ABP/PPG cardiac cycle dicrotic notch with respect to diastolic phase peak |
| 305 | ABP/PPG_D_Avg_DN_wrtDPE | Average duration of the ABP/PPG cardiac cycle dicrotic notch with respect to diastolic phase end point |
| 306 | ABP/PPG_D_Avg_DPP_wrtDPE | Average duration of the ABP/PPG cardiac cycle diastolic phase peak with respect to diastolic phase end point |

7. **Average features with subtracted diastolic end point (307-316)**

| 307 | ABP/PPG_D_Avg_wsDPE_SPO_wrtSPP | Average duration of the ABP/PPG cardiac cycle systolic phase onset with respect to systolic phase peak, with subtracted diastolic phase end point |
| --- | --- | --- |
| 308 | ABP/PPG_D_Avg_wsDPE_SPO_wrtDN | Average duration of the ABP/PPG cardiac cycle systolic phase onset with respect to dicrotic notch, with subtracted diastolic phase end point |
| 309 | ABP/PPG_D_Avg_wsDPE_SPO_wrtDPP | Average duration of the ABP/PPG cardiac cycle systolic phase onset with respect to diastolic phase peak, with subtracted diastolic phase end point |
| 310 | ABP/PPG_D_Avg_wsDPE_SPO_wrtDPE | Average duration of the ABP/PPG cardiac cycle systolic phase onset with respect to diastolic phase end point, with subtracted diastolic phase end point |
| 311 | ABP/PPG_D_Avg_wsDPE_SPP_wrtDN | Average duration of the ABP/PPG cardiac cycle systolic phase peak with respect to dicrotic notch, with subtracted diastolic phase end point |
| 312 | ABP/PPG_D_Avg_wsDPE_SPP_wrtDPP | Average duration of the ABP/PPG cardiac cycle systolic phase peak with respect to diastolic phase peak, with subtracted diastolic phase end point |
| 313 | ABP/PPG_D_Avg_wsDPE_SPP_wrtDPE | Average duration of the ABP/PPG cardiac cycle systolic phase peak with respect to diastolic phase end point, with subtracted diastolic phase end point |
| 314 | ABP/PPG_D_Avg_wsDPE_DN_wrtDPP | Average duration of the ABP/PPG cardiac cycle dicrotic notch with respect to diastolic phase peak, with subtracted diastolic phase end point |
| 315 | ABP/PPG_D_Avg_wsDPE_DN_wrtDPE | Average duration of the ABP/PPG cardiac cycle dicrotic notch with respect to diastolic phase end point, with subtracted diastolic phase end point |
| 316 | ABP/PPG_D_Avg_wsDPE_DPP_wrtDPE | Average duration of the ABP/PPG cardiac cycle diastolic phase peak with respect to diastolic phase end point, with subtracted diastolic phase end point |

8. **Normalized average features (317-326)**

| 317 | ABP/PPG_D_NAvg_SPO_wrtSPP | Average duration of the normalized ABP/PPG cardiac cycle systolic phase onset with respect to systolic phase peak |
| --- | --- | --- |
| 318 | ABP/PPG_D_NAvg_SPO_wrtDN | Average duration of the normalized ABP/PPG cardiac cycle systolic phase onset with respect to dicrotic notch |
| 319 | ABP/PPG_D_NAvg_SPO_wrtDPP | Average duration of the normalized ABP/PPG cardiac cycle systolic phase onset with respect to diastolic phase peak |
| 320 | ABP/PPG_D_NAvg_SPO_wrtDPE | Average duration of the normalized ABP/PPG cardiac cycle systolic phase onset with respect to diastolic phase end point |
| 321 | ABP/PPG_D_NAvg_SPP_wrtDN | Average duration of the normalized ABP/PPG cardiac cycle systolic phase peak with respect to dicrotic notch |
| 322 | ABP/PPG_D_NAvg_SPP_wrtDPP | Average duration of the normalized ABP/PPG cardiac cycle systolic phase peak with respect to diastolic phase peak |
| 323 | ABP/PPG_D_NAvg_SPP_wrtDPE | Average duration of the normalized ABP/PPG cardiac cycle systolic phase peak with respect to diastolic phase end point |
| 324 | ABP/PPG_D_NAvg_DN_wrtDPP | Average duration of the normalized ABP/PPG cardiac cycle dicrotic notch with respect to diastolic phase peak |
| 325 | ABP/PPG_D_NAvg_DN_wrtDPE | Average duration of the normalized ABP/PPG cardiac cycle dicrotic notch with respect to diastolic phase end point |
| 326 | ABP/PPG_D_NAvg_DPP_wrtDPE | Average duration of the normalized ABP/PPG cardiac cycle diastolic phase peak with respect to diastolic phase end point |

9. **Normalized average features with subtracted diastolic end point (327-336)**

| 327 | ABP/PPG_D_NAvg_wsDPE_SPO_wrtSPP | Average duration of the normalized ABP/PPG cardiac cycle systolic phase onset with respect to systolic phase peak, with subtracted diastolic phase end point |
| --- | --- | --- |
| 328 | ABP/PPG_D_NAvg_wsDPE_SPO_wrtDN | Average duration of the normalized ABP/PPG cardiac cycle systolic phase onset with respect to dicrotic notch, with subtracted diastolic phase end point |
| 329 | ABP/PPG_D_NAvg_wsDPE_SPO_wrtDPP | Average duration of the normalized ABP/PPG cardiac cycle systolic phase onset with respect to diastolic phase peak, with subtracted diastolic phase end point |
| 330 | ABP/PPG_D_NAvg_wsDPE_SPO_wrtDPE | Average duration of the normalized ABP/PPG cardiac cycle systolic phase onset with respect to diastolic phase end point, with subtracted diastolic phase end point |
| 331 | ABP/PPG_D_NAvg_wsDPE_SPP_wrtDN | Average duration of the normalized ABP/PPG cardiac cycle systolic phase peak with respect to dicrotic notch, with subtracted diastolic phase end point |
| 332 | ABP/PPG_D_NAvg_wsDPE_SPP_wrtDPP | Average duration of the normalized ABP/PPG cardiac cycle systolic phase peak with respect to diastolic phase peak, with subtracted diastolic phase end point |
| 333 | ABP/PPG_D_NAvg_wsDPE_SPP_wrtDPE | Average duration of the normalized ABP/PPG cardiac cycle systolic phase peak with respect to diastolic phase end point, with subtracted diastolic phase end point |
| 334 | ABP/PPG_D_NAvg_wsDPE_DN_wrtDPP | Average duration of the normalized ABP/PPG cardiac cycle dicrotic notch with respect to diastolic phase peak, with subtracted diastolic phase end point |
| 335 | ABP/PPG_D_NAvg_wsDPE_DN_wrtDPE | Average duration of the normalized ABP/PPG cardiac cycle dicrotic notch with respect to diastolic phase end point, with subtracted diastolic phase end point |
| 336 | ABP/PPG_D_NAvg_wsDPE_DPP_wrtDPE | Average duration of the normalized ABP/PPG cardiac cycle diastolic phase peak with respect to diastolic phase end point, with subtracted diastolic phase end point |

10. **Median features (337-356)**

| 337 | ABP/PPG_D_Med_SPO_wrtSPP | Median duration of the ABP/PPG cardiac cycle systolic phase onset with respect to systolic phase peak |
| --- | --- | --- |
| 338 | ABP/PPG_D_Med_SPO_wrtDN | Median duration of the ABP/PPG cardiac cycle systolic phase onset with respect to dicrotic notch |
| 339 | ABP/PPG_D_Med_SPO_wrtDPP | Median duration of the ABP/PPG cardiac cycle systolic phase onset with respect to diastolic phase peak |
| 340 | ABP/PPG_D_Med_SPO_wrtDPE | Median duration of the ABP/PPG cardiac cycle systolic phase onset with respect to diastolic phase end point |
| 341 | ABP/PPG_D_Med_SPP_wrtDN | Median duration of the ABP/PPG cardiac cycle systolic phase peak with respect to dicrotic notch |
| 342 | ABP/PPG_D_Med_SPP_wrtDPP | Median duration of the ABP/PPG cardiac cycle systolic phase peak with respect to diastolic phase peak |
| 343 | ABP/PPG_D_Med_SPP_wrtDPE | Median duration of the ABP/PPG cardiac cycle systolic phase peak with respect to diastolic phase end point |
| 344 | ABP/PPG_D_Med_DN_wrtDPP | Median duration of the ABP/PPG cardiac cycle dicrotic notch with respect to diastolic phase peak |
| 345 | ABP/PPG_D_Med_DN_wrtDPE | Median duration of the ABP/PPG cardiac cycle dicrotic notch with respect to diastolic phase end point |
| 346 | ABP/PPG_D_Med_DPP_wrtDPE | Median duration of the ABP/PPG cardiac cycle diastolic phase peak with respect to diastolic phase end point |
| 347 | ABP/PPG_D_NMed_SPO_wrtSPP | Median duration of the normalized ABP/PPG cardiac cycle systolic phase onset with respect to systolic phase peak |
| 348 | ABP/PPG_D_NMed_SPO_wrtDN | Median duration of the normalized ABP/PPG cardiac cycle systolic phase onset with respect to dicrotic notch |
| 349 | ABP/PPG_D_NMed_SPO_wrtDPP | Median duration of the normalized ABP/PPG cardiac cycle systolic phase onset with respect to diastolic phase peak |
| 350 | ABP/PPG_D_NMED_SPO_wrtDPE | Median duration of the normalized ABP/PPG cardiac cycle systolic phase onset with respect to diastolic phase end point |
| 351 | ABP/PPG_D_NMed_SPP_wrtDN | Median duration of the normalized ABP/PPG cardiac cycle systolic phase peak with respect to dicrotic notch |
| 352 | ABP/PPG_D_NMed_SPP_wrtDPP | Median duration of the normalized ABP/PPG cardiac cycle systolic phase peak with respect to diastolic phase peak |
| 353 | ABP/PPG_D_NMed_SPP_wrtDPE | Median duration of the normalized ABP/PPG cardiac cycle systolic phase peak with respect to diastolic phase end point |
| 354 | ABP/PPG_D_NMed_DN_wrtDPP | Median duration of the normalized ABP/PPG cardiac cycle dicrotic notch with respect to diastolic phase peak |
| 355 | ABP/PPG_D_NMed_DN_wrtDPE | Median duration of the normalized ABP/PPG cardiac cycle dicrotic notch with respect to diastolic phase end point |
| 356 | ABP/PPG_D_NMed_DPP_wrtDPE | Median duration of the normalized ABP/PPG cardiac cycle diastolic phase peak with respect to diastolic phase end point |

11. **Root mean square features (357-376)**

| 357 | ABP/PPG_D_RMS_SPO_wrtSPP | Root mean square duration of the ABP/PPG cardiac cycle systolic phase onset with respect to systolic phase peak |
| --- | --- | --- |
| 358 | ABP/PPG_D_RMS_SPO_wrtDN | Root mean square duration of the ABP/PPG cardiac cycle systolic phase onset with respect to dicrotic notch |
| 359 | ABP/PPG_D_RMS_SPO_wrtDPP | Root mean square duration of the ABP/PPG cardiac cycle systolic phase onset with respect to diastolic phase peak |
| 360 | ABP/PPG_D_RMS_SPO_wrtDPE | Root mean square duration of the ABP/PPG cardiac cycle systolic phase onset with respect to diastolic phase end point |
| 361 | ABP/PPG_D_RMS_SPP_wrtDN | Root mean square duration of the ABP/PPG cardiac cycle systolic phase peak with respect to dicrotic notch |
| 362 | ABP/PPG_D_RMS_ SPP_wrtDPP | Root mean square duration of the ABP/PPG cardiac cycle systolic phase peak with respect to diastolic phase peak |
| 363 | ABP/PPG_D_RMS_ SPP_wrtDPE | Root mean square duration of the ABP/PPG cardiac cycle systolic phase peak with respect to diastolic phase end point |
| 364 | ABP/PPG_D_RMS_ DN_wrtDPP | Root mean square duration of the ABP/PPG cardiac cycle dicrotic notch with respect to diastolic phase peak |
| 365 | ABP/PPG_D_RMS_ DN_wrtDPE | Root mean square duration of the ABP/PPG cardiac cycle dicrotic notch with respect to diastolic phase end point |
| 366 | ABP/PPG_D_RMS_ DPP_wrtDPE | Root mean square duration of the ABP/PPG cardiac cycle diastolic phase peak with respect to diastolic phase end point |
| 367 | ABP/PPG_D_NRMS_SPO_wrtSPP | Root mean square duration of the normalized ABP/PPG cardiac cycle systolic phase onset with respect to systolic phase peak |
| 368 | ABP/PPG_D_NRMS_SPO_wrtDN | Root mean square duration of the normalized ABP/PPG cardiac cycle systolic phase onset with respect to dicrotic notch |
| 369 | ABP/PPG_D_NRMS_SPO_wrtDPP | Root mean square duration of the normalized ABP/PPG cardiac cycle systolic phase onset with respect to diastolic phase peak |
| 370 | ABP/PPG_D_NRMS_SPO_wrtDPE | Root mean square duration of the normalized ABP/PPG cardiac cycle systolic phase onset with respect to diastolic phase end point |
| 371 | ABP/PPG_D_NRMS_SPP_wrtDN | Root mean square duration of the normalized ABP/PPG cardiac cycle systolic phase peak with respect of dicrotic notch |
| 372 | ABP/PPG_D_NRMS_ SPP _wrtDPP | Root mean square duration of the normalized ABP/PPG cardiac cycle systolic phase peak with respect to diastolic phase peak |
| 373 | ABP/PPG_D_NRMS_ SPP_wrtDPE | Root mean square duration of the normalized ABP/PPG cardiac cycle systolic phase peak with respect to diastolic phase end point |
| 374 | ABP/PPG_D_NRMS_ DN_wrtDPP | Root mean square duration of the normalized ABP/PPG cardiac cycle dicrotic notch with respect to diastolic phase peak |
| 375 | ABP/PPG_D_NRMS_ DN_wrtDPE | Root mean square duration of the normalized ABP/PPG cardiac cycle dicrotic notch with respect to diastolic phase end point |
| 376 | ABP/PPG_D_NRMS_ DPP_wrtDPE | Root mean square duration of the normalized ABP/PPG cardiac cycle diastolic phase peak with respect to diastolic phase end point |

12. **Percentage features (377-412)**

| 377 | ABP/PPG_W_SRP10 | ABP/PPG cardiac cycle systolic rise phase width at 10% of the cardiac cycle |
| --- | --- | --- |
| 378 | ABP/PPG_W_ODP10 | ABP/PPG cardiac cycle overall decay phase width at 10% of the cardiac cycle |
| 379 | ABP/PPG_W_SRP10_ODP10 | Sum of the ABP/PPG cardiac cycle systolic rise phase width and overall decay phase width at 10% of the cardiac cycle |
| 380 | ABP/PPG_RT_ODP10_W_SRP10_W | The ratio of the ABP/PPG cardiac cycle overall decay phase width at 10% of the cardiac cycle to the ABP/PPG cardiac cycle systolic rise phase width at 10% of the cardiac cycle |
| 381 | ABP/PPG_RT_ODP10_W_SRP10_ODP10_W | The ratio of the ABP/PPG cardiac cycle overall decay phase width at 10% of the cardiac cycle to the sum of the ABP/PPG cardiac cycle systolic rise phase width and overall decay phase width at 10% of the cardiac cycle |
| 382 | ABP/PPG_RT_SRP10_W_SRP10_ODP10_W | The ratio of the ABP/PPG cardiac cycle systolic rise phase width at 10% of the cardiac cycle to the sum of the ABP/PPG cardiac cycle systolic rise phase width and overall decay phase width at 10% of the cardiac cycle |
| 383 | ABP/PPG_W_SRP25 | ABP/PPG cardiac cycle systolic rise phase width at 25% of the cardiac cycle |
| 384 | ABP/PPG_W_ODP25 | ABP/PPG cardiac cycle overall decay phase width at 25% of the cardiac cycle |
| 385 | ABP/PPG_W_SRP25_ODP25 | Sum of the ABP/PPG cardiac cycle systolic rise phase width and overall decay phase width at 25% of the cardiac cycle |
| 386 | ABP/PPG_RT_ODP25_W_SRP25_W | The ratio of the ABP/PPG cardiac cycle overall decay phase width at 25% of the cardiac cycle to the ABP/PPG cardiac cycle systolic rise phase width at 25% of the cardiac cycle |
| 387 | ABP/PPG_RT_ODP25_W_SRP25_ODP25_W | The ratio of the ABP/PPG cardiac cycle overall decay phase width at 25% of the cardiac cycle to the sum of the ABP/PPG cardiac cycle systolic rise phase width and overall decay phase width at 25% of the cardiac cycle |
| 388 | ABP/PPG_RT_SRP25_W_SRP25_ODP25_W | The ratio of the ABP/PPG cardiac cycle systolic rise phase width at 25% of the cardiac cycle to the sum of the ABP/PPG cardiac cycle systolic rise phase width and overall decay phase width at 25% of the cardiac cycle |
| 389 | ABP/PPG_W_SRP33 | ABP/PPG cardiac cycle systolic rise phase width at 33% of the cardiac cycle |
| 390 | ABP/PPG_W_ODP33 | ABP/PPG cardiac cycle overall decay phase width at 33% of the cardiac cycle |
| 391 | ABP/PPG_W_SRP33_ODP33 | Sum of the ABP/PPG cardiac cycle systolic rise phase width and overall decay phase width at 33% of the cardiac cycle |
| 392 | ABP/PPG_RT_ODP33_W_SRP33_W | The ratio of the ABP/PPG cardiac cycle overall decay phase width at 33% of the cardiac cycle to the ABP/PPG cardiac cycle systolic rise phase width at 33% of the cardiac cycle |
| 393 | ABP/PPG_RT_ODP33_W_SRP33_ODP33_W | The ratio of the ABP/PPG cardiac cycle overall decay phase width at 33% of the cardiac cycle to the sum of the ABP/PPG cardiac cycle systolic rise phase width and overall decay phase width at 33% of the cardiac cycle |
| 394 | ABP/PPG_RT_SRP33_W_SRP33_ODP33_W | The ratio of the ABP/PPG cardiac cycle systolic rise phase width at 33% of the cardiac cycle to the sum of the ABP/PPG cardiac cycle systolic rise phase width and overall decay phase width at 33% of the cardiac cycle |
| 395 | ABP/PPG_W_SRP50 | ABP/PPG cardiac cycle systolic rise phase width at 50% of the cardiac cycle |
| 396 | ABP/PPG_W_ODP50 | ABP/PPG cardiac cycle overall decay phase width at 50% of the cardiac cycle |
| 397 | ABP/PPG_W_SRP50_ODP50 | Sum of the ABP/PPG cardiac cycle systolic rise phase width and overall decay phase width at 50% of the cardiac cycle |
| 398 | ABP/PPG_RT_ODP50_W_SRP50_W | The ratio of the ABP/PPG cardiac cycle overall decay phase width at 50% of the cardiac cycle to the ABP/PPG cardiac cycle systolic rise phase width at 50% of the cardiac cycle |
| 399 | ABP/PPG_RT_ODP50_W_SRP50_ODP50_W | The ratio of the ABP/PPG cardiac cycle overall decay phase width at 50% of the cardiac cycle to the sum of the ABP/PPG cardiac cycle systolic rise phase width and overall decay phase width at 50% of the cardiac cycle |
| 400 | ABP/PPG_RT_SRP50_W_SRP50_ODP50_W | The ratio of the ABP/PPG cardiac cycle systolic rise phase width at 50% of the cardiac cycle to the sum of the ABP/PPG cardiac cycle systolic rise phase width and overall decay phase width at 50% of the cardiac cycle |
| 401 | ABP/PPG_W_SRP66 | ABP/PPG cardiac cycle systolic rise phase width at 66% of the cardiac cycle |
| 402 | ABP/PPG_W_ODP66 | ABP/PPG cardiac cycle overall decay phase width at 66% of the cardiac cycle |
| 403 | ABP/PPG_W_SRP66_ODP66 | Sum of the ABP/PPG cardiac cycle systolic rise phase width and overall decay phase width at 66% of the cardiac cycle |
| 404 | ABP/PPG_RT_ODP66_W_SRP66_W | The ratio of the ABP/PPG cardiac cycle overall decay phase width at 66% of the cardiac cycle to the ABP/PPG cardiac cycle systolic rise phase width at 66% of the cardiac cycle |
| 405 | ABP/PPG_RT_ODP66_W_SRP66_ODP66_W | The ratio of the ABP/PPG cardiac cycle overall decay phase width at 66% of the cardiac cycle to the sum of the ABP/PPG cardiac cycle systolic rise phase width and overall decay phase width at 66% of the cardiac cycle |
| 406 | ABP/PPG_RT_SRP66_W_SRP66_ODP66_W | The ratio of the ABP/PPG cardiac cycle systolic rise phase width at 66% of the cardiac cycle to the sum of the ABP/PPG cardiac cycle systolic rise phase width and overall decay phase width at 66% of the cardiac cycle |
| 407 | ABP/PPG_W_SRP75 | ABP/PPG cardiac cycle systolic rise phase width at 75% of the cardiac cycle |
| 408 | ABP/PPG_W_ODP75 | ABP/PPG cardiac cycle overall decay phase width at 75% of the cardiac cycle |
| 409 | ABP/PPG_W_SRP75_ODP75 | Sum of the ABP/PPG cardiac cycle systolic rise phase width and overall decay phase width at 75% of the cardiac cycle |
| 410 | ABP/PPG_RT_ODP75_W_SRP75_W | The ratio of the ABP/PPG cardiac cycle overall decay phase width at 75% of the cardiac cycle to the ABP/PPG cardiac cycle systolic rise phase width at 25% of the cardiac cycle |
| 411 | ABP/PPG_RT_ODP75_W_SRP75_ODP75_W | The ratio of the ABP/PPG cardiac cycle overall decay phase width at 75% of the cardiac cycle to the sum of the ABP/PPG cardiac cycle systolic rise phase width and overall decay phase width at 75% of the cardiac cycle |
| 412 | ABP/PPG_RT_SRP75_W_SRP75_ODP75_W | The ratio of the ABP/PPG cardiac cycle systolic rise phase width at 75% of the cardiac cycle to the sum of the ABP/PPG cardiac cycle systolic rise phase width and overall decay phase width at 75% of the cardiac cycle |

13. **Area features (413-422)**

| 413 | ABP/PPG_D_Are_SPO_wrtSPP | Area under the ABP/PPG cardiac cycle systolic phase onset with respect to systolic phase peak |
| --- | --- | --- |
| 414 | ABP/PPG_D_Are_SPO_wrtDN | Area under the ABP/PPG cardiac cycle systolic phase onset with respect to dicrotic notch |
| 415 | ABP/PPG_D_Are_SPO_wrtDPP | Area under the ABP/PPG cardiac cycle systolic phase onset with respect to diastolic phase peak |
| 416 | ABP/PPG_D_Are_SPO_wrtDPE | Area under the ABP/PPG cardiac cycle systolic phase onset with respect to diastolic phase end point |
| 417 | ABP/PPG_D_Are_SPP_wrtDN | Area under the ABP/PPG cardiac cycle systolic phase peak with respect to dicrotic notch |
| 418 | ABP/PPG_D_Are_SPP_wrtDPP | Area under the ABP/PPG cardiac cycle systolic phase peak with respect to diastolic phase peak |
| 419 | ABP/PPG_D_Are_SPP_wrtDPE | Area under the ABP/PPG cardiac cycle systolic phase peak with respect to diastolic phase end point |
| 420 | ABP/PPG_D_Are_DN_wrtDPP | Area under the ABP/PPG cardiac cycle dicrotic notch with respect to diastolic phase peak |
| 421 | ABP/PPG_D_Are_DN_wrtDPE | Area under the ABP/PPG cardiac cycle dicrotic notch with respect to diastolic phase end point |
| 422 | ABP/PPG_D_Are_DPP_wrtDPE | Area under the ABP/PPG cardiac cycle diastolic phase peak with respect to diastolic phase end point |

14. **Area features with subtracted diastolic end point (423-432)**

| 423 | ABP/PPG_D_Are_wsDPE_SPO_wrtSPP | Area under the ABP/PPG cardiac cycle systolic phase onset with respect to systolic phase peak, with subtracted diastolic phase end point |
| --- | --- | --- |
| 424 | ABP/PPG_D_Are_wsDPE_SPO_wrtDN | Area under the ABP/PPG cardiac cycle systolic phase onset with respect to dicrotic notch, with subtracted diastolic phase end point |
| 425 | ABP/PPG_D_Are_wsDPE_SPO_wrtDPP | Area under the ABP/PPG cardiac cycle systolic phase onset with respect to diastolic phase peak, with subtracted diastolic phase end point |
| 426 | ABP/PPG_D_Are_wsDPE_SPO_wrtDPE | Area under the ABP/PPG cardiac cycle systolic phase onset with respect to diastolic phase end point, with subtracted diastolic phase end point |
| 427 | ABP/PPG_D_Are_wsDPE_SPP_wrtDN | Area under the ABP/PPG cardiac cycle systolic phase peak with respect to dicrotic notch, with subtracted diastolic phase end point |
| 428 | ABP/PPG_D_Are_wsDPE_SPP_wrtDPP | Area under the ABP/PPG cardiac cycle systolic phase peak with respect to diastolic phase peak, with subtracted diastolic phase end point |
| 429 | ABP/PPG_D_Are_wsDPE_SPP_wrtDPE | Area under the ABP/PPG cardiac cycle systolic phase peak with respect to diastolic phase end point, with subtracted diastolic phase end point |
| 430 | ABP/PPG_D_Are_wsDPE_DN_wrtDPP | Area under the ABP/PPG cardiac cycle dicrotic notch with respect to diastolic phase peak, with subtracted diastolic phase end point |
| 431 | ABP/PPG_D_Are_wsDPE_DN_wrtDPE | Area under the ABP/PPG cardiac cycle dicrotic notch with respect to diastolic phase end point, with subtracted diastolic phase end point |
| 432 | ABP/PPG_D_Are_wsDPE_DPP_wrtDPE | Area under the ABP/PPG cardiac cycle diastolic phase peak with respect to diastolic phase end point, with subtracted diastolic phase end point |

15. **Normalized area features (433-442)**

| 433 | ABP/PPG_D_NAre_SPO_wrtSPP | Area under the normalized ABP/PPG cardiac cycle systolic phase onset with respect to systolic phase peak |
| --- | --- | --- |
| 434 | ABP/PPG_D_NAre_SPO_wrtDN | Area under the normalized ABP/PPG cardiac cycle systolic phase onset with respect to dicrotic notch |
| 435 | ABP/PPG_D_NAre_SPO_wrtDPP | Area under the normalized ABP/PPG cardiac cycle systolic phase onset with respect to diastolic phase peak |
| 436 | ABP/PPG_D_NAre_SPO_wrtDPE | Area under the normalized ABP/PPG cardiac cycle systolic phase onset with respect to diastolic phase end point |
| 437 | ABP/PPG_D_NAre_SPP_wrtDN | Area under the normalized ABP/PPG cardiac cycle systolic phase peak with respect to dicrotic notch |
| 438 | ABP/PPG_D_NAre_SPP_wrtDPP | Area under the normalized ABP/PPG cardiac cycle systolic phase peak with respect to diastolic phase peak |
| 439 | ABP/PPG_D_NAre_SPP_wrtDPE | Area under the normalized ABP/PPG cardiac cycle systolic phase peak with respect to diastolic phase end point |
| 440 | ABP/PPG_D_NAre_DN_wrtDPP | Area under the normalized ABP/PPG cardiac cycle dicrotic notch with respect to diastolic phase peak |
| 441 | ABP/PPG_D_NAre_DN_wrtDPE | Area under the normalized ABP/PPG cardiac cycle dicrotic notch with respect to diastolic phase end point |
| 442 | ABP/PPG_D_NAre_DPP_wrtDPE | Area under the normalized ABP/PPG cardiac cycle diastolic phase peak with respect to diastolic phase end point |

16. **Normalized area features with subtracted end point (443-452)**

| 443 | ABP/PPG_D_NAre_wsDPE_SPO_wrtSPP | Area under the normalized ABP/PPG cardiac cycle systolic phase onset with respect to systolic phase peak, with subtracted diastolic phase end point |
| --- | --- | --- |
| 444 | ABP/PPG_D_NAre_wsDPE_SPO_wrtDN | Area under the normalized ABP/PPG cardiac cycle systolic phase onset with respect to dicrotic notch, with subtracted diastolic phase end point |
| 445 | ABP/PPG_D_NAre_wsDPE_SPO_wrtDPP | Area under the normalized ABP/PPG cardiac cycle systolic phase onset with respect to diastolic phase peak, with subtracted diastolic phase end point |
| 446 | ABP/PPG_D_NAre_wsDPE_SPO_wrtDPE | Area under the normalized ABP/PPG cardiac cycle systolic phase onset with respect to diastolic phase end point, with subtracted diastolic phase end point |
| 447 | ABP/PPG_D_NAre_wsDPE_SPP_wrtDN | Area under the normalized ABP/PPG cardiac cycle systolic phase peak with respect to dicrotic notch, with subtracted diastolic phase end point |
| 448 | ABP/PPG_D_NAre_wsDPE_SPP_wrtDPP | Area under the normalized ABP/PPG cardiac cycle systolic phase peak with respect to diastolic phase peak, with subtracted diastolic phase end point |
| 449 | ABP/PPG_D_NAre_wsDPE_SPP_wrtDPE | Area under the normalized ABP/PPG cardiac cycle systolic phase peak with respect to diastolic phase end point, with subtracted diastolic phase end point |
| 450 | ABP/PPG_D_NAre_wsDPE_DN_wrtDPP | Area under the normalized ABP/PPG cardiac cycle dicrotic notch with respect to diastolic phase peak, with subtracted diastolic phase end point |
| 451 | ABP/PPG_D_NAre_wsDPE_DN_wrtDPE | Area under the normalized ABP/PPG cardiac cycle dicrotic notch with respect to diastolic phase end point, with subtracted diastolic phase end point |
| 452 | ABP/PPG_D_NAre_wsDPE_DPP_wrtDPE | Area under the normalized ABP/PPG cardiac cycle diastolic phase peak with respect to diastolic phase end point, with subtracted diastolic phase end point |

17. **Area ratio features (453-632)**

| 453 | ABP/PPG_D_RT_Are_SPO_wrtDN_ABP/PPG_D_Are_SPO_wrtSPP | The ratio of the area under the ABP/PPG cardiac cycle systolic phase onset with respect to dicrotic notch to the area under the ABP/PPG cardiac cycle systolic phase onset with respect to systolic phase peak |
| --- | --- | --- |
| 454 | ABP/PPG_D_RT_Are_SPO_wrtDPP_ABP/PPG_D_Are_SPO_wrtSPP | The ratio of the area under the ABP/PPG cardiac cycle systolic phase onset with respect to diastolic phase peak to the area under the ABP/PPG cardiac cycle systolic phase onset with respect to systolic phase peak |
| 455 | ABP/PPG_D_RT_Are_SPO_wrtDPE_ABP/PPG_D_Are_SPO_wrtSPP | The ratio of the area under the ABP/PPG cardiac cycle systolic phase onset with respect to diastolic phase end point to the area under the ABP/PPG cardiac cycle systolic phase onset with respect to systolic phase peak |
| 456 | ABP/PPG_D_RT_Are_SPP_wrtDN_ABP/PPG_D_Are_SPO_wrtSPP | The ratio of the area under the ABP/PPG cardiac cycle systolic phase peak with respect to dicrotic notch to the area under the ABP/PPG cardiac cycle systolic phase onset with respect to systolic phase peak |
| 457 | ABP/PPG_D_RT_Are_SPP_wrtDPP_ABP/PPG_D_Are_SPO_wrtSPP | The ratio of the area under the ABP/PPG cardiac cycle systolic phase peak with respect to diastolic phase peak to the area under the ABP/PPG cardiac cycle systolic phase onset with respect to systolic phase peak |
| 458 | ABP/PPG_D_RT_Are_SPP_wrtDPE_ABP/PPG_D_Are_SPO_wrtSPP | The ratio of the area under the ABP/PPG cardiac cycle systolic phase peak with respect to diastolic phase end point to the area under the ABP/PPG cardiac cycle systolic phase onset with respect to systolic phase peak |
| 459 | ABP/PPG_D_RT_Are_DN_wrtDPP_ABP/PPG_D_Are_SPO_wrtSPP | The ratio of the area under the ABP/PPG cardiac cycle dicrotic notch with respect to diastolic phase peak to the area under the ABP/PPG cardiac cycle systolic phase onset with respect to systolic phase peak |
| 460 | ABP/PPG_D_RT_Are_DN_wrtDPE_ABP/PPG_D_Are_SPO_wrtSPP | The ratio of the area under the ABP/PPG cardiac cycle dicrotic notch with respect to diastolic phase end point to the area under the ABP/PPG cardiac cycle systolic phase onset with respect to systolic phase peak |
| 461 | ABP/PPG_D_RT_Are_DPP_wrtDPE_ABP/PPG_D_Are_SPO_wrtSPP | The ratio of the area under the ABP/PPG cardiac cycle diastolic phase peak with respect to diastolic phase end point to the area under the ABP/PPG cardiac cycle systolic phase onset with respect to systolic phase peak |
| 462 | ABP/PPG_D_RT_Are_SPO_wrtDPP_ABP/PPG_D_Are_SPO_wrtDN | The ratio of the area under the ABP/PPG cardiac cycle systolic phase onset with respect to diastolic phase peak to the area under the ABP/PPG cardiac cycle systolic phase onset with respect to dicrotic notch |
| 463 | ABP/PPG_D_RT_Are_SPO_wrtDPE_ABP/PPG_D_Are_SPO_wrtDN | The ratio of the area under the ABP/PPG cardiac cycle systolic phase onset with respect to diastolic phase end point to the area under the ABP/PPG cardiac cycle systolic phase onset with respect to dicrotic notch |
| 464 | ABP/PPG_D_RT_Are_SPP_wrtDN_ABP/PPG_D_Are_SPO_wrtDN | The ratio of the area under the ABP/PPG cardiac cycle systolic phase peak with respect to dicrotic notch to the area under the ABP/PPG cardiac cycle systolic phase onset with respect to dicrotic notch |
| 465 | ABP/PPG_D_RT_Are_SPP_wrtDPP_ABP/PPG_D_Are_SPO_wrtDN | The ratio of the area under the ABP/PPG cardiac cycle systolic phase peak with respect to diastolic phase peak to the area under the ABP/PPG cardiac cycle systolic phase onset with respect to dicrotic notch |
| 466 | ABP/PPG_D_RT_Are_SPP_wrtDPE_ABP/PPG_D_Are_SPO_wrtDN | The ratio of the area under the ABP/PPG cardiac cycle systolic phase peak with respect to diastolic phase end point to the area under the ABP/PPG cardiac cycle systolic phase onset with respect to dicrotic notch |
| 467 | ABP/PPG_D_RT_Are_DN_wrtDPP_ABP/PPG_D_Are_SPO_wrtDN | The ratio of the area under the ABP/PPG cardiac cycle dicrotic notch with respect to diastolic phase peak to the area under the ABP/PPG cardiac cycle systolic phase onset with respect to dicrotic notch |
| 468 | ABP/PPG_D_RT_Are_DN_wrtDPE_ABP/PPG_D_Are_SPO_wrtDN | The ratio of the area under the ABP/PPG cardiac cycle dicrotic notch with respect to diastolic phase end point to the area under the ABP/PPG cardiac cycle systolic phase onset with respect to dicrotic notch |
| 469 | ABP/PPG_D_RT_Are_DPP_wrtDPE_ABP/PPG_D_Are_SPO_wrtDN | The ratio of the area under the ABP/PPG cardiac cycle diastolic phase peak with respect to diastolic phase end point to the area under the ABP/PPG cardiac cycle systolic phase onset with respect to dicrotic notch |
| 470 | ABP/PPG_D_RT_Are_SPO_wrtDPE_ABP/PPG_D_Are_SPO_wrtDPP | The ratio of the area under the ABP/PPG cardiac cycle systolic phase onset with respect to diastolic phase end point to the area under the ABP/PPG cardiac cycle systolic phase onset with respect to diastolic phase peak |
| 471 | ABP/PPG_D_RT_Are_SPP_wrtDN_ABP/PPG_D_Are_SPO_wrtDPP | The ratio of the area under the ABP/PPG cardiac cycle systolic phase peak with respect to dicrotic notch to the area under the ABP/PPG cardiac cycle systolic phase onset with respect to diastolic phase peak |
| 472 | ABP/PPG_D_RT_Are_SPP_wrtDPP_ABP/PPG_D_Are_SPO_wrtDPP | The ratio of the area under the ABP/PPG cardiac cycle systolic phase peak with respect to diastolic phase peak to the area under the ABP/PPG cardiac cycle systolic phase onset with respect to diastolic phase peak |
| 473 | ABP/PPG_D_RT_Are_SPP_wrtDPE_ABP/PPG_D_Are_SPO_wrtDPP | The ratio of the area under the ABP/PPG cardiac cycle systolic phase peak with respect to diastolic phase end point to the area under the ABP/PPG cardiac cycle systolic phase onset with respect to diastolic phase peak |
| 474 | ABP/PPG_D_RT_Are_DN_wrtDPP_ABP/PPG_D_Are_SPO_wrtDPP | The ratio of the area under the ABP/PPG cardiac cycle dicrotic notch with respect to diastolic phase peak to the area under the ABP/PPG cardiac cycle systolic phase onset with respect to diastolic phase peak |
| 475 | ABP/PPG_D_RT_Are_DN_wrtDPE_ABP/PPG_D_Are_SPO_wrtDPP | The ratio of the area under the ABP/PPG cardiac cycle dicrotic notch with respect to diastolic phase end point to the area under the ABP/PPG cardiac cycle systolic phase onset with respect to diastolic phase peak |
| 476 | ABP/PPG_D_RT_Are_DPP_wrtDPE_ABP/PPG_D_Are_SPO_wrtDPP | The ratio of the area under the ABP/PPG cardiac cycle diastolic phase peak with respect to diastolic phase end point to the area under the ABP/PPG cardiac cycle systolic phase onset with respect to diastolic phase peak |
| 477 | ABP/PPG_D_RT_Are_SPP_wrtDN_ABP/PPG_D_Are_SPO_wrtDPE | The ratio of the area under the ABP/PPG cardiac cycle systolic phase peak with respect to dicrotic notch to the area under the ABP/PPG cardiac cycle systolic phase onset with respect to diastolic phase end point |
| 478 | ABP/PPG_D_RT_Are_SPP_wrtDPP_ABP/PPG_D_Are_SPO_wrtDPE | The ratio of the area under the ABP/PPG cardiac cycle systolic phase peak with respect to diastolic phase peak to the area under the ABP/PPG cardiac cycle systolic phase onset with respect to diastolic phase end point |
| 479 | ABP/PPG_D_RT_Are_SPP_wrtDPE_ABP/PPG_D_Are_SPO_wrtDPE | The ratio of the area under the ABP/PPG cardiac cycle systolic phase peak with respect to diastolic phase end point to the area under the ABP/PPG cardiac cycle systolic phase onset with respect to diastolic phase end point |
| 480 | ABP/PPG_D_RT_Are_DN_wrtDPP_ABP/PPG_D_Are_SPO_wrtDPE | The ratio of the area under the ABP/PPG cardiac cycle dicrotic notch with respect to diastolic phase peak to the area under the ABP/PPG cardiac cycle systolic phase onset with respect to diastolic phase end point |
| 481 | ABP/PPG_D_RT_Are_DN_wrtDPE_ABP/PPG_D_Are_SPO_wrtDPE | The ratio of the area under the ABP/PPG cardiac cycle dicrotic notch with respect to diastolic phase end point to the area under the ABP/PPG cardiac cycle systolic phase onset with respect to diastolic phase end point |
| 482 | ABP/PPG_D_RT_Are_DPP_wrtDPE_ABP/PPG_D_Are_SPO_wrtDPE | The ratio of the area under the ABP/PPG cardiac cycle diastolic phase peak with respect to diastolic phase end point to the area under the ABP/PPG cardiac cycle systolic phase onset with respect to diastolic phase end point |
| 483 | ABP/PPG_D_RT_Are_SPP_wrtDPP_ABP/PPG_D_Are_SPP_wrtDN | The ratio of the area under the ABP/PPG cardiac cycle systolic phase peak with respect to diastolic phase peak to the area under the ABP/PPG cardiac cycle systolic phase peak with respect to dicrotic notch |
| 484 | ABP/PPG_D_RT_Are_SPP_wrtDPE_ABP/PPG_D_Are_SPP_wrtDN | The ratio of the area under the ABP/PPG cardiac cycle systolic phase peak with respect to diastolic phase end point to the area under the ABP/PPG cardiac cycle systolic phase peak with respect to dicrotic notch |
| 485 | ABP/PPG_D_RT_Are_DN_wrtDPP_ABP/PPG_D_Are_SPP_wrtDN | The ratio of the area under the ABP/PPG cardiac cycle dicrotic notch with respect to diastolic phase peak to the area under the ABP/PPG cardiac cycle systolic phase peak with respect to dicrotic notch |
| 486 | ABP/PPG_D_RT_Are_DN_wrtDPE_ABP/PPG_D_Are_SPP_wrtDN | The ratio of the area under the ABP/PPG cardiac cycle dicrotic notch with respect to diastolic phase end point to the area under the ABP/PPG cardiac cycle systolic phase peak with respect to dicrotic notch |
| 487 | ABP/PPG_D_RT_Are_DPP_wrtDPE_ABP/PPG_D_Are_SPP_wrtDN | The ratio of the area under the ABP/PPG cardiac cycle diastolic phase peak with respect to diastolic phase end point to the area under the ABP/PPG cardiac cycle systolic phase peak with respect to dicrotic notch |
| 488 | ABP/PPG_D_RT_Are_SPP_wrtDPE_ABP/PPG_D_Are_SPP_wrtDPP | The ratio of the area under the ABP/PPG cardiac cycle systolic phase peak with respect to diastolic phase end point to the area under the ABP/PPG cardiac cycle systolic phase peak with respect to diastolic phase peak |
| 489 | ABP/PPG_D_RT_Are_DN_wrtDPP_ABP/PPG_D_Are_SPP_wrtDPP | The ratio of the area under the ABP/PPG cardiac cycle dicrotic notch with respect to diastolic phase peak to the area under the ABP/PPG cardiac cycle systolic phase peak with respect to diastolic phase peak |
| 490 | ABP/PPG_D_RT_Are_DN_wrtDPE_ABP/PPG_D_Are_SPP_wrtDPP | The ratio of the area under the ABP/PPG cardiac cycle dicrotic notch with respect to diastolic phase end point to the area under the ABP/PPG cardiac cycle systolic phase peak with respect to diastolic phase peak |
| 491 | ABP/PPG_D_RT_Are_DPP_wrtDPE_ABP/PPG_D_Are_SPP_wrtDPP | The ratio of the area under the ABP/PPG cardiac cycle diastolic phase peak with respect to diastolic phase end point to the area under the ABP/PPG cardiac cycle systolic phase peak with respect to diastolic phase peak |
| 492 | ABP/PPG_D_RT_Are_DN_wrtDPP_ABP/PPG_D_Are_SPP_wrtDPE | The ratio of the area under the ABP/PPG cardiac cycle dicrotic notch with respect to diastolic phase peak to the area under the ABP/PPG cardiac cycle systolic phase peak with respect to diastolic phase end point |
| 493 | ABP/PPG_D_RT_Are_DN_wrtDPE_ABP/PPG_D_Are_SPP_wrtDPE | The ratio of the area under the ABP/PPG cardiac cycle dicrotic notch with respect to diastolic phase end point to the area under the ABP/PPG cardiac cycle systolic phase peak with respect to diastolic phase end point |
| 494 | ABP/PPG_D_RT_Are_DPP_wrtDPE_ABP/PPG_D_Are_SPP_wrtDPE | The ratio of the area under the ABP/PPG cardiac cycle diastolic phase peak with respect to diastolic phase end point to the area under the ABP/PPG cardiac cycle systolic phase peak with respect to diastolic phase end point |
| 495 | ABP/PPG_D_RT_Are_DN_wrtDPE_ABP/PPG_D_Are_DN_wrtDPP | The ratio of the area under the ABP/PPG cardiac cycle dicrotic notch with respect to diastolic phase end point to the area under the ABP/PPG cardiac cycle dicrotic notch with respect to diastolic phase peak |
| 496 | ABP/PPG_D_RT_Are_DPP_wrtDPE_ABP/PPG_D_Are_DN_wrtDPP | The ratio of the area under the ABP/PPG cardiac cycle diastolic phase peak with respect to diastolic phase end point to the area under the ABP/PPG cardiac cycle dicrotic notch with respect to diastolic phase peak |
| 497 | ABP/PPG_D_RT_Are_DPP_wrtDPE_ABP/PPG_D_Are_DN_wrtDPE | The ratio of the area under the ABP/PPG cardiac cycle diastolic phase peak with respect to diastolic phase end point to the area under the ABP/PPG cardiac cycle dicrotic notch with respect to diastolic phase end point |
| 498 | ABP/PPG_D_RT_Are_wsDPE_SPO_wrtDN_ABP/PPG_D_Are_wsDPE_SPO_wrtSPP | The ratio of the area under the ABP/PPG cardiac cycle systolic phase onset with respect to dicrotic notch, with subtracted diastolic phase end point to the area under the ABP/PPG cardiac cycle systolic phase onset with respect to systolic phase peak, with subtracted diastolic phase end point |
| 499 | ABP/PPG_D_RT_Are_wsDPE_SPO_wrtDPP_ABP/PPG_D_Are_wsDPE_SPO_wrtSPP | The ratio of the area under the ABP/PPG cardiac cycle systolic phase onset with respect to diastolic phase peak, with subtracted diastolic phase end point to the area under the ABP/PPG cardiac cycle systolic phase onset with respect to systolic phase peak, with subtracted diastolic phase end point |
| 500 | ABP/PPG_D_RT_Are_wsDPE_SPO_wrtDPE_ABP/PPG_D_Are_wsDPE_SPO_wrtSPP | The ratio of the area under the ABP/PPG cardiac cycle systolic phase onset with respect to diastolic phase end point, with subtracted diastolic phase end point to the area under the ABP/PPG cardiac cycle systolic phase onset with respect to systolic phase peak, with subtracted diastolic phase end point |
| 501 | ABP/PPG_D_RT_Are_wsDPE_SPP_wrtDN_ABP/PPG_D_Are_wsDPE_SPO_wrtSPP | The ratio of the area under the ABP/PPG cardiac cycle systolic phase peak with respect to dicrotic notch, with subtracted diastolic phase end point to the area under the ABP/PPG cardiac cycle systolic phase onset with respect to systolic phase peak, with subtracted diastolic phase end point |
| 502 | ABP/PPG_D_RT_Are_wsDPE_SPP_wrtDPP_ABP/PPG_D_Are_wsDPE_SPO_wrtSPP | The ratio of the area under the ABP/PPG cardiac cycle systolic phase peak with respect to diastolic phase peak, with subtracted diastolic phase end point to the area under the ABP/PPG cardiac cycle systolic phase onset with respect to systolic phase peak, with subtracted diastolic phase end point |
| 503 | ABP/PPG_D_RT_Are_wsDPE_SPP_wrtDPE_ABP/PPG_D_Are_wsDPE_SPO_wrtSPP | The ratio of the area under the ABP/PPG cardiac cycle systolic phase peak with respect to diastolic phase end point, with subtracted diastolic phase end point to the area under the ABP/PPG cardiac cycle systolic phase onset with respect to systolic phase peak, with subtracted diastolic phase end point |
| 504 | ABP/PPG_D_RT_Are_wsDPE_DN_wrtDPP_ABP/PPG_D_Are_wsDPE_SPO_wrtSPP | The ratio of the area under the ABP/PPG cardiac cycle dicrotic notch with respect to diastolic phase peak, with subtracted diastolic phase end point to the area under the ABP/PPG cardiac cycle systolic phase onset with respect to systolic phase peak, with subtracted diastolic phase end point |
| 505 | ABP/PPG_D_RT_Are_wsDPE_DN_wrtDPE_ABP/PPG_D_Are_wsDPE_SPO_wrtSPP | The ratio of the area under the ABP/PPG cardiac cycle dicrotic notch with respect to diastolic phase end point, with subtracted diastolic phase end point to the area under the ABP/PPG cardiac cycle systolic phase onset with respect to systolic phase peak, with subtracted diastolic phase end point |
| 506 | ABP/PPG_D_RT_Are_wsDPE_DPP_wrtDPE_ABP/PPG_D_Are_wsDPE_SPO_wrtSPP | The ratio of the area under the ABP/PPG cardiac cycle diastolic phase peak with respect to diastolic phase end point, with subtracted diastolic phase end point to the area under the ABP/PPG cardiac cycle systolic phase onset with respect to systolic phase peak, with subtracted diastolic phase end point |
| 507 | ABP/PPG_D_RT_Are_wsDPE_SPO_wrtDPP_ABP/PPG_D_Are_wsDPE_SPO_DN | The ratio of the area under the ABP/PPG cardiac cycle systolic phase onset with respect to diastolic phase peak, with subtracted diastolic phase end point to the area under the ABP/PPG cardiac cycle systolic phase onset with respect to dicrotic notch, with subtracted diastolic phase end point |
| 508 | ABP/PPG_D_RT_Are_wsDPE_SPO_wrtDPE_ABP/PPG_D_Are_wsDPE_SPO_DN | The ratio of the area under the ABP/PPG cardiac cycle systolic phase onset with respect to diastolic phase end point, with subtracted diastolic phase end point to the area under the ABP/PPG cardiac cycle systolic phase onset with respect to dicrotic notch, with subtracted diastolic phase end point |
| 509 | ABP/PPG_D_RT_Are_wsDPE_SPP_wrtDN_ABP/PPG_D_Are_wsDPE_SPO_DN | The ratio of the area under the ABP/PPG cardiac cycle systolic phase peak with respect to dicrotic notch, with subtracted diastolic phase end point to the area under the ABP/PPG cardiac cycle systolic phase onset with respect to dicrotic notch, with subtracted diastolic phase end point |
| 510 | ABP/PPG_D_RT_Are_wsDPE_SPP_wrtDPP_ABP/PPG_D_Are_wsDPE_SPO_DN | The ratio of the area under the ABP/PPG cardiac cycle systolic phase peak with respect to diastolic phase peak, with subtracted diastolic phase end point to the area under the ABP/PPG cardiac cycle systolic phase onset with respect to dicrotic notch, with subtracted diastolic phase end point |
| 511 | ABP/PPG_D_RT_Are_wsDPE_SPP_wrtDPE_ABP/PPG_D_Are_wsDPE_SPO_DN | The ratio of the area under the ABP/PPG cardiac cycle systolic phase peak with respect to diastolic phase end point, with subtracted diastolic phase end point to the area under the ABP/PPG cardiac cycle systolic phase onset with respect to dicrotic notch, with subtracted diastolic phase end point |
| 512 | ABP/PPG_D_RT_Are_wsDPE_DN_wrtDPP_ABP/PPG_D_Are_wsDPE_SPO_DN | The ratio of the area under the ABP/PPG cardiac cycle dicrotic notch with respect to diastolic phase peak, with subtracted diastolic phase end point to the area under the ABP/PPG cardiac cycle systolic phase onset with respect to dicrotic notch, with subtracted diastolic phase end point |
| 513 | ABP/PPG_D_RT_Are_wsDPE_DN_wrtDPE_ABP/PPG_D_Are_wsDPE_SPO_DN | The ratio of the area under the ABP/PPG cardiac cycle dicrotic notch with respect to diastolic phase end point, with subtracted diastolic phase end point to the area under the ABP/PPG cardiac cycle systolic phase onset with respect to dicrotic notch, with subtracted diastolic phase end point |
| 514 | ABP/PPG_D_RT_Are_wsDPE_DPP_wrtDPE_ABP/PPG_D_Are_wsDPE_SPO_DN | The ratio of the area under the ABP/PPG cardiac cycle diastolic phase peak with respect to diastolic phase end point, with subtracted diastolic phase end point to the area under the ABP/PPG cardiac cycle systolic phase onset with respect to dicrotic notch, with subtracted diastolic phase end point |
| 515 | ABP/PPG_D_RT_Are_wsDPE_SPO_wrtDPE_ABP/PPG_D_Are_wsDPE_SPO_wrtDPP | The ratio of the area under the ABP/PPG cardiac cycle systolic phase onset with respect to diastolic phase end point, with subtracted diastolic phase end point to the area under the ABP/PPG cardiac cycle systolic phase onset with respect to diastolic phase peak, with subtracted diastolic phase end point |
| 516 | ABP/PPG_D_RT_Are_wsDPE_SPP_wrtDN_ABP/PPG_D_Are_wsDPE_SPO_wrtDPP | The ratio of the area under the ABP/PPG cardiac cycle systolic phase peak with respect to dicrotic notch, with subtracted diastolic phase end point to the area under the ABP/PPG cardiac cycle systolic phase onset with respect to diastolic phase peak, with subtracted diastolic phase end point |
| 517 | ABP/PPG_D_RT_Are_wsDPE_SPP_wrtDPP_ABP/PPG_D_Are_wsDPE_SPO_wrtDPP | The ratio of the area under the ABP/PPG cardiac cycle systolic phase peak with respect to diastolic phase peak, with subtracted diastolic phase end point to the area under the ABP/PPG cardiac cycle systolic phase onset with respect to diastolic phase peak, with subtracted diastolic phase end point |
| 518 | ABP/PPG_D_RT_Are_wsDPE_SPP_wrtDPE_ABP/PPG_D_Are_wsDPE_SPO_wrtDPP | The ratio of the area under the ABP/PPG cardiac cycle systolic phase peak with respect to diastolic phase end point, with subtracted diastolic phase end point to the area under the ABP/PPG cardiac cycle systolic phase onset with respect to diastolic phase peak, with subtracted diastolic phase end point |
| 519 | ABP/PPG_D_RT_Are_wsDPE_DN_wrtDPP_ABP/PPG_D_Are_wsDPE_SPO_wrtDPP | The ratio of the area under the ABP/PPG cardiac cycle dicrotic notch with respect to diastolic phase peak, with subtracted diastolic phase end point to the area under the ABP/PPG cardiac cycle systolic phase onset with respect to diastolic phase peak, with subtracted diastolic phase end point |
| 520 | ABP/PPG_D_RT_Are_wsDPE_DN_wrtDPE_ABP/PPG_D_Are_wsDPE_SPO_wrtDPP | The ratio of the area under the ABP/PPG cardiac cycle dicrotic notch with respect to diastolic phase end point, with subtracted diastolic phase end point to the area under the ABP/PPG cardiac cycle systolic phase onset with respect to diastolic phase peak, with subtracted diastolic phase end point |
| 521 | ABP/PPG_D_RT_Are_wsDPE_DPP_wrtDPE_ABP/PPG_D_Are_wsDPE_SPO_wrtDPP | The ratio of the area under the ABP/PPG cardiac cycle diastolic phase peak with respect to diastolic phase end point, with subtracted diastolic phase end point to the area under the ABP/PPG cardiac cycle systolic phase onset with respect to diastolic phase peak, with subtracted diastolic phase end point |
| 522 | ABP/PPG_D_RT_Are_wsDPE_SPP_wrtDN_ABP/PPG_D_Are_wsDPE_SPO_wrtDPE | The ratio of the area under the ABP/PPG cardiac cycle systolic phase peak with respect to dicrotic notch, with subtracted diastolic phase end point to the area under the ABP/PPG cardiac cycle systolic phase onset with respect to diastolic phase end point, with subtracted diastolic phase end point |
| 523 | ABP/PPG_D_RT_Are_wsDPE_SPP_wrtDPP_ABP/PPG_D_Are_wsDPE_SPO_wrtDPE | The ratio of the area under the ABP/PPG cardiac cycle systolic phase peak with respect to diastolic phase peak, with subtracted diastolic phase end point to the area under the ABP/PPG cardiac cycle systolic phase onset with respect to diastolic phase end point, with subtracted diastolic phase end point |
| 524 | ABP/PPG_D_RT_Are_wsDPE_SPP_wrtDPE_ABP/PPG_D_Are_wsDPE_SPO_wrtDPE | The ratio of the area under the ABP/PPG cardiac cycle systolic phase peak with respect to diastolic phase end point, with subtracted diastolic phase end point to the area under the ABP/PPG cardiac cycle systolic phase onset with respect to diastolic phase end point, with subtracted diastolic phase end point |
| 525 | ABP/PPG_D_RT_Are_wsDPE_DN_wrtDPP_ABP/PPG_D_Are_wsDPE_SPO_wrtDPE | The ratio of the area under the ABP/PPG cardiac cycle dicrotic notch with respect to diastolic phase peak, with subtracted diastolic phase end point to the area under the ABP/PPG cardiac cycle systolic phase onset with respect to diastolic phase end point, with subtracted diastolic phase end point |
| 526 | ABP/PPG_D_RT_Are_wsDPE_DN_wrtDPE_ABP/PPG_D_Are_wsDPE_SPO_wrtDPE | The ratio of the area under the ABP/PPG cardiac cycle dicrotic notch with respect to diastolic phase end point, with subtracted diastolic phase end point to the area under the ABP/PPG cardiac cycle systolic phase onset with respect to diastolic phase end point, with subtracted diastolic phase end point |
| 527 | ABP/PPG_D_RT_Are_wsDPE_DPP_wrtDPE_ABP/PPG_D_Are_wsDPE_SPO_wrtDPE | The ratio of the area under the ABP/PPG cardiac cycle diastolic phase peak with respect to diastolic phase end point, with subtracted diastolic phase end point to the area under the ABP/PPG cardiac cycle systolic phase onset with respect to diastolic phase end point, with subtracted diastolic phase end point |
| 528 | ABP/PPG_D_RT_Are_wsDPE_SPP_wrtDPP_ABP/PPG_D_Are_wsDPE_SPP_wrtDN | The ratio of the area under the ABP/PPG cardiac cycle systolic phase peak with respect to diastolic phase peak, with subtracted diastolic phase end point to the area under the ABP/PPG cardiac cycle systolic phase peak with respect to dicrotic notch, with subtracted diastolic phase end point |
| 529 | ABP/PPG_D_RT_Are_wsDPE_SPP_wrtDPE_ABP/PPG_D_Are_wsDPE_SPP_wrtDN | The ratio of the area under the ABP/PPG cardiac cycle systolic phase peak with respect to diastolic phase end point, with subtracted diastolic phase end point to the area under the ABP/PPG cardiac cycle systolic phase peak with respect to dicrotic notch, with subtracted diastolic phase end point |
| 530 | ABP/PPG_D_RT_Are_wsDPE_DN_wrtDPP_ABP/PPG_D_Are_wsDPE_SPP_wrtDN | The ratio of the area under the ABP/PPG cardiac cycle dicrotic notch with respect to diastolic phase end peak, with subtracted diastolic phase end point to the area under the ABP/PPG cardiac cycle systolic phase peak with respect to dicrotic notch, with subtracted diastolic phase end point |
| 531 | ABP/PPG_D_RT_Are_wsDPE_DN_wrtDPE_ABP/PPG_D_Are_wsDPE_SPP_wrtDN | The ratio of the area under the ABP/PPG cardiac cycle dicrotic notch with respect to diastolic phase end point, with subtracted diastolic phase end point to the area under the ABP/PPG cardiac cycle systolic phase peak with respect to dicrotic notch, with subtracted diastolic phase end point |
| 532 | ABP/PPG_D_RT_Are_wsDPE_DPP_wrtDPE_ABP/PPG_D_Are_wsDPE_SPP_wrtDN | The ratio of the area under the ABP/PPG cardiac cycle diastolic phase peak with respect to diastolic phase end point, with subtracted diastolic phase end point to the area under the ABP/PPG cardiac cycle systolic phase peak with respect to dicrotic notch, with subtracted diastolic phase end point |
| 533 | ABP/PPG_D_RT_Are_wsDPE_SPP_wrtDPE_ABP/PPG_D_Are_wsDPE_SPP_wrtDPP | The ratio of the area under the ABP/PPG cardiac cycle systolic phase peak with respect to diastolic phase end point, with subtracted diastolic phase end point to the area under the ABP/PPG cardiac cycle systolic phase peak with respect to diastolic phase peak, with subtracted diastolic phase end point |
| 534 | ABP/PPG_D_RT_Are_wsDPE_DN_wrtDPP_ABP/PPG_D_Are_wsDPE_SPP_wrtDPP | The ratio of the area under the ABP/PPG cardiac cycle dicrotic notch with respect to diastolic phase peak, with subtracted diastolic phase end point to the area under the ABP/PPG cardiac cycle systolic phase peak with respect to diastolic phase peak, with subtracted diastolic phase end point |
| 535 | ABP/PPG_D_RT_Are_wsDPE_DN_wrtDPE_ABP/PPG_D_Are_wsDPE_SPP_wrtDPP | The ratio of the area under the ABP/PPG cardiac cycle dicrotic notch with respect to diastolic phase end point, with subtracted diastolic phase end point to the area under the ABP/PPG cardiac cycle systolic phase peak with respect to diastolic phase peak, with subtracted diastolic phase end point |
| 536 | ABP/PPG_D_RT_Are_wsDPE_DPP_wrtDPE_ABP/PPG_D_Are_wsDPE_SPP_wrtDPP | The ratio of the area under the ABP/PPG cardiac cycle diastolic phase peak with respect to diastolic phase end point, with subtracted diastolic phase end point to the area under the ABP/PPG cardiac cycle systolic phase peak with respect to diastolic phase peak, with subtracted diastolic phase end point |
| 537 | ABP/PPG_D_RT_Are_wsDPE_DN_wrtDPP_ABP/PPG_D_Are_wsDPE_SPP_wrtDPE | The ratio of the area under the ABP/PPG cardiac cycle dicrotic notch with respect to diastolic phase peak, with subtracted diastolic phase end point to the area under the ABP/PPG cardiac cycle systolic phase peak with respect to diastolic phase end point, with subtracted diastolic phase end point |
| 538 | ABP/PPG_D_RT_Are_wsDPE_DN_wrtDPE_ABP/PPG_D_Are_wsDPE_SPP_wrtDPE | The ratio of the area under the ABP/PPG cardiac cycle dicrotic notch with respect to diastolic phase end point, with subtracted diastolic phase end point to the area under the ABP/PPG cardiac cycle systolic phase peak with respect to diastolic phase end point, with subtracted diastolic phase end point |
| 539 | ABP/PPG_D_RT_Are_wsDPE_DPP_wrtDPE_ABP/PPG_D_Are_wsDPE_SPP_wrtDPE | The ratio of the area under the ABP/PPG cardiac cycle diastolic phase peak with respect to diastolic phase end point, with subtracted diastolic phase end point to the area under the ABP/PPG cardiac cycle systolic phase peak with respect to diastolic phase end point, with subtracted diastolic phase end point |
| 540 | ABP/PPG_D_RT_Are_wsDPE_DN_wrtDPE_ABP/PPG_D_Are_wsDPE_DN_wrtDPP | The ratio of the area under the ABP/PPG cardiac cycle dicrotic notch with respect to diastolic phase end point, with subtracted diastolic phase end point to the area under the ABP/PPG cardiac cycle dicrotic notch with respect to diastolic phase peak, with subtracted diastolic phase end point |
| 541 | ABP/PPG_D_RT_Are_wsDPE_DPP_wrtDPE_ABP/PPG_D_Are_wsDPE_DN_wrtDPP | The ratio of the area under the ABP/PPG cardiac cycle diastolic phase peak with respect to diastolic phase end point, with subtracted diastolic phase end point to the area under the ABP/PPG cardiac cycle dicrotic notch with respect to diastolic phase peak, with subtracted diastolic phase end point |
| 542 | ABP/PPG_D_RT_Are_wsDPE_DPP_wrtDPE_ABP/PPG_D_Are_wsDPE_DN_wrtDPE | The ratio of the area under the ABP/PPG cardiac cycle diastolic phase peak with respect to diastolic phase end point, with subtracted diastolic phase end point to the area under the ABP/PPG cardiac cycle dicrotic notch with respect to diastolic phase end point, with subtracted diastolic phase end point |
| 543 | ABP/PPG_D_RT_NAre_SPO_wrtDN_ABP/PPG_D_NAre_SPO_wrtSPP | The ratio of the area under the normalized ABP/PPG cardiac cycle systolic phase onset with respect to dicrotic notch to the area under the normalized ABP/PPG cardiac cycle systolic phase onset with respect to systolic phase peak |
| 544 | ABP/PPG_D_RT_NAre_SPO_wrtDPP_ABP/PPG_D_NAre_SPO_wrtSPP | The ratio of the area under the normalized ABP/PPG cardiac cycle systolic phase onset with respect to diastolic phase peak to the area under the normalized ABP/PPG cardiac cycle systolic phase onset with respect to systolic phase peak |
| 545 | ABP/PPG_D_RT_NAre_SPO_wrtDPE_ABP/PPG_D_NAre_SPO_wrtSPP | The ratio of the area under the normalized ABP/PPG cardiac cycle systolic phase onset with respect to diastolic phase end point to the area under the normalized ABP/PPG cardiac cycle systolic phase onset with respect to systolic phase peak |
| 546 | ABP/PPG_D_RT_NAre_SPP_wrtDN_ABP/PPG_D_NAre_SPO_wrtSPP | The ratio of the area under the normalized ABP/PPG cardiac cycle systolic phase peak with respect to dicrotic notch to the area under the normalized ABP/PPG cardiac cycle systolic phase onset with respect to systolic phase peak |
| 547 | ABP/PPG_D_RT_NAre_SPP_wrtDPP_ABP/PPG_D_NAre_SPO_wrtSPP | The ratio of the area under the normalized ABP/PPG cardiac cycle systolic phase peak with respect to diastolic phase peak to the area under the normalized ABP/PPG cardiac cycle systolic phase onset with respect to systolic phase peak |
| 548 | ABP/PPG_D_RT_NAre_SPP_wrtDPE_ABP/PPG_D_NAre_SPO_wrtSPP | The ratio of the area under the normalized ABP/PPG cardiac cycle systolic phase peak with respect to diastolic phase end point to the area under the normalized ABP/PPG cardiac cycle systolic phase onset with respect to systolic phase peak |
| 549 | ABP/PPG_D_RT_NAre_DN_wrtDPP_ABP/PPG_D_NAre_SPO_wrtSPP | The ratio of the area under the normalized ABP/PPG cardiac cycle dicrotic notch with respect to diastolic phase peak to the area under the normalized ABP/PPG cardiac cycle systolic phase onset with respect to systolic phase peak |
| 550 | ABP/PPG_D_RT_NAre_DN_wrtDPE_ABP/PPG_D_NAre_SPO_wrtSPP | The ratio of the area under the normalized ABP/PPG cardiac cycle dicrotic notch with respect to diastolic phase end point to the area under the normalized ABP/PPG cardiac cycle systolic phase onset with respect to systolic phase peak |
| 551 | ABP/PPG_D_RT_NAre_DPP_wrtDPE_ABP/PPG_D_NAre_SPO_wrtSPP | The ratio of the area under the normalized ABP/PPG cardiac cycle diastolic phase peak with respect to diastolic phase end point to the area under the normalized ABP/PPG cardiac cycle systolic phase onset with respect to systolic phase peak |
| 552 | ABP/PPG_D_RT_NAre_SPO_wrtDPP_ABP/PPG_D_NAre_SPO_wrtDN | The ratio of the area under the normalized ABP/PPG cardiac cycle systolic phase onset with respect to diastolic phase peak to the area under the normalized ABP/PPG cardiac cycle systolic phase onset with respect to dicrotic notch |
| 553 | ABP/PPG_D_RT_NAre_SPO_wrtDPE_ABP/PPG_D_NAre_SPO_wrtDN | The ratio of the area under the normalized ABP/PPG cardiac cycle systolic phase onset with respect to diastolic phase end point to the area under the normalized ABP/PPG cardiac cycle systolic phase onset with respect to dicrotic notch |
| 554 | ABP/PPG_D_RT_NAre_SPP_wrtDN_ABP/PPG_D_NAre_SPO_wrtDN | The ratio of the area under the normalized ABP/PPG cardiac cycle systolic phase peak with respect to dicrotic notch to the area under the normalized ABP/PPG cardiac cycle systolic phase onset with respect to dicrotic notch |
| 555 | ABP/PPG_D_RT_NAre_SPP_wrtDPP_ABP/PPG_D_NAre_SPO_wrtDN | The ratio of the area under the normalized ABP/PPG cardiac cycle systolic phase peak with respect to diastolic phase peak to the area under the normalized ABP/PPG cardiac cycle systolic phase onset with respect to dicrotic notch |
| 556 | ABP/PPG_D_RT_NAre_SPP_wrtDPE_ABP/PPG_D_NAre_SPO_wrtDN | The ratio of the area under the normalized ABP/PPG cardiac cycle systolic phase peak with respect to diastolic phase end point to the area under the normalized ABP/PPG cardiac cycle systolic phase onset with respect to dicrotic notch |
| 557 | ABP/PPG_D_RT_NAre_DN_wrtDPP_ABP/PPG_D_NAre_SPO_wrtDN | The ratio of the area under the normalized ABP/PPG cardiac cycle dicrotic notch with respect to diastolic phase peak to the area under the normalized ABP/PPG cardiac cycle systolic phase onset with respect to dicrotic notch |
| 558 | ABP/PPG_D_RT_NAre_DN_wrtDPE_ABP/PPG_D_NAre_SPO_wrtDN | The ratio of the area under the normalized ABP/PPG cardiac cycle dicrotic notch with respect to diastolic phase end point to the area under the normalized ABP/PPG cardiac cycle systolic phase onset with respect to dicrotic notch |
| 559 | ABP/PPG_D_RT_NAre_DPP_wrtDPE_ABP/PPG_D_NAre_SPO_wrtDN | The ratio of the area under the normalized ABP/PPG cardiac cycle diastolic phae peak with respect to diastolic phase end point to the area under the normalized ABP/PPG cardiac cycle systolic phase onset with respect to dicrotic notch |
| 560 | ABP/PPG_D_RT_NAre_SPO_wrtDPE_ABP/PPG_D_NAre_SPO_wrtDPP | The ratio of the area under the normalized ABP/PPG cardiac cycle systolic phase onset with respect to diastolic phase end point to the area under the normalized ABP/PPG cardiac cycle systolic phase onset with respect to diastolic phase peak |
| 561 | ABP/PPG_D_RT_NAre_SPP_wrtDN_ABP/PPG_D_NAre_SPO_wrtDPP | The ratio of the area under the normalized ABP/PPG cardiac cycle systolic phase peak with respect to dicrotic notch to the area under the normalized ABP/PPG cardiac cycle systolic phase onset with respect to diastolic phase peak |
| 562 | ABP/PPG_D_RT_NAre_SPP_wrtDPP_ABP/PPG_D_NAre_SPO_wrtDPP | The ratio of the area under the normalized ABP/PPG cardiac cycle systolic phase peak with respect to diastolic phase peak to the area under the normalized ABP/PPG cardiac cycle systolic phase onset with respect to diastolic phase peak |
| 563 | ABP/PPG_D_RT_NAre_SPP_wrtDPE_ABP/PPG_D_NAre_SPO_wrtDPP | The ratio of the area under the normalized ABP/PPG cardiac cycle systolic phase peak with respect to diastolic phase end point to the area under the normalized ABP/PPG cardiac cycle systolic phase onset with respect to diastolic phase peak |
| 564 | ABP/PPG_D_RT_NAre_DN_wrtDPP_ABP/PPG_D_NAre_SPO_wrtDPP | The ratio of the area under the normalized ABP/PPG cardiac cycle dicrotic notch with respect to diastolic phase peak to the area under the normalized ABP/PPG cardiac cycle systolic phase onset with respect to diastolic phase peak |
| 565 | ABP/PPG_D_RT_NAre_DN_wrtDPE_ABP/PPG_D_NAre_SPO_wrtDPP | The ratio of the area under the normalized ABP/PPG cardiac cycle dicrotic notch with respect to diastolic phase end point to the area under the normalized ABP/PPG cardiac cycle systolic phase onset with respect to diastolic phase peak |
| 566 | ABP/PPG_D_RT_NAre_DPP_wrtDPE_ABP/PPG_D_NAre_SPO_wrtDPP | The ratio of the area under the normalized ABP/PPG cardiac cycle diastolic phase peak with respect to diastolic phase end point to the area under the normalized ABP/PPG cardiac cycle systolic phase onset with respect to diastolic phase peak |
| 567 | ABP/PPG_D_RT_NAre_SPP_wrtDN_ABP/PPG_D_NAre_SPO_wrtDPE | The ratio of the area under the normalized ABP/PPG cardiac cycle systolic phase peak with respect to dicrotic notch to the area under the normalized ABP/PPG cardiac cycle systolic phase onset with respect to diastolic phase end point |
| 568 | ABP/PPG_D_RT_NAre_SPP_wrtDPP_ABP/PPG_D_NAre_SPO_wrtDPE | The ratio of the area under the normalized ABP/PPG cardiac cycle systolic phase peak with respect to diastolic phase peak to the area under the normalized ABP/PPG cardiac cycle systolic phase onset with respect to diastolic phase end point |
| 569 | ABP/PPG_D_RT_NAre_SPP_wrtDPE_ABP/PPG_D_NAre_SPO_wrtDPE | The ratio of the area under the normalized ABP/PPG cardiac cycle systolic phase peak with respect to diastolic phase end point to the area under the normalized ABP/PPG cardiac cycle systolic phase onset with respect to diastolic phase end point |
| 570 | ABP/PPG_D_RT_NAre_DN_wrtDPP_ABP/PPG_D_NAre_SPO_wrtDPE | The ratio of the area under the normalized ABP/PPG cardiac cycle dicrotic notch peak with respect to diastolic phase peak to the area under the normalized ABP/PPG cardiac cycle systolic phase onset with respect to diastolic phase end point |
| 571 | ABP/PPG_D_RT_NAre_DN_wrtDPE_ABP/PPG_D_NAre_SPO_wrtDPE | The ratio of the area under the normalized ABP/PPG cardiac cycle dicrotic notch with respect to diastolic phase end point to the area under the normalized ABP/PPG cardiac cycle systolic phase onset with respect to diastolic phase end point |
| 572 | ABP/PPG_D_RT_NAre_DPP_wrtDPE_ABP/PPG_D_NAre_SPO_wrtDPE | The ratio of the area under the normalized ABP/PPG cardiac cycle diastolic phase peak with respect to diastolic phase end point to the area under the normalized ABP/PPG cardiac cycle systolic phase onset with respect to diastolic phase end point |
| 573 | ABP/PPG_D_RT_NAre_SPP_wrtDPP_ABP/PPG_D_NAre_SPP_wrtDN | The ratio of the area under the normalized ABP/PPG cardiac cycle systolic phase peak with respect to diastolic phase peak to the area under the normalized ABP/PPG cardiac cycle systolic phase peak with respect to dicrotic notch |
| 574 | ABP/PPG_D_RT_NAre_SPP_wrtDPE_ABP/PPG_D_NAre_SPP_wrtDN | The ratio of the area under the normalized ABP/PPG cardiac cycle systolic phase peak with respect to diastolic phase end point to the area under the normalized ABP/PPG cardiac cycle systolic phase peak with respect to dicrotic notch |
| 575 | ABP/PPG_D_RT_NAre_DN_wrtDPP_ABP/PPG_D_NAre_SPP_wrtDN | The ratio of the area under the normalized ABP/PPG cardiac cycle dicrotic notch with respect to diastolic phase peak to the area under the normalized ABP/PPG cardiac cycle systolic phase peak with respect to dicrotic notch |
| 576 | ABP/PPG_D_RT_NAre_DN_wrtDPE_ABP/PPG_D_NAre_SPP_wrtDN | The ratio of the area under the normalized ABP/PPG cardiac cycle dicrotic notch with respect to diastolic phase end point to the area under the normalized ABP/PPG cardiac cycle systolic phase peak with respect to dicrotic notch |
| 577 | ABP/PPG_D_RT_NAre_DPP_wrtDPE_ABP/PPG_D_NAre_SPP_wrtDN | The ratio of the area under the normalized ABP/PPG cardiac cycle diastolic phase peak with respect to diastolic phase end point to the area under the normalized ABP/PPG cardiac cycle systolic phase peak with respect to dicrotic notch |
| 578 | ABP/PPG_D_RT_NAre_SPP_wrtDPE_ABP/PPG_D_NAre_SPP_wrtDPP | The ratio of the area under the normalized ABP/PPG cardiac cycle systolic phase peak with respect to diastolic phase end point to the area under the normalized ABP/PPG cardiac cycle systolic phase peak with respect to diastolic phase peak |
| 579 | ABP/PPG_D_RT_NAre_DN_wrtDPP_ABP/PPG_D_NAre_SPP_wrtDPP | The ratio of the area under the normalized ABP/PPG cardiac cycle dicrotic notch with respect to diastolic phase peak to the area under the normalized ABP/PPG cardiac cycle systolic phase peak with respect to diastolic phase peak |
| 580 | ABP/PPG_D_RT_NAre_DN_wrtDPE_ABP/PPG_D_NAre_SPP_wrtDPP | The ratio of the area under the normalized ABP/PPG cardiac cycle dicrotic notch with respect to diastolic phase end point to the area under the normalized ABP/PPG cardiac cycle systolic phase peak with respect to diastolic phase peak |
| 581 | ABP/PPG_D_RT_NAre_DPP_wrtDPE_ABP/PPG_D_NAre_SPP_wrtDPP | The ratio of the area under the normalized ABP/PPG cardiac cycle diastolic phase peak with respect to diastolic phase end point to the area under the normalized ABP/PPG cardiac cycle systolic phase peak with respect to diastolic phase peak |
| 582 | ABP/PPG_D_RT_NAre_DN_wrtDPP_ABP/PPG_D_NAre_SPP_wrtDPE | The ratio of the area under the normalized ABP/PPG cardiac cycle dicrotic notch with respect to diastolic phase peak to the area under the normalized ABP/PPG cardiac cycle systolic phase peak with respect to diastolic phase end point |
| 583 | ABP/PPG_D_RT_NAre_DN_wrtDPE_ABP/PPG_D_NAre_SPP_wrtDPE | The ratio of the area under the normalized ABP/PPG cardiac cycle dicrotic notch with respect to diastolic phase end point to the area under the normalized ABP/PPG cardiac cycle systolic phase peak with respect to diastolic phase end point |
| 584 | ABP/PPG_D_RT_NAre_DPP_wrtDPE_ABP/PPG_D_NAre_SPP_wrtDPE | The ratio of the area under the normalized ABP/PPG cardiac cycle diastolic phase peak with respect to diastolic phase end point to the area under the normalized ABP/PPG cardiac cycle systolic phase peak with respect to diastolic phase end point |
| 585 | ABP/PPG_D_RT_NAre_DN_wrtDPE_ABP/PPG_D_NAre_DN_wrtDPP | The ratio of the area under the normalized ABP/PPG cardiac cycle dicrotic notch with respect to diastolic phase end point to the area under the normalized ABP/PPG cardiac cycle dicrotic notch with respect to diastolic phase peak |
| 586 | ABP/PPG_D_RT_NAre_DPP_wrtDPE_ABP/PPG_D_NAre_DN_wrtDPP | The ratio of the area under the normalized ABP/PPG cardiac cycle diastolic phase peak with respect to diastolic phase end point to the area under the normalized ABP/PPG cardiac cycle dicrotic notch with respect to diastolic phase peak |
| 587 | ABP/PPG_D_RT_NAre_DPP_wrtDPE_ABP/PPG_D_NAre_DN_wrtDPE | The ratio of the area under the normalized ABP/PPG cardiac cycle diastolic phase peak with respect to diastolic phase end point to the area under the normalized ABP/PPG cardiac cycle dicrotic notch with respect to diastolic phase end point |
| 588 | ABP/PPG_D_RT_NAre_wsDPE_SPO_wrtDN_ABP/PPG_D_NAre_wsDPE_SPO_wrtSPP | The ratio of the area under the normalized ABP/PPG cardiac cycle systolic phase onset with respect to dicrotic notch duration, with subtracted diastolic phase end point to the area under the normalized ABP/PPG cardiac cycle systolic phase onset with respect to systolic phase peak, with subtracted diastolic phase end point |
| 589 | ABP/PPG_D_RT_NAre_wsDPE_SPO_wrtDPP_ABP/PPG_D_NAre_wsDPE_SPO_wrtSPP | The ratio of the area under the normalized ABP/PPG cardiac cycle systolic phase onset with respect to diastolic phase peak, with subtracted diastolic phase end point to the area under the normalized ABP/PPG cardiac cycle systolic phase onset with respect to systolic phase peak, with subtracted diastolic phase end point |
| 590 | ABP/PPG_D_RT_NAre_WsDPE_SPO_WrtDPE_ABP/PPG_D_NAre_wsDPE_SPO_wrtSPP | The ratio of the area under the normalized ABP/PPG cardiac cycle systolic phase onset with respect to diastolic phase end point, with subtracted diastolic phase end point to the area under the normalized ABP/PPG cardiac cycle systolic phase onset with respect to systolic phase peak, with subtracted diastolic phase end point |
| 591 | ABP/PPG_D_RT_NAre_wsDPE_SPP_wrtDN_ABP/PPG_D_NAre_wsDPE_SPO_wrtSPP | The ratio of the area under the normalized ABP/PPG cardiac cycle systolic phase peak with respect to dicrotic notch, with subtracted diastolic phase end point to the area under the normalized ABP/PPG cardiac cycle systolic phase onset with respect to systolic phase peak, with subtracted diastolic phase end point |
| 592 | ABP/PPG_D_RT_NAre_wsDPE_SPP_wrtDPP_ABP/PPG_D_NAre_wsDPE_SPO_wrtSPP | The ratio of the area under the normalized ABP/PPG cardiac cycle systolic phase peak with respect to diastolic phase peak, with subtracted diastolic phase end point to the area under the normalized ABP/PPG cardiac cycle systolic phase onset with respect to systolic phase peak, with subtracted diastolic phase end point |
| 593 | ABP/PPG_D_RT_NAre_wsDPE_SPP_wrtDPE_ABP/PPG_D_NAre_wsDPE_SPO_wrtSPP | The ratio of the area under the normalized ABP/PPG cardiac cycle systolic phase peak with respect to diastolic phase end point, with subtracted diastolic phase end point to the area under the normalized ABP/PPG cardiac cycle systolic phase onset with respect to systolic phase peak, with subtracted diastolic phase end point |
| 594 | ABP/PPG_D_RT_NAre_wsDPE_DN_wrtDPP_ABP/PPG_D_NAre_wsDPE_SPO_wrtSPP | The ratio of the area under the normalized ABP/PPG cardiac cycle dicrotic notch with respect to diastolic phase peak, with subtracted diastolic phase end point to the area under the normalized ABP/PPG cardiac cycle systolic phase onset with respect to systolic phase peak, with subtracted diastolic phase end point |
| 595 | ABP/PPG_D_RT_NAre_wsDPE_DN_wrtDPE_ABP/PPG_D_NAre_wsDPE_SPO_wrtSPP | The ratio of the area under the normalized ABP/PPG cardiac cycle dicrotic notch with respect to diastolic phase end point, with subtracted diastolic phase end point to the area under the normalized ABP/PPG cardiac cycle systolic phase onset with respect to systolic phase peak, with subtracted diastolic phase end point |
| 596 | ABP/PPG_D_RT_NAre_wsDPE_DPP_wrtDPE_ABP/PPG_D_NAre_wsDPE_SPO_wrtSPP | The ratio of the area under the normalized ABP/PPG cardiac cycle diastolic phase peak with respect to diastolic phase end point, with subtracted diastolic phase end point to the area under the normalized ABP/PPG cardiac cycle systolic phase onset with respect to systolic phase peak, with subtracted diastolic phase end point |
| 597 | ABP/PPG_D_RT_NAre_wsDPE_SPO_wrtDPP_ABP/PPG_D_NAre_wsDPE_SPO_wrtDN | The ratio of the area under the normalized ABP/PPG cardiac cycle systolic phase onset with respect to diastolic phase peak, with subtracted diastolic phase end point to the area under the normalized ABP/PPG cardiac cycle systolic phase onset with respect to dicrotic notch, with subtracted diastolic phase end point |
| 598 | ABP/PPG_D_RT_NAre_wsDPE_SPO_wrtDPE_ABP/PPG_D_NAre_wsDPE_SPO_wrtDN | The ratio of the area under the normalized ABP/PPG cardiac cycle systolic phase onset with respect to diastolic phase end point, with subtracted diastolic phase end point to the area under the normalized ABP/PPG cardiac cycle systolic phase onset with respect to dicrotic notch, with subtracted diastolic phase end point |
| 599 | ABP/PPG_D_RT_NAre_wsDPE_SPP_wrtDN_ABP/PPG_D_NAre_wsDPE_SPO_wrtDN | The ratio of the area under the normalized ABP/PPG cardiac cycle systolic phase peak with respect to dicrotic notch, with subtracted diastolic phase end point to the area under the normalized ABP/PPG cardiac cycle systolic phase onset with respect to dicrotic notch, with subtracted diastolic phase end point |
| 600 | ABP/PPG_D_RT_NAre_wsDPE_SPP_wrtDPP_ABP/PPG_D_NAre_wsDPE_SPO_wrtDN | The ratio of the area under the normalized ABP/PPG cardiac cycle systolic phase peak with respect to diastolic phase peak, with subtracted diastolic phase end point to the area under the normalized ABP/PPG cardiac cycle systolic phase onset with respect to dicrotic notch, with subtracted diastolic phase end point |
| 601 | ABP/PPG_D_RT_NAre_wsDPE_SPP_wrtDPE_ABP/PPG_D_NAre_wsDPE_SPO_wrtDN | The ratio of the area under the normalized ABP/PPG cardiac cycle systolic phase peak with respect to diastolic phase end point, with subtracted diastolic phase end point to the area under the normalized ABP/PPG cardiac cycle systolic phase onset with respect to dicrotic notch, with subtracted diastolic phase end point |
| 602 | ABP/PPG_D_RT_NAre_wsDPE_DN_wrtDPP_ABP/PPG_D_NAre_wsDPE_SPO_wrtDN | The ratio of the area under the normalized ABP/PPG cardiac cycle dicrotic notch with respect to diastolic phase peak, with subtracted diastolic phase end point to the area under the normalized ABP/PPG cardiac cycle systolic phase onset with respect to dicrotic notch, with subtracted diastolic phase end point |
| 603 | ABP/PPG_D_RT_NAre_wsDPE_DN_wrtDPE_ABP/PPG_D_NAre_wsDPE_SPO_wrtDN | The ratio of the area under the normalized ABP/PPG cardiac cycle dicrotic notch with respect to diastolic phase end point, with subtracted diastolic phase end point to the area under the normalized ABP/PPG cardiac cycle systolic phase onset with respect to dicrotic notch, with subtracted diastolic phase end point |
| 604 | ABP/PPG_D_RT_NAre_wsDPE_DPP_wrtDPE_ABP/PPG_D_NAre_wsDPE_SPO_wrtDN | The ratio of the area under the normalized ABP/PPG cardiac cycle diastolic phase peak with respect to diastolic phase end point, with subtracted diastolic phase end point to the area under the normalized ABP/PPG cardiac cycle systolic phase onset with respect to dicrotic notch, with subtracted diastolic phase end point |
| 605 | ABP/PPG_D_RT_NAre_wsDPE_SPO_wrtDPE_ABP/PPG_D_NAre_wsDPE_SPO_wrtDPP | The ratio of normalized area of systolic phase onset with respect to diastolic phase end point, with subtracted diastolic phase end point to the area under the normalized ABP/PPG cardiac cycle systolic phase onset with respect to diastolic phase peak, with subtracted diastolic phase end point |
| 606 | ABP/PPG_D_RT_NAre_wsDPE_SPP_wrtDN_ABP/PPG_D_NAre_wsDPE_SPO_wrtDPP | The ratio of normalized area of systolic phase peak with respect to dicrotic notch, with subtracted diastolic phase end point to the area under the normalized ABP/PPG cardiac cycle systolic phase onset with respect to diastolic phase peak, with subtracted diastolic phase end point |
| 607 | ABP/PPG_D_RT_NAre_wsDPE_SPP_wrtDPP_ABP/PPG_D_NAre_wsDPE_SPO_wrtDPP | The ratio of the area under the normalized ABP/PPG cardiac cycle systolic phase peak with respect to diastolic phase peak, with subtracted diastolic phase end point to the area under the normalized ABP/PPG cardiac cycle systolic phase onset with respect to diastolic phase peak, with subtracted diastolic phase end point |
| 608 | ABP/PPG_D_RT_NAre_wsDPE_SPP_wrtDPE_ABP/PPG_D_NAre_wsDPE_SPO_wrtDPP | The ratio of normalized area of systolic phase peak with respect to diastolic phase end point, with subtracted diastolic phase end point to the area under the normalized ABP/PPG cardiac cycle systolic phase onset with respect to diastolic phase peak, with subtracted diastolic phase end point |
| 609 | ABP/PPG_D_RT_NAre_wsDPE_DN_wrtDPP_ABP/PPG_D_NAre_wsDPE_SPO_wrtDPP | The ratio of the area under the normalized ABP/PPG cardiac cycle dicrotic notch with respect to diastolic phase peak, with subtracted diastolic phase end point to the area under the normalized ABP/PPG cardiac cycle systolic phase onset with respect to diastolic phase peak, with subtracted diastolic phase end point |
| 610 | ABP/PPG_D_RT_NAre_wsDPE_DN_wrtDPE_ABP/PPG_D_NAre_wsDPE_SPO_wrtDPP | The ratio of normalized area of dicrotic notch with respect to diastolic phase end point, with subtracted diastolic phase end point to the area under the normalized ABP/PPG cardiac cycle systolic phase onset with respect to diastolic phase peak, with subtracted diastolic phase end point |
| 611 | ABP/PPG_D_RT_NAre_wsDPE_DPP_wrtDPE_ABP/PPG_D_NAre_wsDPE_SPO_wrtDPP | The ratio of normalized area of diastolic phase peak with respect to diastolic phase end point, with subtracted diastolic phase end point to the area under the normalized ABP/PPG cardiac cycle systolic phase onset with respect to diastolic phase peak, with subtracted diastolic phase end point |
| 612 | ABP/PPG_D_RT_NAre_wsDPE_SPP_wrtDN_ABP/PPG_D_NAre_wsDPE_SPO_wrtDPE | The ratio of normalized area of systolic phase peak with respect to dicrotic notch, with subtracted diastolic phase end point to the area under the normalized ABP/PPG cardiac cycle systolic phase onset with respect to diastolic phase end point, with subtracted diastolic phase end point |
| 613 | ABP/PPG_D_RT_NAre_wsDPE_SPP_wrtDPP_ABP/PPG_D_NAre_wsDPE_SPO_wrtDPE | The ratio of the area under the normalized ABP/PPG cardiac cycle systolic phase peak with respect to diastolic phase peak, with subtracted diastolic phase end point to the area under the normalized ABP/PPG cardiac cycle systolic phase onset with respect to diastolic phase end point, with subtracted diastolic phase end point |
| 614 | ABP/PPG_D_RT_NAre_wsDPE_SPP_wrtDPE_ABP/PPG_D_NAre_wsDPE_SPO_wrtDPE | The ratio of the area under the normalized ABP/PPG cardiac cycle systolic phase peak with respect to diastolic phase end point, with subtracted diastolic phase end point to the area under the normalized ABP/PPG cardiac cycle systolic phase onset with respect to diastolic phase end point, with subtracted diastolic phase end point |
| 615 | ABP/PPG_D_RT_NAre_wsDPE_DN_wrtDPP_ABP/PPG_D_NAre_wsDPE_SPO_wrtDPE | The ratio of the area under the normalized ABP/PPG cardiac cycle dicrotic notch with respect to diastolic phase peak, with subtracted diastolic phase end point to the area under the normalized ABP/PPG cardiac cycle systolic phase onset with respect to diastolic phase end point |
| 616 | ABP/PPG_D_RT_NAre_wsDPE_DN_wrtDPE_ABP/PPG_D_NAre_wsDPE_SPO_wrtDPE | The ratio of the area under the normalized ABP/PPG cardiac cycle dicrotic notch with respect to diastolic phase end point, with subtracted diastolic phase end point to the area under the normalized ABP/PPG cardiac cycle systolic phase onset with respect to diastolic phase end point, with subtracted diastolic phase end point |
| 617 | ABP/PPG_D_RT_NAre_wsDPE_DPP_wrtDPE_ABP/PPG_D_NAre_wsDPE_SPO_wrtDPE | The ratio of the area under the normalized ABP/PPG cardiac cycle diastolic phase peak with respect to diastolic phase end point, with subtracted diastolic phase end point to the area under the normalized ABP/PPG cardiac cycle systolic phase onset with respect to diastolic phase end point, with subtracted diastolic phase end point |
| 618 | ABP/PPG_D_RT_NAre_wsDPE_SPP_wrtDPP_ABP/PPG_D_NAre_wsDPE_SPP_wrtDN | The ratio of the area under the normalized ABP/PPG cardiac cycle systolic phase peak with respect to diastolic phase peak, with subtracted diastolic phase end point to the area under the normalized ABP/PPG cardiac cycle systolic phase peak with respect to dicrotic notch, with subtracted diastolic phase end point |
| 619 | ABP/PPG_D_RT_NAre_wsDPE_SPP_wrtDPE_ABP/PPG_D_NAre_wsDPE_SPP_wrtDN | The ratio of the area under the normalized ABP/PPG cardiac cycle systolic phase peak with respect to diastolic phase end point, with subtracted diastolic phase end point to the area under the normalized ABP/PPG cardiac cycle systolic phase peak with respect to dicrotic notch, with subtracted diastolic phase end point |
| 620 | ABP/PPG_D_RT_NAre_wsDPE_DN_wrtDPP_ABP/PPG_D_NAre_wsDPE_SPP_wrtDN | The ratio of the area under the normalized ABP/PPG cardiac cycle dicrotic notch with respect to diastolic phase peak, with subtracted diastolic phase end point to the area under the normalized ABP/PPG cardiac cycle systolic phase peak with respect to dicrotic notch, with subtracted diastolic phase end point |
| 621 | ABP/PPG_D_RT_NAre_wsDPE_DN_wrtDPE_ABP/PPG_D_NAre_wsDPE_SPP_wrtDN | The ratio of the area under the normalized ABP/PPG cardiac cycle dicrotic notch with respect to diastolic phase end point, with subtracted diastolic phase end point to the area under the normalized ABP/PPG cardiac cycle systolic phase peak with respect to dicrotic notch, with subtracted diastolic phase end point |
| 622 | ABP/PPG_D_RT_NAre_wsDPE_DPP_wrtDPE_ABP/PPG_D_NAre_wsDPE_SPP_wrtDN | The ratio of the area under the normalized ABP/PPG cardiac cycle diastolic phase peak with respect to diastolic phase end point, with subtracted diastolic phase end point to the area under the normalized ABP/PPG cardiac cycle systolic phase peak with respect to dicrotic notch, with subtracted diastolic phase end point |
| 623 | ABP/PPG_D_RT_NAre_wsDPE_SPP_wrtDPE_ABP/PPG_D_NAre_wsDPE_SPP_wrtDPP | The ratio of the area under the normalized ABP/PPG cardiac cycle systolic phase peak with respect to diastolic phase end point, with subtracted diastolic phase end point to the area under the normalized ABP/PPG cardiac cycle systolic phase peak with respect to diastolic phase peak, with subtracted diastolic phase end point |
| 624 | ABP/PPG_D_RT_NAre_wsDPE_DN_wrtDPP_ABP/PPG_D_NAre_wsDPE_SPP_wrtDPP | The ratio of the area under the normalized ABP/PPG cardiac cycle dicrotic notch with respect to diastolic phase peak, with subtracted diastolic phase end point to the area under the normalized ABP/PPG cardiac cycle systolic phase peak with respect to diastolic phase peak, with subtracted diastolic phase end point |
| 625 | ABP/PPG_D_RT_NAre_wsDPE_DN_wrtDPE_ABP/PPG_D_NAre_wsDPE_SPP_wrtDPP | The ratio of normalized area of dicrotic notch with respect to diastolic phase end point, with subtracted diastolic phase end point to the area under the normalized ABP/PPG cardiac cycle systolic phase peak with respect to diastolic phase peak, with subtracted diastolic phase end point |
| 626 | ABP/PPG_D_RT_NAre_wsDPE_DPP_wrtDPE_ABP/PPG_D_NAre_wsDPE_SPP_wrtDPP | The ratio of the area under the normalized ABP/PPG cardiac cycle diastolic phase peak with respect to diastolic phase end point, with subtracted diastolic phase end point to the area under the normalized ABP/PPG cardiac cycle systolic phase peak with respect to diastolic phase peak, with subtracted diastolic phase end point |
| 627 | ABP/PPG_D_RT_NAre_wsDPE_DN_wrtDPP_ABP/PPG_D_NAre_wsDPE_SPP_wrtDPE | The ratio of the area under the normalized ABP/PPG cardiac cycle dicrotic notch with respect to diastolic phase peak, with subtracted diastolic phase end point to the area under the normalized ABP/PPG cardiac cycle systolic phase peak with respect to diastolic phase end point, with subtracted diastolic phase end point |
| 628 | ABP/PPG_D_RT_NAre_wsDPE_DN_wrtDPE_ABP/PPG_D_NAre_wsDPE_SPP_wrtDPE | The ratio of the area under the normalized ABP/PPG cardiac cycle dicrotic notch with respect to diastolic phase end point, with subtracted diastolic phase end point to the area under the normalized ABP/PPG cardiac cycle systolic phase peak with respect to diastolic phase end point, with subtracted diastolic phase end point |
| 629 | ABP/PPG_D_RT_NAre_wsDPE_DPP_wrtDPE_ABP/PPG_D_NAre_wsDPE_SPP_wrtDPE | The ratio of the area under the normalized ABP/PPG cardiac cycle diastolic phase peak with respect to diastolic phase end point, with subtracted diastolic phase end point to the area under the normalized ABP/PPG cardiac cycle systolic phase peak with respect to diastolic phase end point, with subtracted diastolic phase end point |
| 630 | ABP/PPG_D_RT_NAre_wsDPE_DN_wrtDPE_ABP/PPG_D_NAre_wsDPE_DN_wrtDPP | The ratio of normalized area of dicrotic notch with respect to diastolic phase end point, with subtracted diastolic phase end point to the area under the normalized ABP/PPG cardiac cycle dicrotic notch with respect to diastolic phase peak, with subtracted diastolic phase end point |
| 631 | ABP/PPG_D_RT_NAre_wsDPE_DPP_wrtDPE_ABP/PPG_D_NAre_wsDPE_DN_wrtDPP | The ratio of normalized area of diastolic phase peak with respect to diastolic phase end point, with subtracted diastolic phase end point to the area under the normalized ABP/PPG cardiac cycle dicrotic notch with respect to diastolic phase peak, with subtracted diastolic phase end point |
| 632 | ABP/PPG_D_RT_NAre_wsDPE_DPP_wrtDPE_ABP/PPG_D_NAre_wsDPE_DN_wrtDPE | The ratio of the area under the normalized ABP/PPG cardiac cycle diastolic phase peak with respect to diastolic phase end point, with subtracted diastolic phase end point to the area under the normalized ABP/PPG cardiac cycle dicrotic notch with respect to diastolic phase end point, with subtracted diastolic phase end point |

18. **First derivative features (636-642)**

| 636 | ABP/PPG_FD_NO_ZC | ABP/PPG cardiac cycle first derivative number of zero crossings |
| --- | --- | --- |
| 637 | ABP/PPG_AM_FD_Upeak_wrtzero | ABP/PPG cardiac cycle first derivative U peak amplitude with respect to zero |
| 638 | ABP/PPG_AM_FD_Vpeak_wrtzero | ABP/PPG cardiac cycle first derivative Vpeak amplitude with respect to zero |
| 639 | ABP/PPG_AM_FD_Wpeak_wrtzero | ABP/PPG cardiac cycle first derivative Wpeak amplitude with respect to zero |
| 640 | ABP/PPG_AM_FD_Upeak_wrtVpeak | ABP/PPG cardiac cycle first derivative Upeak amplitude with respect to Vpeak |
| 641 | ABP/PPG_AM_FD_Upeak_wrtWpeak | ABP/PPG cardiac cycle first derivative Upeak amplitude with respect to Wpeak |
| 642 | ABP/PPG_AM_FD_Vpeak_wrtWpeak | ABP/PPG cardiac cycle first derivative Vpeak amplitude with respect to Wpeak |

19. **First derivative amplitude ratio features (643-657)**

| 643 | ABP/PPG_AM_RT_FD_Vpeak_wrtzero_ABP/PPG_AM_FD_Upeak_wrtzero | The ratio of the ABP/PPG cardiac cycle first derivative Vpeak amplitude with respect to zero to the ABP/PPG cardiac cycle first derivative Upeak amplitude with respect to zero |
| --- | --- | --- |
| 644 | ABP/PPG_AM_RT_FD_Wpeak_wrtzero_ABP/PPG_AM_FD_Upeak_wrtzero | The ratio of the ABP/PPG cardiac cycle first derivative Wpeak amplitude with respect to zero to the ABP/PPG cardiac cycle first derivative Upeak amplitude with respect to zero |
| 645 | ABP/PPG_AM_RT_FD_Upeak_wrtVpeak_ABP/PPG_AM_FD_Upeak_wrtzero | The ratio of the ABP/PPG cardiac cycle first derivative Upeak amplitude with respect to Vpeak to the ABP/PPG cardiac cycle first derivative Upeak amplitude with respect to zero |
| 646 | ABP/PPG_AM_RT_FD_Upeak_wrtWpeak_ABP/PPG_AM_FD_Upeak_wrtzero | The ratio of the ABP/PPG cardiac cycle first derivative Upeak amplitude with respect to Wpeak to the ABP/PPG cardiac cycle first derivative Upeak amplitude with respect to zero |
| 647 | ABP/PPG_AM_RT_FD_Vpeak_wrtWpeak_ABP/PPG_AM_FD_Upeak_wrtzero | The ratio of the ABP/PPG cardiac cycle first derivative Vpeak amplitude with respect to Wpeak to the ABP/PPG cardiac cycle first derivative Upeak amplitude with respect to zero |
| 648 | ABP/PPG_AM_RT_FD_Wpeak_wrtzero_ABP/PPG_AM_FD_Vpeak_wrtzero | The ratio of the ABP/PPG cardiac cycle first derivative Wpeak amplitude with respect to zero to the ABP/PPG cardiac cycle first derivative Vpeak amplitude with respect to zero |
| 649 | ABP/PPG_AM_RT_FD_Upeak_wrtVpeak_ABP/PPG_AM_FD_Vpeak_wrtzero | The ratio of the ABP/PPG cardiac cycle first derivative Upeak amplitude with respect to Vpeak to the ABP/PPG cardiac cycle first derivative Vpeak amplitude with respect to zero |
| 650 | ABP/PPG_AM_RT_FD_Upeak_wrtWpeak_ABP/PPG_AM_FD_Vpeak_wrtzero | The ratio of the ABP/PPG cardiac cycle first derivative Upeak amplitude with respect to Wpeak to the ABP/PPG cardiac cycle first derivative Vpeak amplitude with respect to zero |
| 651 | ABP/PPG_AM_RT_FD_Vpeak_wrtWpeak_ABP/PPG_AM_FD_Vpeak_wrtzero | The ratio of the ABP/PPG cardiac cycle first derivative Vpeak amplitude with respect to Wpeak to the ABP/PPG cardiac cycle first derivative Vpeak amplitude with respect to zero |
| 652 | ABP/PPG_AM_RT_FD_Upeak_wrtVpeak_ABP/PPG_AM_FD_Wpeak_wrtzero | The ratio of the ABP/PPG cardiac cycle first derivative Upeak amplitude with respect to Vpeak to the ABP/PPG cardiac cycle first derivative W peak amplitude with respect to zero |
| 653 | ABP/PPG_AM_RT_FD_Upeak_wrtWpeak_ABP/PPG_AM_FD_Wpeak_wrtzero | The ratio of the ABP/PPG cardiac cycle first derivative Upeak amplitude with respect to Wpeak to the ABP/PPG cardiac cycle first derivative Wpeak amplitude with respect to zero |
| 654 | ABP/PPG_AM_RT_FD_Vpeak_wrtWpeak_ABP/PPG_AM_FD_Wpeak_wrtzero | The ratio of the ABP/PPG cardiac cycle first derivative Vpeak amplitude with respect to Wpeak to the ABP/PPG cardiac cycle first derivative Wpeak amplitude with respect to zero |
| 655 | ABP/PPG_AM_RT_FD_Upeak_wrtWpeak_ABP/PPG_AM_FD_Upeak_wrtVpeak | The ratio of the ABP/PPG cardiac cycle first derivative Upeak amplitude with respect to Wpeak to the ABP/PPG cardiac cycle first derivative Upeak amplitude with respect to Vpeak |
| 656 | ABP/PPG_AM_RT_FD_Vpeak_wrtWpeak_ABP/PPG_AM_FD_Upeak_wrtVpeak | The ratio of the ABP/PPG cardiac cycle first derivative Vpeak amplitude with respect to Wpeak to the ABP/PPG cardiac cycle first derivative Upeak amplitude with respect to Vpeak |
| 657 | ABP/PPG_AM_RT_FD_Vpeak_wrtWpeak_ABP/PPG_AM_FD_Upeak_wrtWpeak | The ratio of the ABP/PPG cardiac cycle first derivative Vpeak amplitude with respect to Wpeak to the ABP/PPG cardiac cycle first derivative Upeak amplitude with respect to Wpeak |

20. **First derivative duration features (658-660)**

| 658 | ABP/PPG_D_FD_Upeak_wrtVpeak | Duration of the ABP/PPG cardiac cycle first derivative Upeak with respect to Vpeak |
| --- | --- | --- |
| 659 | ABP/PPG_D_FD_Upeak_wrtWpeak | Duration of the ABP/PPG cardiac cycle first derivative Upeak with respect to Wpeak |
| 660 | ABP/PPG_D_FD_Vpeak_wrtWpeak | Duration of the ABP/PPG cardiac cycle first derivative Vpeak with respect to Wpeak |

21. **First derivative duration ratio features (661-663)**

| 661 | ABP/PPG_D_RT_FD_Upeak_wrtWpeak_ABP/PPG_D_FD_Upeak_wrtVpeak | The ratio of the ABP/PPG cardiac cycle first derivative Upeak duration with respect to Wpeak to the ABP/PPG cardiac cycle first derivative Upeak duration with respect to Vpeak |
| --- | --- | --- |
| 662 | ABP/PPG_D_RT_FD_Vpeak_wrtWpeak_ABP/PPG_D_FD_Upeak_wrtVpeak | The ratio of the ABP/PPG cardiac cycle first derivative Vpeak duration with respect to Wpeak to the ABP/PPG cardiac cycle first derivative Upeak duration with respect to Vpeak |
| 663 | ABP/PPG_D_RT_FD_Vpeak_wrtWpeak_ABP/PPG_D_FD_Upeak_wrtWpeak | The ratio of the ABP/PPG cardiac cycle first derivative Vpeak duration with respect to Wpeak to the ABP/PPG cardiac cycle first derivative Upeak duration with respect to Wpeak |

22. **Second derivative amplitude features with respect to zero (664-669)**

| 664 | ABP/PPG_SD_NO_ZC | ABP/PPG cardiac cycle second derivative number of zero crossings |
| --- | --- | --- |
| 665 | ABP/PPG_AM_SD_Apeak_wrtzero | ABP/PPG cardiac cycle second derivative Apeak amplitude with respect to zero |
| 666 | ABP/PPG_AM_SD_Bpeak_wrtzero | ABP/PPG cardiac cycle second derivative Bpeak amplitude with respect to zero |
| 667 | ABP/PPG_AM_SD_Cpeak_wrtzero | ABP/PPG cardiac cycle second derivative Cpeak amplitude with respect to zero |
| 668 | ABP/PPG_AM_SD_Dpeak_wrtzero | ABP/PPG cardiac cycle second derivative Dpeak amplitude with respect to zero |
| 669 | ABP/PPG_AM_SD_Epeak_wrtzero | ABP/PPG cardiac cycle second derivative Epeak amplitude with respect to zero |

23. **Second derivative amplitude features with respect to peaks (670-680)**

| 670 | ABP/PPG_AM_SD_Apeak_wrtBpeak | ABP/PPG cardiac cycle second derivative Apeak amplitude with respect to Bpeak |
| --- | --- | --- |
| 671 | ABP/PPG_AM_SD_Apeak_wrtCpeak | ABP/PPG cardiac cycle second derivative Apeak amplitude with respect to Cpeak |
| 672 | ABP/PPG_AM_SD_Apeak_wrtDpeak | ABP/PPG cardiac cycle second derivative Apeak amplitude with respect to Dpeak |
| 673 | ABP/PPG_AM_SD_Apeak_wrtEpeak | ABP/PPG cardiac cycle second derivative Apeak amplitude with respect to Epeak |
| 674 | ABP/PPG_AM_SD_Bpeak_wrtCpeak | ABP/PPG cardiac cycle second derivative Bpeak amplitude with respect to Cpeak |
| 675 | ABP/PPG_AM_SD_Bpeak_wrtDpeak | ABP/PPG cardiac cycle second derivative Bpeak amplitude with respect to Dpeak |
| 676 | ABP/PPG_AM_SD_Bpeak_wrtEpeak | ABP/PPG cardiac cycle second derivative Bpeak amplitude with respect to Epeak |
| 678 | ABP/PPG_AM_SD_Cpeak_wrtDpeak | ABP/PPG cardiac cycle second derivative Cpeak amplitude with respect to Dpeak |
| 679 | ABP/PPG_AM_SD_Cpeak_wrtEpeak | ABP/PPG cardiac cycle second derivative Cpeak amplitude with respect to Epeak |
| 680 | ABP/PPG_AM_SD_Dpeak_wrtEpeak | ABP/PPG cardiac cycle second derivative Dpeak amplitude with respect to Epeak |

24. **Second derivative amplitude ratio features (681-785)**

| 681 | ABP/PPG_AM_RT_SD_Bpeak_wrtzero_ABP/PPG_AM_SD_Apeak_wrtzero | The ratio of the ABP/PPG cardiac cycle second derivative  Bpeak amplitude with respect to zero to the ABP/PPG cardiac cycle second derivative Apeak amplitude with respect to zero |
| --- | --- | --- |
| 682 | ABP/PPG_AM_RT_SD_Cpeak_wrtzero_ABP/PPG_AM_SD_Apeak_wrtzero | The ratio of the ABP/PPG cardiac cycle second derivative  Cpeak amplitude with respect to zero to the ABP/PPG cardiac cycle second derivative Apeak amplitude with respect to zero |
| 683 | ABP/PPG_AM_RT_SD_Dpeak_wrtzero_ABP/PPG_AM_SD_Apeak_wrtzero | The ratio of the ABP/PPG cardiac cycle second derivative  Dpeak amplitude with respect to zero to the ABP/PPG cardiac cycle second derivative Apeak amplitude with respect to zero |
| 684 | ABP/PPG_AM_RT_SD_Epeak_wrtzero_ABP/PPG_AM_SD_Apeak_wrtzero | The ratio of the ABP/PPG cardiac cycle second derivative Epeak amplitude with respect to zero to the ABP/PPG cardiac cycle second derivative Apeak amplitude with respect to zero |
| 685 | ABP/PPG_AM_RT_SD_Apeak_wrtBpeak_ABP/PPG_AM_SD_Apeak_wrtzero | The ratio of the ABP/PPG cardiac cycle second derivative Apeak amplitude with respect to Bpeak to the ABP/PPG cardiac cycle second derivative Apeak amplitude with respect to zero |
| 686 | ABP/PPG_AM_RT_SD_Apeak_wrtCpeak_ABP/PPG_AM_SD_Apeak_wrtzero | The ratio of the ABP/PPG cardiac cycle second derivative Apeak amplitude with respect to Cpeak to the ABP/PPG cardiac cycle second derivative Apeak amplitude with respect to zero |
| 687 | ABP/PPG_AM_RT_SD_Apeak_wrtDpeak_ABP/PPG_AM_SD_Apeak_wrtzero | The ratio of the ABP/PPG cardiac cycle second derivative Apeak amplitude with respect to Dpeak to the ABP/PPG cardiac cycle second derivative Apeak amplitude with respect to zero |
| 688 | ABP/PPG_AM_RT_SD_Apeak_wrtEpeak_ABP/PPG_AM_SD_Apeak_wrtzero | The ratio of the ABP/PPG cardiac cycle second derivative Apeak amplitude with respect to Epeak to the ABP/PPG cardiac cycle second derivative Apeak amplitude with respect to zero |
| 689 | ABP/PPG_AM_RT_SD_Bpeak_wrtCpeak_ABP/PPG_AM_SD_Apeak_wrtzero | The ratio of the ABP/PPG cardiac cycle second derivative Bpeak amplitude with respect to Cpeak to the ABP/PPG cardiac cycle second derivative Apeak amplitude with respect to zero |
| 690 | ABP/PPG_AM_RT_SD_Bpeak_wrtDpeak_ABP/PPG_AM_SD_Apeak_wrtzero | The ratio of the ABP/PPG cardiac cycle second derivative Bpeak amplitude with respect to D peak to the ABP/PPG cardiac cycle second derivative Apeak amplitude with respect to zero |
| 691 | ABP/PPG_AM_RT_SD_Bpeak_wrtEpeak_ABP/PPG_AM_SD_Apeak_wrtzero | The ratio of the ABP/PPG cardiac cycle second derivative Bpeak amplitude with respect to Epeak to the ABP/PPG cardiac cycle second derivative Apeak amplitude with respect to zero |
| 692 | ABP/PPG_AM_RT_SD_Cpeak_wrtDpeak_ABP/PPG_AM_SD_Apeak_wrtzero | The ratio of the ABP/PPG cardiac cycle second derivative Cpeak amplitude with respect to Dpeak to the ABP/PPG cardiac cycle second derivative Apeak amplitude with respect to zero |
| 693 | ABP/PPG_AM_RT_SD_Cpeak_wrtEpeak_ABP/PPG_AM_SD_Apeak_wrtzero | The ratio of the ABP/PPG cardiac cycle second derivative Cpeak amplitude with respect to Epeak to the ABP/PPG cardiac cycle second derivative Apeak amplitude with respect to zero |
| 694 | ABP/PPG_AM_RT_SD_Dpeak_wrtEpeak_ABP/PPG_AM_SD_Apeak_wrtzero | The ratio of the ABP/PPG cardiac cycle second derivative Dpeak amplitude with respect to Epeak to the ABP/PPG cardiac cycle second derivative Apeak amplitude with respect to zero |
| 695 | ABP/PPG_AM_RT_SD_Cpeak_wrtzero_ABP/PPG_AM_SD_Bpeak_wrtzero | The ratio of the ABP/PPG cardiac cycle second derivative Cpeak amplitude with respect to zero to the ABP/PPG cardiac cycle second derivative Bpeak amplitude with respect to zero |
| 696 | ABP/PPG_AM_RT_SD_Dpeak_wrtzero_ABP/PPG_AM_SD_Bpeak_wrtzero | The ratio of the ABP/PPG cardiac cycle second derivative Dpeak amplitude with respect to zero to the ABP/PPG cardiac cycle second derivative Bpeak amplitude with respect to zero |
| 697 | ABP/PPG_AM_RT_SD_Epeak_wrtzero_ABP/PPG_AM_SD_Bpeak_wrtzero | The ratio of the ABP/PPG cardiac cycle second derivative Epeak amplitude with respect to zero to the ABP/PPG cardiac cycle second derivative Bpeak amplitude with respect to zero |
| 698 | ABP/PPG_AM_RT_SD_Apeak_wrtBpeak_ABP/PPG_AM_SD_Bpeak_wrtzero | The ratio of the ABP/PPG cardiac cycle second derivative Apeak amplitude with respect to Bpeak to the ABP/PPG cardiac cycle second derivative Bpeak amplitude with respect to zero |
| 699 | ABP/PPG_AM_RT_SD_Apeak_wrtCpeak_ABP/PPG_AM_SD_Bpeak_wrtzero | The ratio of the ABP/PPG cardiac cycle second derivative Apeak amplitude with respect to Cpeak to the ABP/PPG cardiac cycle second derivative Bpeak amplitude with respect to zero |
| 700 | ABP/PPG_AM_RT_SD_Apeak_wrtDpeak_ABP/PPG_AM_SD_Bpeak_wrtzero | The ratio of the ABP/PPG cardiac cycle second derivative Apeak amplitude with respect to Dpeak to the ABP/PPG cardiac cycle second derivative Bpeak amplitude with respect to zero |
| 701 | ABP/PPG_AM_RT_SD_Apeak_wrtEpeak_ABP/PPG_AM_SD_Bpeak_wrtzero | The ratio of the ABP/PPG cardiac cycle second derivative Apeak amplitude with respect to Epeak to The ratio of the ABP/PPG cardiac cycle second derivative Bpeak amplitude with respect to zero |
| 702 | ABP/PPG_AM_RT_SD_Bpeak_wrtCpeak_ABP/PPG_AM_SD_Bpeak_wrtzero | The ratio of the ABP/PPG cardiac cycle second derivative Bpeak amplitude with respect to Cpeak to the ABP/PPG cardiac cycle second derivative Bpeak amplitude with respect to zero |
| 703 | ABP/PPG_AM_RT_SD_Bpeak_wrtDpeak_ABP/PPG_AM_SD_Bpeak_wrtzero | The ratio of the ABP/PPG cardiac cycle second derivative Bpeak amplitude with respect to Dpeak to the ABP/PPG cardiac cycle second derivative Bpeak amplitude with respect to zero |
| 704 | ABP/PPG_AM_RT_SD_Bpeak_wrtEpeak_ABP/PPG_AM_SD_Bpeak_wrtzero | The ratio of the ABP/PPG cardiac cycle second derivative Bpeak amplitude with respect to Epeak to the ABP/PPG cardiac cycle second derivative Bpeak amplitude with respect to zero |
| 705 | ABP/PPG_AM_RT_SD_Cpeak_wrtDpeak_ABP/PPG_AM_SD_Bpeak_wrtzero | The ratio of the ABP/PPG cardiac cycle second derivative Cpeak amplitude with respect to Dpeak to the ABP/PPG cardiac cycle second derivative Bpeak amplitude with respect to zero |
| 706 | ABP/PPG_AM_RT_SD_Cpeak_wrtEpeak_ABP/PPG_AM_SD_Bpeak_wrtzero | The ratio of the ABP/PPG cardiac cycle second derivative Cpeak amplitude with respect to Epeak to the ABP/PPG cardiac cycle second derivative Bpeak amplitude with respect to zero |
| 707 | ABP/PPG_AM_RT_SD_Dpeak_wrtEpeak_ABP/PPG_AM_SD_Bpeak_wrtzero | The ratio of the ABP/PPG cardiac cycle second derivative Dpeak amplitude with respect to Epeak to the ABP/PPG cardiac cycle second derivative Bpeak amplitude with respect to zero |
| 708 | ABP/PPG_AM_RT_SD_Dpeak_wrtzero_ABP/PPG_AM_SD_Cpeak_wrtzero | The ratio of the ABP/PPG cardiac cycle second derivative Dpeak amplitude with respect to zero to the ABP/PPG cardiac cycle second derivative Cpeak amplitude with respect to zero |
| 709 | ABP/PPG_AM_RT_SD_Epeak_wrtzero_ABP/PPG_AM_SD_Cpeak_wrtzero | The ratio of the ABP/PPG cardiac cycle second derivative Epeak amplitude with respect to zero to the ABP/PPG cardiac cycle second derivative Cpeak amplitude with respect to zero |
| 710 | ABP/PPG_AM_RT_SD_Apeak_wrtBpeak_ABP/PPG_AM_SD_Cpeak_wrtzero | The ratio of the ABP/PPG cardiac cycle second derivative Apeak amplitude with respect to Bpeak to the ABP/PPG cardiac cycle second derivative Cpeak amplitude with respect to zero |
| 711 | ABP/PPG_AM_RT_SD_Apeak_wrtCpeak_ABP/PPG_AM_SD_Cpeak_wrtzero | The ratio of the ABP/PPG cardiac cycle second derivative Apeak amplitude with respect to Cpeak to the ABP/PPG cardiac cycle second derivative Cpeak amplitude with respect to zero |
| 712 | ABP/PPG_AM_RT_SD_Apeak_wrtDpeak_ABP/PPG_AM_SD_Cpeak_wrtzero | The ratio of the ABP/PPG cardiac cycle second derivative Apeak amplitude with respect to Dpeak to the ABP/PPG cardiac cycle second derivative Cpeak amplitude with respect to zero |
| 713 | ABP/PPG_AM_RT_SD_Apeak_wrtEpeak_ABP/PPG_AM_SD_Cpeak_wrtzero | The ratio of the ABP/PPG cardiac cycle second derivative Apeak amplitude with respect to Epeak to the ABP/PPG cardiac cycle second derivative Cpeak amplitude with respect to zero |
| 714 | ABP/PPG_AM_RT_SD_Bpeak_wrtCpeak_ABP/PPG_AM_SD_Cpeak_wrtzero | The ratio of the ABP/PPG cardiac cycle second derivative Bpeak amplitude with respect to Cpeak to the ABP/PPG cardiac cycle second derivative Cpeak amplitude with respect to zero |
| 715 | ABP/PPG_AM_RT_SD_Bpeak_wrtDpeak_ABP/PPG_AM_SD_Cpeak_wrtzero | The ratio of the ABP/PPG cardiac cycle second derivative Bpeak amplitude with respect to Dpeak to the ABP/PPG cardiac cycle second derivative Cpeak amplitude with respect to zero |
| 716 | ABP/PPG_AM_RT_SD_Bpeak_wrtEpeak_ABP/PPG_AM_SD_Cpeak_wrtzero | The ratio of the ABP/PPG cardiac cycle second derivative Bpeak amplitude with respect to Epeak to the ABP/PPG cardiac cycle second derivative Cpeak amplitude with respect to zero |
| 717 | ABP/PPG_AM_RT_SD_Cpeak_wrtDpeak_ABP/PPG_AM_SD_Cpeak_wrtzero | The ratio of the ABP/PPG cardiac cycle second derivative Cpeak amplitude with respect to D peak to the ABP/PPG cardiac cycle second derivative Cpeak amplitude with respect to zero |
| 718 | ABP/PPG_AM_RT_SD_Cpeak_wrtEpeak_ABP/PPG_AM_SD_Cpeak_wrtzero | The ratio of the ABP/PPG cardiac cycle second derivative Cpeak amplitude with respect to Epeak to the ABP/PPG cardiac cycle second derivative Cpeak amplitude with respect to zero |
| 719 | ABP/PPG_AM_RT_SD_Dpeak_wrtEpeak_ABP/PPG_AM_SD_Cpeak_wrtzero | The ratio of the ABP/PPG cardiac cycle second derivative Dpeak amplitude with respect to Epeak to the ABP/PPG cardiac cycle second derivative Cpeak amplitude with respect to zero |
| 720 | ABP/PPG_AM_RT_SD_Epeak_wrtzero_ABP/PPG_AM_SD_Dpeak_wrtzero | The ratio of the ABP/PPG cardiac cycle second derivative Epeak amplitude with respect to zero to the ABP/PPG cardiac cycle second derivative Dpeak amplitude with respect to zero |
| 721 | ABP/PPG_AM_RT_SD_Apeak_wrtBpeak_ABP/PPG_AM_SD_Dpeak_wrtzero | The ratio of the ABP/PPG cardiac cycle second derivative Apeak amplitude with respect to Bpeak to the ABP/PPG cardiac cycle second derivative Dpeak amplitude with respect to zero |
| 722 | ABP/PPG_AM_RT_SD_Apeak_wrtCpeak_ABP/PPG_AM_SD_Dpeak_wrtzero | The ratio of the ABP/PPG cardiac cycle second derivative Apeak amplitude with respect to Cpeak to the ABP/PPG cardiac cycle second derivative Dpeak amplitude with respect to zero |
| 723 | ABP/PPG_AM_RT_SD_Apeak_wrtDpeak_ABP/PPG_AM_SD_Dpeak_wrtzero | The ratio of the ABP/PPG cardiac cycle second derivative Apeak amplitude with respect to Dpeak to the ABP/PPG cardiac cycle second derivative Dpeak amplitude with respect to zero |
| 724 | ABP/PPG_AM_RT_SD_Apeak_wrtEpeak_ABP/PPG_AM_SD_Dpeak_wrtzero | The ratio of the ABP/PPG cardiac cycle second derivative Apeak amplitude with respect to Epeak to the ABP/PPG cardiac cycle second derivative Dpeak amplitude with respect to zero |
| 725 | ABP/PPG_AM_RT_SD_Bpeak_wrtCpeak_ABP/PPG_AM_SD_Dpeak_wrtzero | The ratio of the ABP/PPG cardiac cycle second derivative Bpeak amplitude with respect to Cpeak to the ABP/PPG cardiac cycle second derivative Dpeak amplitude with respect to zero |
| 726 | ABP/PPG_AM_RT_SD_Bpeak_wrtDpeak_ABP/PPG_AM_SD_Dpeak_wrtzero | The ratio of the ABP/PPG cardiac cycle second derivative Bpeak amplitude with respect to Dpeak to the ABP/PPG cardiac cycle second derivative Dpeak amplitude with respect to zero |
| 727 | ABP/PPG_AM_RT_SD_Bpeak_wrtEpeak_ABP/PPG_AM_SD_Dpeak_wrtzero | The ratio of the ABP/PPG cardiac cycle second derivative Bpeak amplitude with respect to Epeak to the ABP/PPG cardiac cycle second derivative Dpeak amplitude with respect to zero |
| 728 | ABP/PPG_AM_RT_SD_Cpeak_wrtDpeak_ABP/PPG_AM_SD_Dpeak_wrtzero | The ratio of the ABP/PPG cardiac cycle second derivative Cpeak amplitude with respect to Dpeak to the ABP/PPG cardiac cycle second derivative Dpeak amplitude with respect to zero |
| 729 | ABP/PPG_AM_RT_SD_Cpeak_wrtEpeak_ABP/PPG_AM_SD_Dpeak_wrtzero | The ratio of the ABP/PPG cardiac cycle second derivative Cpeak amplitude with respect to Epeak to the ABP/PPG cardiac cycle second derivative Dpeak amplitude with respect to zero |
| 730 | ABP/PPG_AM_RT_SD_Dpeak_wrtEpeak_ABP/PPG_AM_SD_Dpeak_wrtzero | The ratio of the ABP/PPG cardiac cycle second derivative Dpeak amplitude with respect to Epeak to the ABP/PPG cardiac cycle second derivative Dpeak amplitude with respect to zero |
| 731 | ABP/PPG_AM_RT_SD_Apeak_wrtBpeak_ABP/PPG_AM_SD_Epeak_wrtzero | The ratio of the ABP/PPG cardiac cycle second derivative Apeak amplitude with respect to Bpeak to the ABP/PPG cardiac cycle second derivative Epeak amplitude with respect to zero |
| 732 | ABP/PPG_AM_RT_SD_Apeak_wrtCpeak_ABP/PPG_AM_SD_Epeak_wrtzero | The ratio of the ABP/PPG cardiac cycle second derivative Apeak amplitude with respect to Cpeak to the ABP/PPG cardiac cycle second derivative Epeak amplitude with respect to zero |
| 733 | ABP/PPG_AM_RT_SD_Apeak_wrtDpeak_ABP/PPG_AM_SD_Epeak_wrtzero | The ratio of the ABP/PPG cardiac cycle second derivative Apeak amplitude with respect to Dpeak to the ABP/PPG cardiac cycle second derivative Epeak amplitude with respect to zero |
| 734 | ABP/PPG_AM_RT_SD_Apeak_wrtEpeak_ABP/PPG_AM_SD_Epeak_wrtzero | The ratio of the ABP/PPG cardiac cycle second derivative Apeak amplitude with respect to Epeak to the ABP/PPG cardiac cycle second derivative Epeak amplitude with respect to zero |
| 735 | ABP/PPG_AM_RT_SD_Bpeak_wrtCpeak_ABP/PPG_AM_SD_Epeak_wrtzero | The ratio of the ABP/PPG cardiac cycle second derivative Bpeak amplitude with respect to Cpeak to the ABP/PPG cardiac cycle second derivative Epeak amplitude with respect to zero |
| 736 | ABP/PPG_AM_RT_SD_Bpeak_wrtDpeak_ABP/PPG_AM_SD_Epeak_wrtzero | The ratio of the ABP/PPG cardiac cycle second derivative Bpeak amplitude with respect to Dpeak to second derviative of PPG cardiac cycle Epeak amplitude with respect to zero |
| 737 | ABP/PPG_AM_RT_SD_Bpeak_wrtEpeak_ABP/PPG_AM_SD_Epeak_wrtzero | The ratio of the ABP/PPG cardiac cycle second derivative Bpeak amplitude with respect to Epeak to the ABP/PPG cardiac cycle second derivative Epeak amplitude with respect to zero |
| 738 | ABP/PPG_AM_RT_SD_Cpeak_wrtDpeak_ABP/PPG_AM_SD_Epeak_wrtzero | The ratio of the ABP/PPG cardiac cycle second derivative Cpeak amplitude with respect to Dpeak to the ABP/PPG cardiac cycle second derivative Epeak amplitude with respect to zero |
| 739 | ABP/PPG_AM_RT_SD_Cpeak_wrtEpeak_ABP/PPG_AM_SD_Epeak_wrtzero | The ratio of the ABP/PPG cardiac cycle second derivative Cpeak amplitude with respect to Epeak to the ABP/PPG cardiac cycle second derivative Epeak amplitude with respect to zero |
| 740 | ABP/PPG_AM_RT_SD_Dpeak_wrtEpeak_ABP/PPG_AM_SD_Epeak_wrtzero | The ratio of the ABP/PPG cardiac cycle second derivative Dpeak amplitude with respect to Epeak to the ABP/PPG cardiac cycle second derivative Epeak amplitude with respect to zero |
| 741 | ABP/PPG_AM_RT_SD_Apeak_wrtCpeak_ABP/PPG_AM_SD_Apeak_wrtBpeak | The ratio of the ABP/PPG cardiac cycle second derivative Apeak amplitude with respect to Cpeak to the ABP/PPG cardiac cycle second derivative Apeak amplitude with respect to Bpeak |
| 742 | ABP/PPG_AM_RT_SD_Apeak_wrtDpeak_ABP/PPG_AM_SD_Apeak_wrtBpeak | The ratio of the ABP/PPG cardiac cycle second derivative Apeak amplitude with respect to Dpeak to the ABP/PPG cardiac cycle second derivative Apeak amplitude with respect to Bpeak |
| 743 | ABP/PPG_AM_RT_SD_Apeak_wrtEpeak_ABP/PPG_AM_SD_Apeak_wrtBpeak | The ratio of the ABP/PPG cardiac cycle second derivative Apeak amplitude with respect to Epeak to the ABP/PPG cardiac cycle second derivative Apeak amplitude with respect to Bpeak |
| 744 | ABP/PPG_AM_RT_SD_Bpeak_wrtCpeak_ABP/PPG_AM_SD_Apeak_wrtBpeak | The ratio of the ABP/PPG cardiac cycle second derivative Bpeak amplitude with respect to Cpeak to the ABP/PPG cardiac cycle second derivative Apeak amplitude with respect to Bpeak |
| 745 | ABP/PPG_AM_RT_SD_Bpeak_wrtDpeak_ABP/PPG_AM_SD_Apeak_wrtBpeak | The ratio of the ABP/PPG cardiac cycle second derivative Bpeak amplitude with respect to Dpeak to the ABP/PPG cardiac cycle second derivative Apeak amplitude with respect to Bpeak |
| 746 | ABP/PPG_AM_RT_SD_Bpeak_wrtEpeak_ABP/PPG_AM_SD_Apeak_wrtBpeak | The ratio of the ABP/PPG cardiac cycle second derivative Bpeak amplitude with respect to Epeak to the ABP/PPG cardiac cycle second derivative Apeak amplitude with respect to Bpeak |
| 747 | ABP/PPG_AM_RT_SD_Cpeak_wrtDpeak_ABP/PPG_AM_SD_Apeak_wrtBpeak | The ratio of the ABP/PPG cardiac cycle second derivative Cpeak amplitude with respect to Dpeak to the ABP/PPG cardiac cycle second derivative Apeak amplitude with respect to Bpeak |
| 748 | ABP/PPG_AM_RT_SD_Cpeak_wrtEpeak_ABP/PPG_AM_SD_Apeak_wrtBpeak | The ratio of the ABP/PPG cardiac cycle second derivative Cpeak amplitude with respect to Epeak to the ABP/PPG cardiac cycle second derivative Apeak amplitude with respect to Bpeak |
| 749 | ABP/PPG_AM_RT_SD_Dpeak_wrtEpeak_ABP/PPG_AM_SD_Apeak_wrtBpeak | The ratio of the ABP/PPG cardiac cycle second derivative Dpeak amplitude with respect to Epeak to the ABP/PPG cardiac cycle second derivative Apeak amplitude with respect to Bpeak |
| 750 | ABP/PPG_AM_RT_SD_Apeak_wrtDpeak_ABP/PPG_AM_SD_Apeak_wrtCpeak | The ratio of the ABP/PPG cardiac cycle second derivative Apeak amplitude with respect to Dpeak to the ABP/PPG cardiac cycle second derivative Apeak amplitude with respect to Cpeak |
| 751 | ABP/PPG_AM_RT_SD_Apeak_wrtEpeak_ABP/PPG_AM_SD_Apeak_wrtCpeak | The ratio of the ABP/PPG cardiac cycle second derivative Apeak amplitude with respect to Epeak to the ABP/PPG cardiac cycle second derivative Apeak amplitude with respect to Cpeak |
| 752 | ABP/PPG_AM_RT_SD_Bpeak_wrtCpeak_ABP/PPG_AM_SD_Apeak_wrtCpeak | The ratio of the ABP/PPG cardiac cycle second derivative Bpeak amplitude with respect to Cpeak to the ABP/PPG cardiac cycle second derivative Apeak amplitude with respect to Cpeak |
| 753 | ABP/PPG_AM_RT_SD_Bpeak_wrtDpeak_ABP/PPG_AM_SD_Apeak_wrtCpeak | The ratio of the ABP/PPG cardiac cycle second derivative Bpeak amplitude with respect to Dpeak to the ABP/PPG cardiac cycle second derivative Apeak amplitude with respect to Cpeak |
| 754 | ABP/PPG_AM_RT_SD_Bpeak_wrtEpeak_ABP/PPG_AM_SD_Apeak_wrtCpeak | The ratio of the ABP/PPG cardiac cycle second derivative Bpeak amplitude with respect to Epeak to the ABP/PPG cardiac cycle second derivative Apeak amplitude with respect to Cpeak |
| 755 | ABP/PPG_AM_RT_SD_Cpeak_wrtDpeak_ABP/PPG_AM_SD_Apeak_wrtCpeak | The ratio of the ABP/PPG cardiac cycle second derivative Cpeak amplitude with respect to Dpeak to the ABP/PPG cardiac cycle second derivative Apeak amplitude with respect to Cpeak |
| 756 | ABP/PPG_AM_RT_SD_Cpeak_wrtEpeak_ABP/PPG_AM_SD_Apeak_wrtCpeak | The ratio of the ABP/PPG cardiac cycle second derivative Cpeak amplitude with respect to Epeak to the ABP/PPG cardiac cycle second derivative Apeak amplitude with respect to Cpeak |
| 757 | ABP/PPG_AM_RT_SD_Dpeak_wrtEpeak_ABP/PPG_AM_SD_Apeak_wrtCpeak | The ratio of the ABP/PPG cardiac cycle second derivative Dpeak amplitude with respect to Epeak to the ABP/PPG cardiac cycle second derivative Apeak amplitude with respect to Cpeak |
| 758 | ABP/PPG_AM_RT_SD_Apeak_wrtEpeak_ABP/PPG_AM_SD_Apeak_wrtDpeak | The ratio of the ABP/PPG cardiac cycle second derivative Apeak amplitude with respect to Epeak to the ABP/PPG cardiac cycle second derivative Apeak amplitude with respect to Dpeak |
| 759 | ABP/PPG_AM_RT_SD_Bpeak_wrtCpeak_ABP/PPG_AM_SD_Apeak_wrtDpeak | The ratio of the ABP/PPG cardiac cycle second derivative Bpeak amplitude with respect to Cpeak to the ABP/PPG cardiac cycle second derivative Apeak amplitude with respect to Dpeak |
| 760 | ABP/PPG_AM_RT_SD_Bpeak_wrtDpeak_ABP/PPG_AM_SD_Apeak_wrtDpeak | The ratio of the ABP/PPG cardiac cycle second derivative Bpeak amplitude with respect to Dpeak to the ABP/PPG cardiac cycle second derivative Apeak amplitude with respect to Dpeak |
| 761 | ABP/PPG_AM_RT_SD_Bpeak_wrtEpeak_ABP/PPG_AM_SD_Apeak_wrtDpeak | The ratio of the ABP/PPG cardiac cycle second derivative Bpeak amplitude with respect to Epeak to the ABP/PPG cardiac cycle second derivative Apeak amplitude with respect to Dpeak |
| 762 | ABP/PPG_AM_RT_SD_Cpeak_wrtDpeak_ABP/PPG_AM_SD_Apeak_wrtDpeak | The ratio of the ABP/PPG cardiac cycle second derivative Cpeak amplitude with respect to Dpeak to the ABP/PPG cardiac cycle second derivative Apeak amplitude with respect to Dpeak |
| 763 | ABP/PPG_AM_RT_SD_Cpeak_wrtEpeak_ABP/PPG_AM_SD_Apeak_wrtDpeak | The ratio of the ABP/PPG cardiac cycle second derivative Cpeak amplitude with respect to Epeak to the ABP/PPG cardiac cycle second derivative Apeak amplitude with respect to Dpeak |
| 764 | ABP/PPG_AM_RT_SD_Dpeak_wrtEpeak_ABP/PPG_AM_SD_Apeak_wrtDpeak | The ratio of the ABP/PPG cardiac cycle second derivative Dpeak amplitude with respect to Epeak to the ABP/PPG cardiac cycle second derivative Apeak amplitude with respect to Dpeak |
| 765 | ABP/PPG_AM_RT_SD_Bpeak_wrtCpeak_ABP/PPG_AM_SD_Apeak_wrtEpeak | The ratio of the ABP/PPG cardiac cycle second derivative Bpeak amplitude with respect to Cpeak to the ABP/PPG cardiac cycle second derivative Apeak amplitude with respect to Epeak |
| 766 | ABP/PPG_AM_RT_SD_Bpeak_wrtDpeak_ABP/PPG_AM_SD_Apeak_wrtEpeak | The ratio of the ABP/PPG cardiac cycle second derivative Bpeak amplitude with respect to Dpeak to the ABP/PPG cardiac cycle second derivative Apeak amplitude with respect to Epeak |
| 767 | ABP/PPG_AM_RT_SD_Bpeak_wrtEpeak_ABP/PPG_AM_SD_Apeak_wrtEpeak | The ratio of the ABP/PPG cardiac cycle second derivative Bpeak amplitude with respect to Epeak to the ABP/PPG cardiac cycle second derivative Apeak amplitude with respect to Epeak |
| 768 | ABP/PPG_AM_RT_SD_Cpeak_wrtDpeak_ABP/PPG_AM_SD_Apeak_wrtEpeak | The ratio of the ABP/PPG cardiac cycle second derivative Cpeak amplitude with respect to Dpeak to the ABP/PPG cardiac cycle second derivative Apeak amplitude with respect to Epeak |
| 769 | ABP/PPG_AM_RT_SD_Cpeak_wrtEpeak_ABP/PPG_AM_SD_Apeak_wrtEpeak | The ratio of the ABP/PPG cardiac cycle second derivative Cpeak amplitude with respect to Epeak to the ABP/PPG cardiac cycle second derivative Apeak amplitude with respect to Epeak |
| 770 | ABP/PPG_AM_RT_SD_Dpeak_wrtEpeak_ABP/PPG_AM_SD_Apeak_wrtEpeak | The ratio of the ABP/PPG cardiac cycle second derivative Dpeak amplitude with respect to Epeak to the ABP/PPG cardiac cycle second derivative Apeak amplitude with respect to Epeak |
| 771 | ABP/PPG_AM_RT_SD_Bpeak_wrtDpeak_ABP/PPG_AM_SD_Bpeak_wrtCpeak | The ratio of the ABP/PPG cardiac cycle second derivative Bpeak amplitude with respect to Dpeak to the ABP/PPG cardiac cycle second derivative Bpeak amplitude with respect to Cpeak |
| 772 | ABP/PPG_AM_RT_SD_Bpeak_wrtEpeak_ABP/PPG_AM_SD_Bpeak_wrtCpeak | The ratio of the ABP/PPG cardiac cycle second derivative Bpeak amplitude with respect to Epeak to the ABP/PPG cardiac cycle second derivative Bpeak amplitude with respect to Cpeak |
| 773 | ABP/PPG_AM_RT_SD_Cpeak_wrtDpeak_ABP/PPG_AM_SD_Bpeak_wrtCpeak | The ratio of the ABP/PPG cardiac cycle second derivative Cpeak amplitude with respect to Dpeak to the ABP/PPG cardiac cycle second derivative Bpeak amplitude with respect to Cpeak |
| 774 | ABP/PPG_AM_RT_SD_Cpeak_wrtEpeak_ABP/PPG_AM_SD_Bpeak_wrtCpeak | The ratio of the ABP/PPG cardiac cycle second derivative Cpeak amplitude with respect to Epeak to the ABP/PPG cardiac cycle second derivative Bpeak amplitude with respect to Cpeak |
| 775 | ABP/PPG_AM_RT_SD_Dpeak_wrtEpeak_ABP/PPG_AM_SD_Bpeak_wrtCpeak | The ratio of the ABP/PPG cardiac cycle second derivative Dpeak amplitude with respect to Epeak to the ABP/PPG cardiac cycle second derivative Bpeak amplitude with respect to Cpeak |
| 776 | ABP/PPG_AM_RT_SD_Bpeak_wrtEpeak_ABP/PPG_AM_SD_Bpeak_wrtDpeak | The ratio of the ABP/PPG cardiac cycle second derivative Bpeak amplitude with respect to Epeak to the ABP/PPG cardiac cycle second derivative Bpeak amplitude with respect to Dpeak |
| 777 | ABP/PPG_AM_RT_SD_Cpeak_wrtDpeak_ABP/PPG_AM_SD_Bpeak_wrtDpeak | The ratio of the ABP/PPG cardiac cycle second derivative Cpeak amplitude with respect to Dpeak to the ABP/PPG cardiac cycle second derivative Bpeak amplitude with respect to Dpeak |
| 778 | ABP/PPG_AM_RT_SD_Cpeak_wrtEpeak_ABP/PPG_AM_SD_Bpeak_wrtDpeak | The ratio of the ABP/PPG cardiac cycle second derivative Cpeak amplitude with respect to Epeak to the ABP/PPG cardiac cycle second derivative Bpeak amplitude with respect to Dpeak |
| 779 | ABP/PPG_AM_RT_SD_Dpeak_wrtEpeak_ABP/PPG_AM_SD_Bpeak_wrtDpeak | The ratio of the ABP/PPG cardiac cycle second derivative Dpeak amplitude with respect to Epeak to the ABP/PPG cardiac cycle second derivative Bpeak amplitude with respect to Dpeak |
| 780 | ABP/PPG_AM_RT_SD_Cpeak_wrtDpeak_ABP/PPG_AM_SD_Bpeak_wrtEpeak | The ratio of the ABP/PPG cardiac cycle second derivative Cpeak amplitude with respect to Dpeak to the ABP/PPG cardiac cycle second derivative Bpeak amplitude with respect to Epeak |
| 781 | ABP/PPG_AM_RT_SD_Cpeak_wrtEpeak_ABP/PPG_AM_SD_Bpeak_wrtEpeak | The ratio of the ABP/PPG cardiac cycle second derivative Cpeak amplitude with respect to Epeak to the ABP/PPG cardiac cycle second derivative Bpeak amplitude with respect to Epeak |
| 782 | ABP/PPG_AM_RT_SD_Dpeak_wrtEpeak_ABP/PPG_AM_SD_Bpeak_wrtEpeak | The ratio of the ABP/PPG cardiac cycle second derivative Dpeak amplitude with respect to Epeak to the ABP/PPG cardiac cycle second derivative Bpeak amplitude with respect to Epeak |
| 783 | ABP/PPG_AM_RT_SD_Cpeak_wrtEpeak_ABP/PPG_AM_SD_Cpeak_wrtDpeak | The ratio of the ABP/PPG cardiac cycle second derivative Cpeak amplitude with respect to Epeak to the ABP/PPG cardiac cycle second derivative Cpeak amplitude with respect to Dpeak |
| 784 | ABP/PPG_AM_RT_SD_Dpeak_wrtEpeak_ABP/PPG_AM_SD_Cpeak_wrtDpeak | The ratio of the ABP/PPG cardiac cycle second derivative Dpeak amplitude with respect to Epeak to the ABP/PPG cardiac cycle second derivative Cpeak amplitude with respect to Dpeak |
| 785 | ABP/PPG_AM_RT_SD_Dpeak_wrtEpeak_ABP/PPG_AM_SD_Cpeak_wrtEpeak | The ratio of the ABP/PPG cardiac cycle second derivative Dpeak amplitude with respect to Epeak to the ABP/PPG cardiac cycle second derivative Cpeak amplitude with respect to Epeak |

25. **Second derivative duration features (786-795)**

| 786 | ABP/PPG_D_SD_Apeak_wrtBpeak | Duration of the ABP/PPG cardiac cycle second derivative Apeak duration with respect to Bpeak |
| --- | --- | --- |
| 787 | ABP/PPG_D_SD_Apeak_wrtCpeak | Duration of the ABP/PPG cardiac cycle second derivative Apeak duration with respect to Cpeak |
| 788 | ABP/PPG_D_SD_Apeak_wrtDpeak | Duration of the ABP/PPG cardiac cycle second derivative Apeak duration with respect to Dpeak |
| 789 | ABP/PPG_D_SD_Apeak_wrtEpeak | Duration of the ABP/PPG cardiac cycle second derivative Apeak duration with respect to Epeak |
| 790 | ABP/PPG_D_SD_Bpeak_wrtCpeak | Duration of the ABP/PPG cardiac cycle second derivative Bpeak duration with respect to Cpeak |
| 791 | ABP/PPG_D_SD_Bpeak_wrtDpeak | Duration of the ABP/PPG cardiac cycle second derivative Bpeak duration with respect to Dpeak |
| 792 | ABP/PPG_D_SD_Bpeak_wrtEpeak | Duration of the ABP/PPG cardiac cycle second derivative Bpeak duration with respect to Epeak |
| 793 | ABP/PPG_D_SD_Cpeak_wrtDpeak | Duration of the ABP/PPG cardiac cycle second derivative Cpeak duration with respect to Dpeak |
| 794 | ABP/PPG_D_SD_Cpeak_wrtEpeak | Duration of the ABP/PPG cardiac cycle second derivative Cpeak duration with respect to Epeak |
| 795 | ABP/PPG_D_SD_Dpeak_wrtEpeak | Duration of the ABP/PPG cardiac cycle second derivative Dpeak duration with respect to Epeak |

26. **Second derivative duration ratio features (796-840)**

| 796 | ABP/PPG_D_RT_SD_Apeak_wrtCpeak_ABP/PPG_D_SD_Apeak_wrtBpeak | The ratio of the ABP/PPG cardiac cycle second derivative Apeak duration with respect to Cpeak to the ABP/PPG cardiac cycle second derivative Apeak duration with respect to Bpeak |
| --- | --- | --- |
| 797 | ABP/PPG_D_RT_SD_Apeak_wrtDpeak_ABP/PPG_D_SD_Apeak_wrtBpeak | The ratio of the ABP/PPG cardiac cycle second derivative Apeak duration with respect to Dpeak to the ABP/PPG cardiac cycle second derivative Apeak duration with respect to Bpeak |
| 798 | ABP/PPG_D_RT_SD_Apeak_wrtEpeak_ABP/PPG_D_SD_Apeak_wrtBpeak | The ratio of the ABP/PPG cardiac cycle second derivative Apeak duration with respect to Epeak to the ABP/PPG cardiac cycle second derivative Apeak duration with respect to Bpeak |
| 799 | ABP/PPG_D_RT_SD_Bpeak_wrtCpeak_ABP/PPG_D_SD_Apeak_wrtBpeak | The ratio of the ABP/PPG cardiac cycle second derivative Bpeak duration with respect to Cpeak to the ABP/PPG cardiac cycle second derivative Apeak duration with respect to Bpeak |
| 800 | ABP/PPG_D_RT_SD_Bpeak_wrtDpeak_ABP/PPG_D_SD_Apeak_wrtBpeak | The ratio of the ABP/PPG cardiac cycle second derivative Bpeak duration with respect to Dpeak to the ABP/PPG cardiac cycle second derivative Apeak duration with respect to Bpeak |
| 801 | ABP/PPG_D_RT_SD_Bpeak_wrtEpeak_ABP/PPG_D_SD_Apeak_wrtBpeak | The ratio of the ABP/PPG cardiac cycle second derivative Bpeak duration with respect to Epeak to the ABP/PPG cardiac cycle second derivative Apeak duration with respect to Bpeak |
| 802 | ABP/PPG_D_RT_SD_Cpeak_wrtDpeak_ABP/PPG_D_SD_Apeak_wrtBpeak | The ratio of the ABP/PPG cardiac cycle second derivative Cpeak duration with respect to Dpeak to the ABP/PPG cardiac cycle second derivative Apeak duration with respect to Bpeak |
| 803 | ABP/PPG_D_RT_SD_Cpeak_wrtEpeak_ABP/PPG_D_SD_Apeak_wrtBpeak | The ratio of the ABP/PPG cardiac cycle second derivative Cpeak duration with respect to Epeak to the ABP/PPG cardiac cycle second derivative Apeak duration with respect to Bpeak |
| 804 | ABP/PPG_D_RT_SD_Dpeak_wrtEpeak_ABP/PPG_D_SD_Apeak_wrtBpeak | The ratio of the ABP/PPG cardiac cycle second derivative Dpeak duration with respect to Epeak to the ABP/PPG cardiac cycle second derivative Apeak duration with respect to Bpeak |
| 805 | ABP/PPG_D_RT_SD_Apeak_wrtDpeak_ABP/PPG_D_SD_Apeak_wrtCpeak | The ratio of the ABP/PPG cardiac cycle second derivative Apeak duration with respect to Dpeak to the ABP/PPG cardiac cycle second derivative Apeak duration with respect to Cpeak |
| 806 | ABP/PPG_D_RT_SD_Apeak_wrtEpeak_ABP/PPG_D_SD_Apeak_wrtCpeak | The ratio of the ABP/PPG cardiac cycle second derivative Apeak duration with respect to Epeak to the ABP/PPG cardiac cycle second derivative Apeak duration with respect to Cpeak |
| 807 | ABP/PPG_D_RT_SD_Bpeak_wrtCpeak_ABP/PPG_D_SD_Apeak_wrtCpeak | The ratio of the ABP/PPG cardiac cycle second derivative Bpeak duration with respect to Cpeak to the ABP/PPG cardiac cycle second derivative Apeak duration with respect to Cpeak |
| 808 | ABP/PPG_D_RT_SD_Bpeak_wrtDpeak_ABP/PPG_D_SD_Apeak_wrtCpeak | The ratio of the ABP/PPG cardiac cycle second derivative Bpeak duration with respect to Dpeak to the ABP/PPG cardiac cycle second derivative Apeak duration with respect to Cpeak |
| 809 | ABP/PPG_D_RT_SD_Bpeak_wrtEpeak_ABP/PPG_D_SD_Apeak_wrtCpeak | The ratio of the ABP/PPG cardiac cycle second derivative Bpeak duration with respect to Epeak to the ABP/PPG cardiac cycle second derivative Apeak duration with respect to Cpeak |
| 810 | ABP/PPG_D_RT_SD_Cpeak_wrtDpeak_ABP/PPG_D_SD_Apeak_wrtCpeak | The ratio of the ABP/PPG cardiac cycle second derivative Cpeak duration with respect to Dpeak to the ABP/PPG cardiac cycle second derivative Apeak duration with respect to Cpeak |
| 811 | ABP/PPG_D_RT_SD_Cpeak_wrtEpeak_ABP/PPG_D_SD_Apeak_wrtCpeak | The ratio of the ABP/PPG cardiac cycle second derivative Cpeak duration with respect to Epeak to the ABP/PPG cardiac cycle second derivative Apeak duration with respect to Cpeak |
| 812 | ABP/PPG_D_RT_SD_Dpeak_wrtEpeak_ABP/PPG_D_SD_Apeak_wrtCpeak | The ratio of the ABP/PPG cardiac cycle second derivative Dpeak duration with respect to Epeak to the ABP/PPG cardiac cycle second derivative Apeak duration with respect to Cpeak |
| 813 | ABP/PPG_D_RT_SD_Apeak_wrtEpeak_ABP/PPG_D_SD_Apeak_wrtDpeak | The ratio of the ABP/PPG cardiac cycle second derivative Apeak duration with respect to Epeak to the ABP/PPG cardiac cycle second derivative Apeak duration with respect to Dpeak |
| 814 | ABP/PPG_D_RT_SD_Bpeak_wrtCpeak_ABP/PPG_D_SD_Apeak_wrtDpeak | The ratio of the ABP/PPG cardiac cycle second derivative Bpeak duration with respect to Cpeak to the ABP/PPG cardiac cycle second derivative Apeak duration with respect to Dpeak |
| 815 | ABP/PPG_D_RT_SD_Bpeak_wrtDpeak_ABP/PPG_D_SD_Apeak_wrtDpeak | The ratio of the ABP/PPG cardiac cycle second derivative Bpeak duration with respect to Dpeak to the ABP/PPG cardiac cycle second derivative Apeak duration with respect to Dpeak |
| 816 | ABP/PPG_D_RT_SD_Bpeak_wrtEpeak_ABP/PPG_D_SD_Apeak_wrtDpeak | The ratio of the ABP/PPG cardiac cycle second derivative Bpeak duration with respect to Epeak to the ABP/PPG cardiac cycle second derivative Apeak duration with respect to Dpeak |
| 817 | ABP/PPG_D_RT_SD_Cpeak_wrtDpeak_ABP/PPG_D_SD_Apeak_wrtDpeak | The ratio of the ABP/PPG cardiac cycle second derivative Cpeak duration with respect to Dpeak to the ABP/PPG cardiac cycle second derivative Apeak duration with respect to Dpeak |
| 818 | ABP/PPG_D_RT_SD_Cpeak_wrtEpeak_ABP/PPG_D_SD_Apeak_wrtDpeak | The ratio of the ABP/PPG cardiac cycle second derivative Cpeak duration with respect to Epeak to the ABP/PPG cardiac cycle second derivative Apeak duration with respect to Dpeak |
| 819 | ABP/PPG_D_RT_SD_Dpeak_wrtEpeak_ABP/PPG_D_SD_Apeak_wrtDpeak | The ratio of the ABP/PPG cardiac cycle second derivative Dpeak duration with respect to Epeak to the ABP/PPG cardiac cycle second derivative Apeak duration with respect to Dpeak |
| 820 | ABP/PPG_D_RT_SD_Bpeak_wrtCpeak_ABP/PPG_D_SD_Apeak_wrtEpeak | The ratio of the ABP/PPG cardiac cycle second derivative Bpeak duration with respect to Cpeak to the ABP/PPG cardiac cycle second derivative Apeak duration with respect to Epeak |
| 821 | ABP/PPG_D_RT_SD_Bpeak_wrtDpeak_ABP/PPG_D_SD_Apeak_wrtEpeak | The ratio of the ABP/PPG cardiac cycle second derivative Bpeak duration with respect to Dpeak to the ABP/PPG cardiac cycle second derivative Apeak duration with respect to Epeak |
| 822 | ABP/PPG_D_RT_SD_Bpeak_wrtEpeak_ABP/PPG_D_SD_Apeak_wrtEpeak | The ratio of the ABP/PPG cardiac cycle second derivative Bpeak duration with respect to Epeak to the ABP/PPG cardiac cycle second derivative Apeak duration with respect to Epeak |
| 823 | ABP/PPG_D_RT_SD_Cpeak_wrtDpeak_ABP/PPG_D_SD_Apeak_wrtEpeak | The ratio of the ABP/PPG cardiac cycle second derivative Cpeak duration with respect to Dpeak to the ABP/PPG cardiac cycle second derivative Apeak duration with respect to Epeak |
| 824 | ABP/PPG_D_RT_SD_Cpeak_wrtEpeak_ABP/PPG_D_SD_Apeak_wrtEpeak | The ratio of the ABP/PPG cardiac cycle second derivative Cpeak duration with respect to Epeak to the ABP/PPG cardiac cycle second derivative Apeak duration with respect to Epeak |
| 825 | ABP/PPG_D_RT_SD_Dpeak_wrtEpeak_ABP/PPG_D_SD_Apeak_wrtEpeak | The ratio of the ABP/PPG cardiac cycle second derivative Dpeak duration with respect to Epeak to the ABP/PPG cardiac cycle second derivative Apeak duration with respect to Epeak |
| 826 | ABP/PPG_D_RT_SD_Bpeak_wrtDpeak_ABP/PPG_D_SD_Bpeak_wrtCpeak | The ratio of the ABP/PPG cardiac cycle second derivative Bpeak duration with respect to Dpeak to the ABP/PPG cardiac cycle second derivative Bpeak duration with respect to Cpeak |
| 827 | ABP/PPG_D_RT_SD_Bpeak_wrtEpeak_ABP/PPG_D_SD_Bpeak_wrtCpeak | The ratio of the ABP/PPG cardiac cycle second derivative Bpeak duration with respect to Epeak to the ABP/PPG cardiac cycle second derivative Bpeak duration with respect to Cpeak |
| 828 | ABP/PPG_D_RT_SD_Cpeak_wrtDpeak_ABP/PPG_D_SD_Bpeak_wrtCpeak | The ratio of the ABP/PPG cardiac cycle second derivative Cpeak duration with respect to Dpeak to the ABP/PPG cardiac cycle second derivative Bpeak duration with respect to Cpeak |
| 829 | ABP/PPG_D_RT_SD_Cpeak_wrtEpeak_ABP/PPG_D_SD_Bpeak_wrtCpeak | The ratio of the ABP/PPG cardiac cycle second derivative Cpeak duration with respect to Epeak to the ABP/PPG cardiac cycle second derivative Bpeak duration with respect to Cpeak |
| 830 | ABP/PPG_D_RT_SD_Dpeak_wrtEpeak_ABP/PPG_D_SD_Bpeak_wrtCpeak | The ratio of the ABP/PPG cardiac cycle second derivative Dpeak duration with respect to Epeak to the ABP/PPG cardiac cycle second derivative Bpeak duration with respect to Cpeak |
| 831 | ABP/PPG_D_RT_SD_Bpeak_wrtEpeak_ABP/PPG_D_SD_Bpeak_wrtDpeak | The ratio of the ABP/PPG cardiac cycle second derivative Bpeak duration with respect to Epeak to the ABP/PPG cardiac cycle second derivative Bpeak duration with respect to Dpeak |
| 832 | ABP/PPG_D_RT_SD_Cpeak_wrtDpeak_ABP/PPG_D_SD_Bpeak_wrtDpeak | The ratio of the ABP/PPG cardiac cycle second derivative Cpeak duration with respect to Dpeak to the ABP/PPG cardiac cycle second derivative Bpeak duration with respect to Dpeak |
| 833 | ABP/PPG_D_RT_SD_Cpeak_wrtEpeak_ABP/PPG_D_SD_Bpeak_wrtDpeak | The ratio of the ABP/PPG cardiac cycle second derivative Cpeak duration with respect to Epeak to the ABP/PPG cardiac cycle second derivative Dpeak duration with respect to Cpeak |
| 834 | ABP/PPG_D_RT_SD_Dpeak_wrtEpeak_ABP/PPG_D_SD_Bpeak_wrtDpeak | The ratio of the ABP/PPG cardiac cycle second derivative Dpeak duration with respect to Epeak to the ABP/PPG cardiac cycle second derivative Bpeak duration with respect to Dpeak |
| 835 | ABP/PPG_D_RT_SD_Cpeak_wrtDpeak_ABP/PPG_D_SD_Bpeak_wrtEpeak | The ratio of the ABP/PPG cardiac cycle second derivative Cpeak duration with respect to Dpeak to the ABP/PPG cardiac cycle second derivative Bpeak duration with respect to Epeak |
| 836 | ABP/PPG_D_RT_SD_Cpeak_wrtEpeak_ABP/PPG_D_SD_Bpeak_wrtEpeak | The ratio of the ABP/PPG cardiac cycle second derivative Cpeak duration with respect to Epeak to the ABP/PPG cardiac cycle second derivative Dpeak duration with respect to Epeak |
| 837 | ABP/PPG_D_RT_SD_Dpeak_wrtEpeak_ABP/PPG_D_SD_Bpeak_wrtEpeak | The ratio of the ABP/PPG cardiac cycle second derivative Dpeak duration with respect to Epeak to the ABP/PPG cardiac cycle second derivative Bpeak duration with respect to Epeak |
| 838 | ABP/PPG_D_RT_SD_Cpeak_wrtEpeak_ABP/PPG_D_SD_Cpeak_wrtDpeak | The ratio of the ABP/PPG cardiac cycle second derivative Cpeak duration with respect to Epeak to the ABP/PPG cardiac cycle second derivative Cpeak duration with respect to Dpeak |
| 839 | ABP/PPG_D_RT_SD_Dpeak_wrtEpeak_ABP/PPG_D_SD_Cpeak_wrtDpeak | The ratio of the ABP/PPG cardiac cycle second derivative Dpeak duration with respect to Epeak to the ABP/PPG cardiac cycle second derivative Cpeak duration with respect to Dpeak |
| 840 | ABP/PPG_D_RT_SD_Dpeak_wrtEpeak_ABP/PPG_D_SD_Cpeak_wrtEpeak | The ratio of the ABP/PPG cardiac cycle second derivative Dpeak duration with respect to Epeak to the ABP/PPG cardiac cycle second derivative Cpeak duration with respect to Epeak |

27. **Frequency domain features (841-852)**

| 841 | ABP/PPG_AM_P1 | The amplitude of the first peak observed in the FFT of the ABP/PPG cardiac cycle |
| --- | --- | --- |
| 842 | ABP/PPG_AM_P2 | The amplitude of the second peak observed in the FFT of the ABP/PPG cardiac cycle |
| 843 | ABP/PPG_AM_P3 | The amplitude of the third peak observed in the FFT of the ABP/PPG cardiac cycle |
| 844 | ABP/PPG_F_F1 | The frequency corresponding to the first peak in the FFT of the ABP/PPG cardiac cycle |
| 845 | ABP/PPG_F_F2 | The frequency corresponding to the second peak in the FFT of the ABP/PPG cardiac cycle |
| 846 | ABP/PPG_F_F3 | The frequency corresponding to the third peak in the FFT of the ABP/PPG cardiac cycle |
| 847 | ABP/PPG_RT_ AM_P1_P2 | The ratio of the first peak amplitude to that of the second peak amplitude in the FFT of the ABP/PPG cardiac cycle |
| 848 | ABP/PPG_RT_ AM_P1_P3 | The ratio of the first peak amplitude to that of the third peak amplitude in the FFT of the ABP/PPG cardiac cycle |
| 849 | ABP/PPG_RT_ AM_P2_P3 | The ratio of the second peak amplitude to that of the third peak amplitude in the FFT of the ABP/PPG cardiac cycle |
| 850 | ABP/PPG_RT_ F_F1_F2 | The ratio of the frequency values of the first peak and the second peak in the FFT of the ABP/PPG cardiac cycle |
| 851 | ABP/PPG_RT_ F_F1_F3 | The ratio of the frequency values of the first peak and the third peak in the FFT of the ABP/PPG cardiac cycle |
| 852 | ABP/PPG_RT_ F_F2_F3 | The ratio of the frequency values of the second peak and the third peak in the FFT of the ABP/PPG cardiac cycle |
